# Combined sleep, domain-specific physical activity, and nutrition in relation to all-cause mortality among US adults

**DOI:** 10.64898/2026.07.23.26358753

**Authors:** Qingyang Lu, Nicholas A. Koemel, Raaj Kishore Biswas, Wenxin Bian, Joshua F. Wiley, Matthew P. Pase, Sean P.A. Drummond, Patrick Olivier, Margaret Allman Farinelli, Peter A. Cistulli, Stephen J. Simpson, Matthew N. Ahmadi, Emmanuel Stamatakis

## Abstract

**Background:** Sleep duration, physical activity, and nutrition (SPAN) are each associated with mortality risk, yet they are usually examined individually or in pairs, with limited attention to their combined associations and differences across physical activity domains. We examined combined SPAN behaviours in relation to all-cause mortality across different physical activity domains (occupational physical activity (OPA), leisure-time physical activity (LTPA) and transportational physical activity (TPA)).

**Methods:** We included adults aged 18 years or older from the National Health and Nutrition Examination Survey 2007–2018. Sleep duration was self-reported in hours/day. Physical activity, including moderate-to-vigorous physical activity (MVPA) across domains, was assessed using the Global Physical Activity Questionnaire. Diet quality was estimated from 24-hour dietary recalls using the Healthy Eating Index-2020 (HEI-2020). We created 27 mutually exclusive joint categories using tertiles of physical activity and HEI-2020, and guideline-based sleep-duration groups (<7, 7–8, and >8 h/day), with the lowest combination as the reference. We used Cox proportional hazards models to estimate hazard ratios (HRs) and 95% CIs for all-cause mortality. To estimate the minimum combined variations associated with lower mortality risk, we used the 5^th^ percentile of each behaviour as the reference.

**Results:** Among 31,875 participants (median age 48.0 years; 51.4% female), 2,623 deaths occurred over a median follow-up of 6.75 years. In joint categorical analyses, a combination of optimal sleep (7-8 h/day), high diet quality (HEI-2020: >56.0), and high LTPA (>35.4 min/day) or high TPA (>25.7 min/day) was associated with the lowest all-cause mortality risk (HR = 0.44, 95%CI: 0.29-0.65; HR = 0.54, 95%CI: 0.37-0.77). The lowest HR for OPA was observed for sleep duration (3.0–6.5 h/day), moderate OPA (1.4–114.0 min/day), and high diet quality (HR = 0.53, 95%CI: 0.37-0.75). Relative to the reference values (sleep: 5 h/day, MVPA/PA domains: 0 min/day and HEI-2020: 32.29), a combined minimum increment of 15 min/day of sleep, 5 HEI-2020 points, in combination with either 10.8 min/day of total MVPA, 24.0 min/day of OPA, or 5.2 min/day of LTPA, were associated with 10% lower all-cause mortality. For TPA, the 10% lower risk corresponded to 30 additional min/day of sleep, 5 HEI-2020 points, and 5.5 min/day of transport PA.

**Conclusions:** Modest combined increments in SPAN were associated with lower all-cause mortality among US adults across physical activity domains, with combinations including higher amounts of LTPA showing the most favourable profile.

## Introduction

Multi-behaviour lifestyle studies have explored that combinations of health behaviours, such as sleep, physical activity, diet, alcohol use, and smoking, are associated with chronic diseases, mortality, and life expectancy(1–3). These findings support the value of assessing health behaviours jointly rather than in isolation. Among these behaviours, sleep, physical activity, and nutrition (SPAN) represents a modifiable behavioural triad because they are repeated daily, biologically interdependent, and central to energy balance and circadian regulation that impact a broad spectrum of health outcomes and the risk of premature mortality (4–7).

Previous studies have explored the two-way joint associations of individual SPAN behaviours with mortality, such as diet and physical activity (PA) (8–10), PA and sleep(11, 12), and sleep and diet(13, 14). However, few studies have focused on all three SPAN behaviours and evaluated their potential synergistic associations with mortality risk(15–18). Recent SPAN studies (15–17) using self-reported diet quality and accelerometer-derived measures of sleep duration and moderate-to-vigorous physical activity (MVPA) showed that these behaviours, in combination, were associated with meaningful reductions in chronic diseases and premature death at substantial lower doses than when investigated individually. However, much of this evidence was derived from the UK Biobank, a cohort that is potentially healthier and less socioeconomically heterogeneous than the general UK population(19–22). Complementary evidence from the National Health and Nutrition Examination Survey (NHANES) may, therefore, help assess associations between SPAN and mortality beyond the context of relatively healthy volunteer cohorts(23).

Additionally, existing SPAN studies have generally examined physical activity in a domain-agnostic manner, typically using total MVPA (15–17). This may overlook the possibility that the health associations of PA depend not only on total volume, but also on how activity is accumulated across daily life (24, 25). Emerging evidence suggests that physical activity accumulation patterns, including incidental activity, bout patterns, and domain-specific activity including occupational physical activity (OPA), leisure-time physical activity (LTPA), and transportational physical activity (TPA), may have distinct associations with mortality and cardiovascular outcomes (24–31). In particular, higher levels of LTPA have been consistently associated with lower risks of all-cause mortality(28, 32, 33), while evidence for OPA and TPA has been mixed(32, 34–36). Clarifying the associations between domains of physical activity and mortality within a SPAN combination may help refine our understanding of physical activity–mortality relationships and identify more context-specific behavioural patterns.

Therefore, we aimed to explore the associations between combined SPAN behaviours and all-cause mortality risk in a sample of US adults while considering both total MVPA and domain of physical activity(16).

## Methods

### Study population

We used data from NHANES, which recruited participants in annual cycles from 2007 to 2018(37). NHANES is a continuous biennial US cross-sectional survey combining household interviews and in-person examinations to assess sociodemographic factors, health behaviours, diet, and physiological measures(38, 39). Written informed consent was obtained from all participants before participation in NHANES(40). We included participants aged 18 years or older with complete eligible data on SPAN behaviours. To reduce the risk of reverse causation, we excluded participants with recorded mortality within the first year of follow-up.

### Mortality Ascertainment

The mortality data were extracted from the National Death Index (NDI) with linkage through to 31 December 2019(41). The underlying causes of death were coded according to the International Statistical Classification of Diseases and Related Health Problems, Tenth Revision (ICD-10). Participants were followed from the date of survey participation until death or the end of follow-up, whichever occurred first.

### Sleep, physical activity, and nutrition assessment

Sleep duration was derived from self-reported questionnaire data. For 2007–2014, it was assessed as hours per day based on the question “How much sleep do you usually get at night on weekdays or workdays?” For 2016–2018, sleep duration was calculated from reported sleep and wake times(42). Physical activity data and corresponding domains of physical activity were estimated using the Global Physical Activity Questionnaire (GPAQ) in NHANES(43). Transportational physical activity was classified as moderate physical activity (MPA) because NHANES analytic guidelines assign it a metabolic equivalent (MET) value of 4.0, which falls within the standard moderate-intensity range (3.0–5.9 METs)(44). NHANES includes separate questions on MPA and vigorous physical activity (VPA) for OPA and LTPA; we therefore calculated MVPA for both OPA and LTPA. We calculated MVPA by summing all MPA minutes and twice the duration of VPA, consistent with previous NHANES studies(45–49). Total MVPA was then derived by combining TPA-derived MPA with MVPA accumulated through OPA and LTPA. NHANES assesses PA using weekly frequency, reported as days per week, and duration, reported as minutes per day. We therefore calculated weekly volume first and then divided it by seven to estimate average daily minutes of PA.

Dietary quality was evaluated using data from 24-hour dietary recalls. We assessed diet quality using the Healthy Eating Index–2020 (HEI-2020), which aligns with the Dietary Guidelines for Americans and has been validated in the US population and NHANES(50–52). HEI-2020 comprises 13 dietary components, each scored on a maximum of 5 or 10 points, producing a total score from 0 to 100, with higher scores indicating greater adherence to dietary recommendations(50). We calculated the mean of HEI-2020 if two-day recall data were available or otherwise used the first day’s data for HEI-2020 calculations.

### Statistical analysis

We used Cox proportional hazards models to estimate associations between SPAN and all-cause mortality. Proportional hazards assumptions were evaluated and met by Schoenfeld residuals. To minimise the influence of extreme values, sleep duration was winsorised at the 1^st^ and 99^th^ percentiles (53) and MPA and VPA were winsorised at the 90^th^ percentile based on their empirical distributions(54), as PA were highly right-skewed.

We categorised each SPAN behaviour into three levels and conducted joint categorical analyses (27 mutually exclusive groups) to identify associations of combined SPAN and all-cause mortality. We divided HEI-2020 and total MVPA into tertiles (low, moderate, high). For PA domains, due to highly right-skewed distributions, we divided each domain into 0 minutes (low), the bottom 50% of non-zero values (moderate), and the top 50% of non-zero values (high). We followed the recommendations from the American Academy of Sleep Medicine to divide sleep duration as <7 hours (short), 7– 8 hours (optimal), and >8 hours (long) (55, 56). The reference group was defined as low HEI-2020, short sleep duration, and low PA.

We estimated the minimum variations in combined SPAN behaviours by identifying the smallest joint increments from a 5^th^ percentile baseline associated with a clinically meaningful reduction in all-cause mortality risk. Consistent with previous studies(15–17), we defined a clinically meaningful lower risk as a 10% reduction, or the nearest predicted integer and estimated the minimum combined or individual increments of SPAN corresponding to predicted hazard. Clinically meaningful reductions were determined using thresholds commonly applied in clinical trials to minimise reliance or less robust estimations due to residual confounding or regression dilution bias (15–17). In this study, the reference point was sleep duration: 5 hours/day; physical activity: 0 minutes/day across MVPA and PA domains; diet quality: 32.29 HEI-2020 points. We used restricted cubic spline models to account for potential non-linear associations between SPAN behaviours and all-cause mortality in the minimum-variation analyses (15–17).

Models were adjusted for age, sex, ethnicity, education, ratio of family income to poverty, alcohol consumption, smoking status, self-reported sedentary activity time, total energy intake, previous diagnosis of cardiovascular disease (CVD), cancer or type 2 diabetes (T2D), and family history of heart attack and diabetes. When examining each PA domain as the primary exposure, we mutually adjusted for the other two domains in the multivariable models to reduce potential confounding. Multiple imputation (5 imputed times) was performed for covariates if missingness was observed(57). All analyses were conducted using R (version 4.5.1). Reporting followed Strengthening the Reporting of Observational Studies in Epidemiology (STROBE) guidelines.

### Sensitivity analysis

We repeated the main analyses, excluding participants with only one 24 h dietary recall. We additionally adjusted for body mass index (BMI) in our sensitivity analysis, a variable that may be on the causal pathway as a mediator of SPAN and all-cause mortality. We excluded participants with baseline CVD and cancer in our sensitivity analysis. We repeated the analyses using the Alternate Healthy Eating Index(58, 59), and after excluding underweight participants (BMI <18.5 kg/m²), current smokers, and those reporting poor health(60). To preserve sample size while accounting for information from two dietary recalls, we incorporated the NHANES complex survey design in sensitivity analyses only, using separate weighting approaches according to NHANES analytic guidelines(38). We excluded participants with sex-specific implausible daily energy intakes (<600 or >3500 kcal/day for women; <800 or >4200 kcal/day for men)(61). We also conducted subgroup analyses by sex, race and ethnicity, education, and ratio of family income to poverty in categorical analyses for combined SPAN behaviours. We repeated the analyses using multiple imputation with 10 times and complete-case data. We also tested a WHO guideline-based categorisation of MVPA as low (0 min/day), moderate (1.4–20.7 min/day), and high (≥21.4 min/day, approximately ≥150 min/week).

## Results

### Study participants

The core analytical sample included 31,875 participants (median age [IQR], 48.0 [32.0–63.0] years; 51.4% women), among whom 2,623 all-cause deaths were recorded (**Table 1 and Supplementary Figure 1**). Following exclusion of deaths occurring within the first year of follow-up, the median follow-up duration was 6.75 years (IQR: 3.83–9.83). Median total MVPA was 34.3 min/day (IQR: 0.0-107.1). Across physical activity domains, median OPA, LTPA, and TPA were 0.0 min/day (IQR: 0.0-68.6), 0.0 min/day (IQR: 0.0-34.3), and 0.0 min/day (IQR: 0.0-4.3), respectively. Median sleep duration was 7.0 h/day (IQR: 6.0-8.0), and median HEI-2020 score was 50.25 (IQR: 42.2-59.3).

**Table 1.**
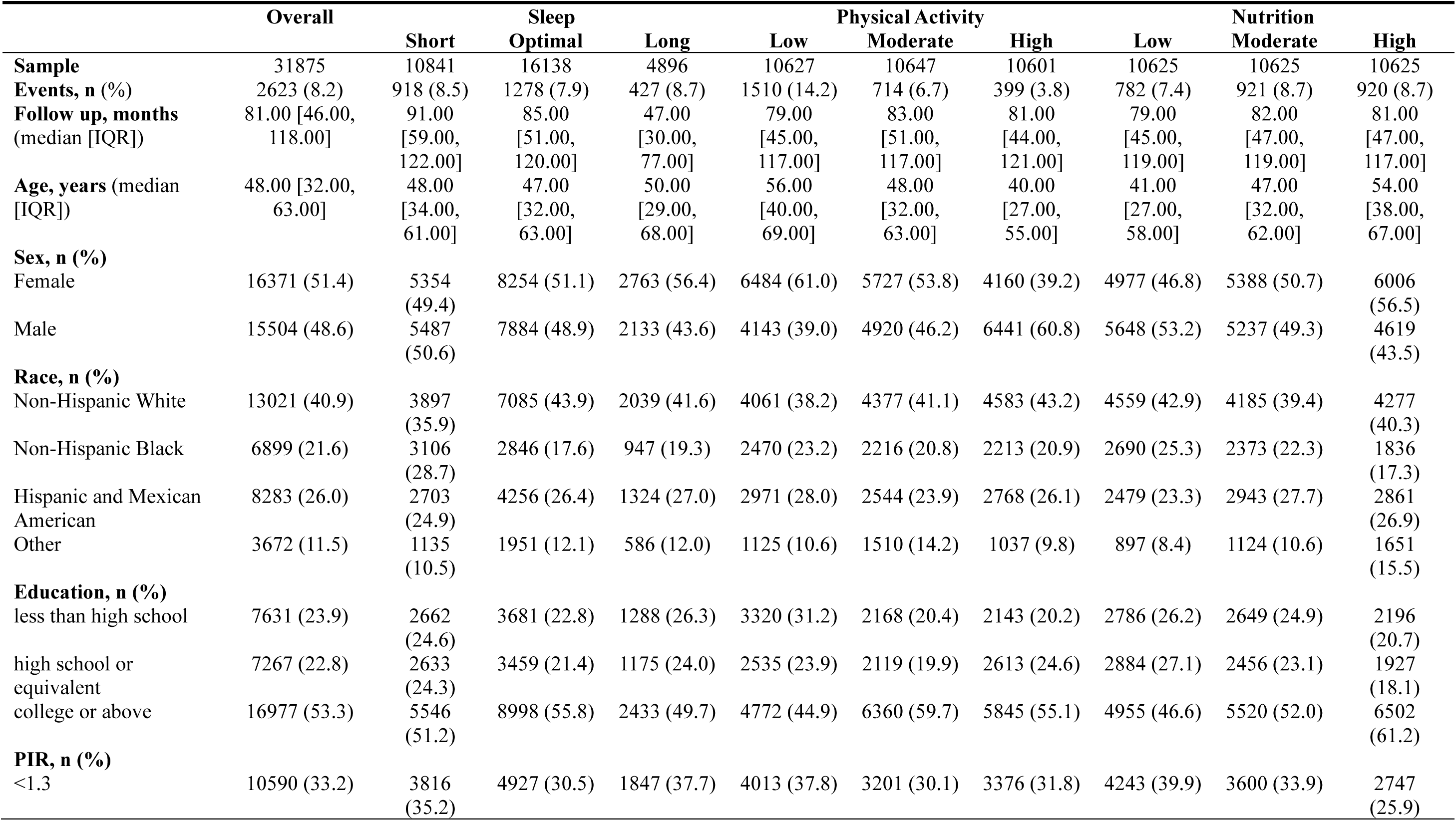
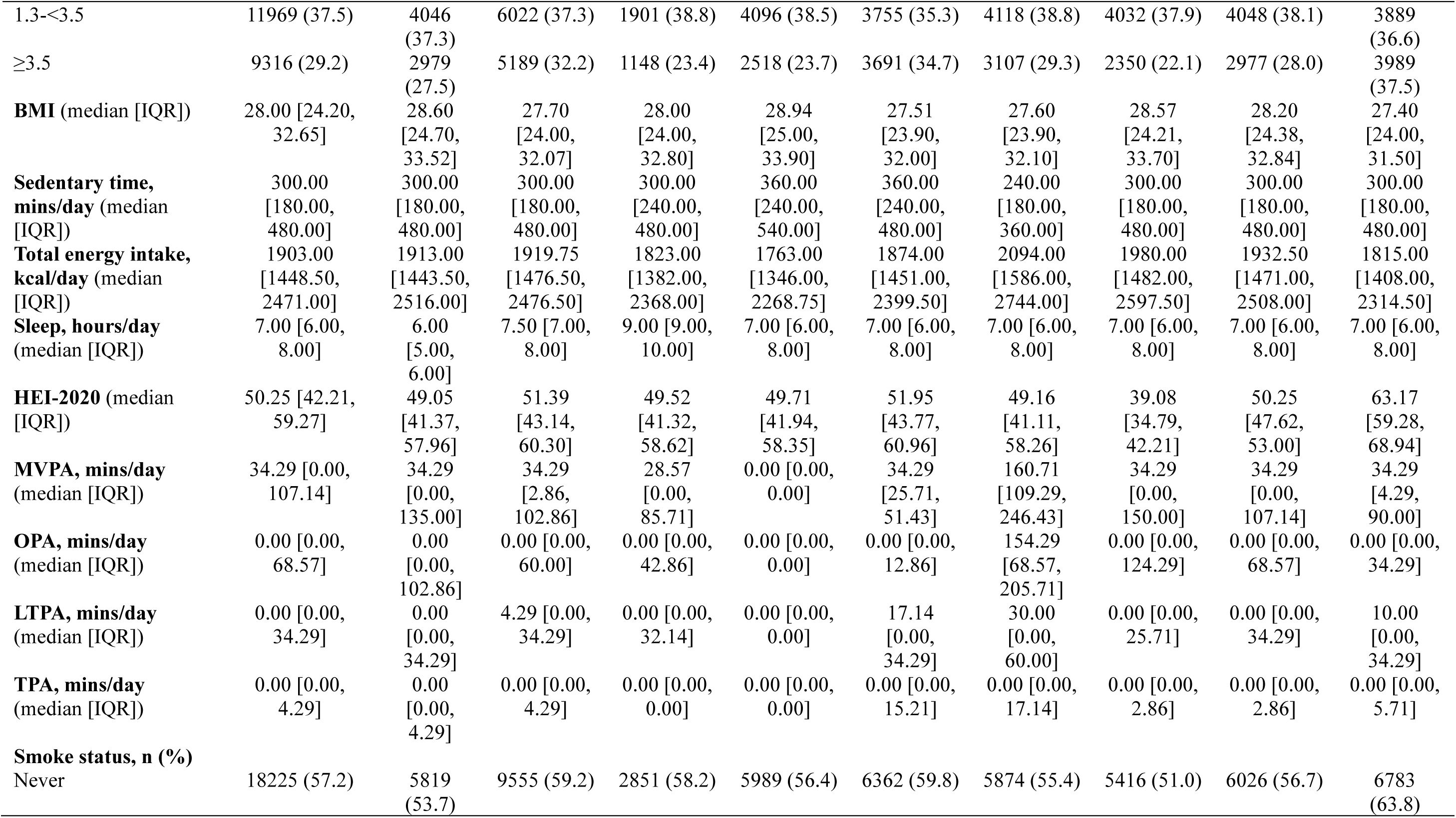
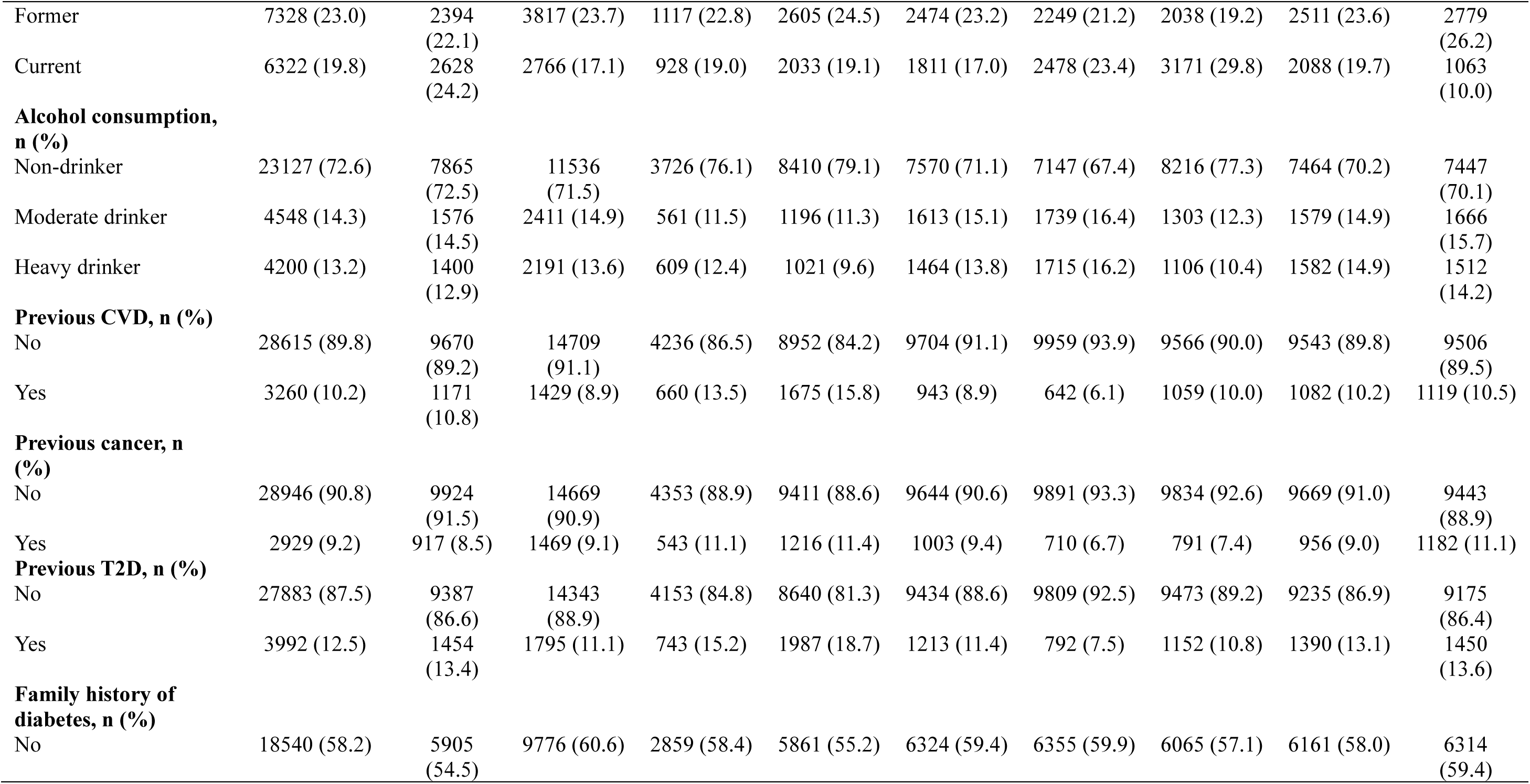
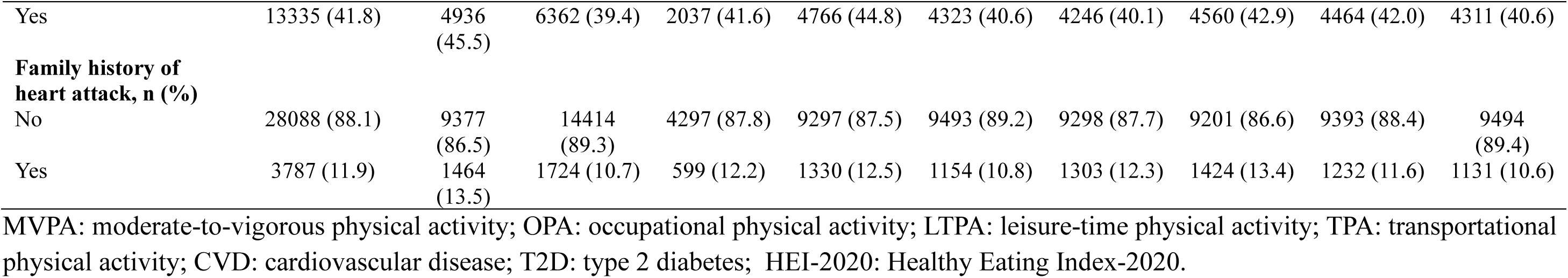
Participants’ characteristics by sleep duration, physical activity and nutrition.

|  | Overall |  | Sleep |  |  | Physical Activity |  |  | Nutrition |  |
| --- | --- | --- | --- | --- | --- | --- | --- | --- | --- | --- |
|  |  | Short | Optimal | Long | Low | Moderate | High | Low | Moderate | High |
| **Sample** | 31875 | 10841 | 16138 | 4896 | 10627 | 10647 | 10601 | 10625 | 10625 | 10625 |
| **Events, n (%)** | 2623 (8.2) | 918 (8.5) | 1278 (7.9) | 427 (8.7) | 1510 (14.2) | 714 (6.7) | 399 (3.8) | 782 (7.4) | 921 (8.7) | 920 (8.7) |
| **Follow up, months**<br>(median [IQR]) | 81.00 [46.00,<br>118.00] | 91.00<br>[59.00,<br>122.00] | 85.00<br>[51.00,<br>120.00] | 47.00<br>[30.00,<br>77.00] | 79.00<br>[45.00,<br>117.00] | 83.00<br>[51.00,<br>117.00] | 81.00<br>[44.00,<br>121.00] | 79.00<br>[45.00,<br>119.00] | 82.00<br>[47.00,<br>119.00] | 81.00<br>[47.00,<br>117.00] |
| **Age, years** (median<br>[IQR]) | 48.00 [32.00,<br>63.00] | 48.00<br>[34.00,<br>61.00] | 47.00<br>[32.00,<br>63.00] | 50.00<br>[29.00,<br>68.00] | 56.00<br>[40.00,<br>69.00] | 48.00<br>[32.00,<br>63.00] | 40.00<br>[27.00,<br>55.00] | 41.00<br>[27.00,<br>58.00] | 47.00<br>[32.00,<br>62.00] | 54.00<br>[38.00,<br>67.00] |
| **Sex, n (%)** |  |  |  |  |  |  |  |  |  |  |
| Female | 16371 (51.4) | 5354<br>(49.4) | 8254 (51.1) | 2763 (56.4) | 6484 (61.0) | 5727 (53.8) | 4160 (39.2) | 4977 (46.8) | 5388 (50.7) | 6006<br>(56.5) |
| Male | 15504 (48.6) | 5487<br>(50.6) | 7884 (48.9) | 2133 (43.6) | 4143 (39.0) | 4920 (46.2) | 6441 (60.8) | 5648 (53.2) | 5237 (49.3) | 4619<br>(43.5) |
| **Race, n (%)** |  |  |  |  |  |  |  |  |  |  |
| Non-Hispanic White | 13021 (40.9) | 3897<br>(35.9) | 7085 (43.9) | 2039 (41.6) | 4061 (38.2) | 4377 (41.1) | 4583 (43.2) | 4559 (42.9) | 4185 (39.4) | 4277<br>(40.3) |
| Non-Hispanic Black | 6899 (21.6) | 3106<br>(28.7) | 2846 (17.6) | 947 (19.3) | 2470 (23.2) | 2216 (20.8) | 2213 (20.9) | 2690 (25.3) | 2373 (22.3) | 1836<br>(17.3) |
| Hispanic and Mexican<br>American | 8283 (26.0) | 2703<br>(24.9) | 4256 (26.4) | 1324 (27.0) | 2971 (28.0) | 2544 (23.9) | 2768 (26.1) | 2479 (23.3) | 2943 (27.7) | 2861<br>(26.9) |
| Other | 3672 (11.5) | 1135<br>(10.5) | 1951 (12.1) | 586 (12.0) | 1125 (10.6) | 1510 (14.2) | 1037 (9.8) | 897 (8.4) | 1124 (10.6) | 1651<br>(15.5) |
| **Education, n (%)** |  |  |  |  |  |  |  |  |  |  |
| less than high school | 7631 (23.9) | 2662<br>(24.6) | 3681 (22.8) | 1288 (26.3) | 3320 (31.2) | 2168 (20.4) | 2143 (20.2) | 2786 (26.2) | 2649 (24.9) | 2196<br>(20.7) |
| high school or<br>equivalent | 7267 (22.8) | 2633<br>(24.3) | 3459 (21.4) | 1175 (24.0) | 2535 (23.9) | 2119 (19.9) | 2613 (24.6) | 2884 (27.1) | 2456 (23.1) | 1927<br>(18.1) |
| college or above | 16977 (53.3) | 5546<br>(51.2) | 8998 (55.8) | 2433 (49.7) | 4772 (44.9) | 6360 (59.7) | 5845 (55.1) | 4955 (46.6) | 5520 (52.0) | 6502<br>(61.2) |
| **PIR, n (%)** |  |  |  |  |  |  |  |  |  |  |
| <1.3 | 10590 (33.2) | 3816<br>(35.2) | 4927 (30.5) | 1847 (37.7) | 4013 (37.8) | 3201 (30.1) | 3376 (31.8) | 4243 (39.9) | 3600 (33.9) | 2747<br>(25.9) |

**Table 1 (Continued)** Participants' characteristics by sleep duration, physical activity and nutrition.
|  | Overall | Short | Sleep<br>Optimal | Long | Low | Physical Activity<br>Moderate | High | Low | Nutrition<br>Moderate | High |
| --- | --- | --- | --- | --- | --- | --- | --- | --- | --- | --- |
| 1.3-<3.5 | 11969 (37.5) | 4046<br>(37.3) | 6022 (37.3) | 1901 (38.8) | 4096 (38.5) | 3755 (35.3) | 4118 (38.8) | 4032 (37.9) | 4048 (38.1) | 3889<br>(36.6) |
| ≥3.5 | 9316 (29.2) | 2979<br>(27.5) | 5189 (32.2) | 1148 (23.4) | 2518 (23.7) | 3691 (34.7) | 3107 (29.3) | 2350 (22.1) | 2977 (28.0) | 3989<br>(37.5) |
| **BMI** (median [IQR]) | 28.00 [24.20,<br>32.65] | 28.60<br>[24.70,<br>33.52] | 27.70<br>[24.00,<br>32.07] | 28.00<br>[24.00,<br>32.80] | 28.94<br>[25.00,<br>33.90] | 27.51<br>[23.90,<br>32.00] | 27.60<br>[23.90,<br>32.10] | 28.57<br>[24.21,<br>33.70] | 28.20<br>[24.38,<br>32.84] | 27.40<br>[24.00,<br>31.50] |
| **Sedentary time,**<br>**mins/day** (median<br>[IQR]) | 300.00<br>[180.00,<br>480.00] | 300.00<br>[180.00,<br>480.00] | 300.00<br>[180.00,<br>480.00] | 300.00<br>[240.00,<br>480.00] | 360.00<br>[240.00,<br>540.00] | 360.00<br>[240.00,<br>480.00] | 240.00<br>[180.00,<br>360.00] | 300.00<br>[180.00,<br>480.00] | 300.00<br>[180.00,<br>480.00] | 300.00<br>[180.00,<br>480.00] |
| **Total energy intake,**<br>**kcal/day** (median<br>[IQR]) | 1903.00<br>[1448.50,<br>2471.00] | 1913.00<br>[1443.50,<br>2516.00] | 1919.75<br>[1476.50,<br>2476.50] | 1823.00<br>[1382.00,<br>2368.00] | 1763.00<br>[1346.00,<br>2268.75] | 1874.00<br>[1451.00,<br>2399.50] | 2094.00<br>[1586.00,<br>2744.00] | 1980.00<br>[1482.00,<br>2597.50] | 1932.50<br>[1471.00,<br>2508.00] | 1815.00<br>[1408.00,<br>2314.50] |
| **Sleep, hours/day**<br>(median [IQR]) | 7.00 [6.00,<br>8.00] | 6.00<br>[5.00,<br>6.00] | 7.50 [7.00,<br>8.00] | 9.00 [9.00,<br>10.00] | 7.00 [6.00,<br>8.00] | 7.00 [6.00,<br>8.00] | 7.00 [6.00,<br>8.00] | 7.00 [6.00,<br>8.00] | 7.00 [6.00,<br>8.00] | 7.00 [6.00,<br>8.00] |
| **HEI-2020** (median<br>[IQR]) | 50.25 [42.21,<br>59.27] | 49.05<br>[41.37,<br>57.96] | 51.39<br>[43.14,<br>60.30] | 49.52<br>[41.32,<br>58.62] | 49.71<br>[41.94,<br>58.35] | 51.95<br>[43.77,<br>60.96] | 49.16<br>[41.11,<br>58.26] | 39.08<br>[34.79,<br>42.21] | 50.25<br>[47.62,<br>53.00] | 63.17<br>[59.28,<br>68.94] |
| **MVPA, mins/day**<br>(median [IQR]) | 34.29 [0.00,<br>107.14] | 34.29<br>[0.00,<br>135.00] | 34.29<br>[2.86,<br>102.86] | 28.57<br>[0.00,<br>85.71] | 0.00 [0.00,<br>0.00] | 34.29<br>[25.71,<br>51.43] | 160.71<br>[109.29,<br>246.43] | 34.29<br>[0.00,<br>150.00] | 34.29<br>[0.00,<br>107.14] | 34.29<br>[4.29,<br>90.00] |
| **OPA, mins/day**<br>(median [IQR]) | 0.00 [0.00,<br>68.57] | 0.00<br>[0.00,<br>102.86] | 0.00 [0.00,<br>60.00] | 0.00 [0.00,<br>42.86] | 0.00 [0.00,<br>0.00] | 0.00 [0.00,<br>12.86] | 154.29<br>[68.57,<br>205.71] | 0.00 [0.00,<br>124.29] | 0.00 [0.00,<br>68.57] | 0.00 [0.00,<br>34.29] |
| **LTPA, mins/day**<br>(median [IQR]) | 0.00 [0.00,<br>34.29] | 0.00<br>[0.00,<br>34.29] | 4.29 [0.00,<br>34.29] | 0.00 [0.00,<br>32.14] | 0.00 [0.00,<br>0.00] | 17.14<br>[0.00,<br>34.29] | 30.00<br>[0.00,<br>60.00] | 0.00 [0.00,<br>25.71] | 0.00 [0.00,<br>34.29] | 10.00<br>[0.00,<br>34.29] |
| **TPA, mins/day**<br>(median [IQR]) | 0.00 [0.00,<br>4.29] | 0.00<br>[0.00,<br>4.29] | 0.00 [0.00,<br>4.29] | 0.00 [0.00,<br>0.00] | 0.00 [0.00,<br>0.00] | 0.00 [0.00,<br>15.21] | 0.00 [0.00,<br>17.14] | 0.00 [0.00,<br>2.86] | 0.00 [0.00,<br>2.86] | 0.00 [0.00,<br>5.71] |
| **Smoke status, n (%)** |  |  |  |  |  |  |  |  |  |  |
| Never | 18225 (57.2) | 5819<br>(53.7) | 9555 (59.2) | 2851 (58.2) | 5989 (56.4) | 6362 (59.8) | 5874 (55.4) | 5416 (51.0) | 6026 (56.7) | 6783<br>(63.8) |

**Table 1 (Continued)** Participants' characteristics by sleep duration, physical activity and nutrition.
|  | Overall | Short | Sleep<br>Optimal | Long | Low | Physical Activity<br>Moderate | High | Low | Nutrition<br>Moderate | High |
| --- | --- | --- | --- | --- | --- | --- | --- | --- | --- | --- |
| Former | 7328 (23.0) | 2394<br>(22.1) | 3817 (23.7) | 1117 (22.8) | 2605 (24.5) | 2474 (23.2) | 2249 (21.2) | 2038 (19.2) | 2511 (23.6) | 2779<br>(26.2) |
| Current | 6322 (19.8) | 2628<br>(24.2) | 2766 (17.1) | 928 (19.0) | 2033 (19.1) | 1811 (17.0) | 2478 (23.4) | 3171 (29.8) | 2088 (19.7) | 1063<br>(10.0) |
| **Alcohol consumption, n (%)** |  |  |  |  |  |  |  |  |  |  |
| Non-drinker | 23127 (72.6) | 7865<br>(72.5) | 11536<br>(71.5) | 3726 (76.1) | 8410 (79.1) | 7570 (71.1) | 7147 (67.4) | 8216 (77.3) | 7464 (70.2) | 7447<br>(70.1) |
| Moderate drinker | 4548 (14.3) | 1576<br>(14.5) | 2411 (14.9) | 561 (11.5) | 1196 (11.3) | 1613 (15.1) | 1739 (16.4) | 1303 (12.3) | 1579 (14.9) | 1666<br>(15.7) |
| Heavy drinker | 4200 (13.2) | 1400<br>(12.9) | 2191 (13.6) | 609 (12.4) | 1021 (9.6) | 1464 (13.8) | 1715 (16.2) | 1106 (10.4) | 1582 (14.9) | 1512<br>(14.2) |
| **Previous CVD, n (%)** |  |  |  |  |  |  |  |  |  |  |
| No | 28615 (89.8) | 9670<br>(89.2) | 14709<br>(91.1) | 4236 (86.5) | 8952 (84.2) | 9704 (91.1) | 9959 (93.9) | 9566 (90.0) | 9543 (89.8) | 9506<br>(89.5) |
| Yes | 3260 (10.2) | 1171<br>(10.8) | 1429 (8.9) | 660 (13.5) | 1675 (15.8) | 943 (8.9) | 642 (6.1) | 1059 (10.0) | 1082 (10.2) | 1119 (10.5) |
| **Previous cancer, n (%)** |  |  |  |  |  |  |  |  |  |  |
| No | 28946 (90.8) | 9924<br>(91.5) | 14669<br>(90.9) | 4353 (88.9) | 9411 (88.6) | 9644 (90.6) | 9891 (93.3) | 9834 (92.6) | 9669 (91.0) | 9443<br>(88.9) |
| Yes | 2929 (9.2) | 917 (8.5) | 1469 (9.1) | 543 (11.1) | 1216 (11.4) | 1003 (9.4) | 710 (6.7) | 791 (7.4) | 956 (9.0) | 1182 (11.1) |
| **Previous T2D, n (%)** |  |  |  |  |  |  |  |  |  |  |
| No | 27883 (87.5) | 9387<br>(86.6) | 14343<br>(88.9) | 4153 (84.8) | 8640 (81.3) | 9434 (88.6) | 9809 (92.5) | 9473 (89.2) | 9235 (86.9) | 9175<br>(86.4) |
| Yes | 3992 (12.5) | 1454<br>(13.4) | 1795 (11.1) | 743 (15.2) | 1987 (18.7) | 1213 (11.4) | 792 (7.5) | 1152 (10.8) | 1390 (13.1) | 1450<br>(13.6) |
| **Family history of diabetes, n (%)** |  |  |  |  |  |  |  |  |  |  |
| No | 18540 (58.2) | 5905<br>(54.5) | 9776 (60.6) | 2859 (58.4) | 5861 (55.2) | 6324 (59.4) | 6355 (59.9) | 6065 (57.1) | 6161 (58.0) | 6314<br>(59.4) |

**Table 1 (Continued)** Participants' characteristics by sleep duration, physical activity and nutrition.
|  | Overall | Short | Sleep<br>Optimal | Long | Low | Physical Activity<br>Moderate | High | Low | Nutrition<br>Moderate | High |
| --- | --- | --- | --- | --- | --- | --- | --- | --- | --- | --- |
| Yes | 13335 (41.8) | 4936<br>(45.5) | 6362 (39.4) | 2037 (41.6) | 4766 (44.8) | 4323 (40.6) | 4246 (40.1) | 4560 (42.9) | 4464 (42.0) | 4311 (40.6) |
| **Family history of**<br>**heart attack, n (%)** |  |  |  |  |  |  |  |  |  |  |
| No | 28088 (88.1) | 9377<br>(86.5) | 14414<br>(89.3) | 4297 (87.8) | 9297 (87.5) | 9493 (89.2) | 9298 (87.7) | 9201 (86.6) | 9393 (88.4) | 9494<br>(89.4) |
| Yes | 3787 (11.9) | 1464<br>(13.5) | 1724 (10.7) | 599 (12.2) | 1330 (12.5) | 1154 (10.8) | 1303 (12.3) | 1424 (13.4) | 1232 (11.6) | 1131 (10.6) |

MVPA: moderate-to-vigorous physical activity; OPA: occupational physical activity; LTPA: leisure-time physical activity; TPA: transportational physical activity; CVD: cardiovascular disease; T2D: type 2 diabetes; HEI-2020: Healthy Eating Index-2020.

### Individual SPAN behaviours in relation to all-cause mortality

Overall, non-linear associations with all-cause mortality were observed for MVPA and sleep duration, whereas diet quality showed an approximately linear inverse association with all-cause mortality (**Supplementary Figure 2**). For physical activity domains, non-linear associations with all-cause mortality were observed for OPA and LTPA, whereas TPA showed a near-linear association (**Supplementary Figure 3**).

### Combined SPAN behaviours and all-cause mortality across PA domains

Compared with the reference group (**Figure 1**), the combination of optimal sleep duration (7-8 h/day), high MVPA (> 72.9 min/day), and moderate diet quality (HEI-2020: 45.0-56.0) was associated with the lowest risk of all-cause mortality (hazard ratio [HR]=0.50, 95% CI: 0.37–0.67). A similar association was observed for the combination of optimal sleep duration, high MVPA, and high diet quality (HEI-2020 > 56.0, HR = 0.51, 95% CI: 0.39–0.67).

**Figure 1.**
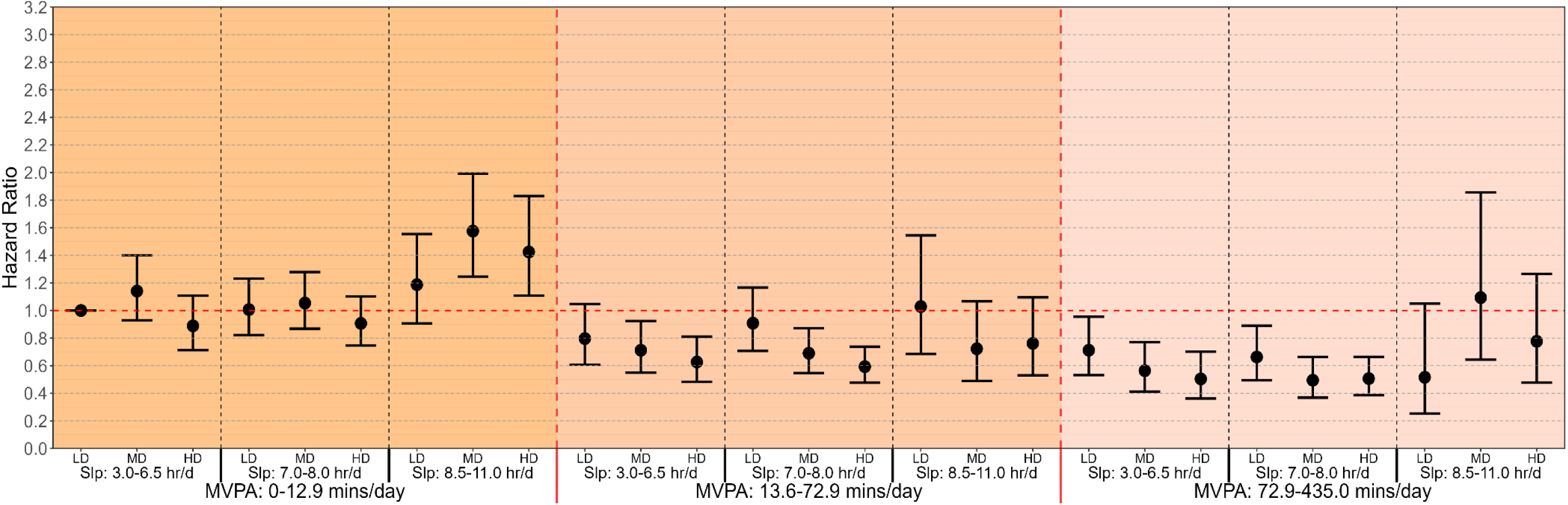
Multivariable-adjusted associations of combined sleep, moderate-to-vigorous physical activity (MVPA) and nutrition with all-cause mortality risk. **Legend**: Model is adjusted for age, sex, ethnicity, education, ratio of family income to poverty, alcohol consumption, smoking status, sedentary activity time, total energy intake, previous diagnosis of cardiovascular disease, cancer or type 2 diabetes, and family history of heart attack and diabetes (n = 31,875; events = 2,623). Sleep duration (hours/day), MVPA (minutes/day), and nutrition (Healthy Eating Index-2020 (HEI2020)) were treated as a joint term. The specific groups for each exposure were divided as sleep duration: 3.0-6.5 hours/day (short), 7.0-8.0 hours/day (optimal) and 8.5-11.0 hours/day (long); MVPA: 0-12.9 minutes/day (low), 13.6-72.9 minutes/day (moderate) and 72.9-435.0 minutes/day (high); HEI-2020: 10.0-45.0 (low), 45.0-56.0 (moderate), and 56.0-93.8 (high). The lowest group for all three exposures were treated as reference group. LD: low diet quality; MD: moderate diet quality; HD: high diet quality; Slp: sleep duration.

For OPA, compared to the reference group (**Figure 2A**), we observed the lowest all-cause mortality risk among participants with moderate OPA (1.4 – 111.5 min/day), short sleep (3.0-6.5 h/day), and high diet quality (HR=0.53, 95% CI: 0.37-0.75). We also found lower all-cause mortality with combinations of optimal sleep, high OPA (> 113.0 min/day), and moderate diet quality (HR=0.61, 95% CI: 0.43-0.85), or with high diet quality (HR=0.66, 95% CI: 0.47-0.92). For LTPA, compared to the reference group, we observed the lowest all-cause mortality risk in the combination of optimal sleep, high LTPA (> 35.4 min/day) and moderate diet quality (HR=0.44, 95% CI: 0.30-0.66, **Figure 2B**). In **Figure 2C**, we showed that the combination of optimal sleep, high TPA (> 25.7 min/day), and high diet quality was associated with the lowest all-cause mortality (HR=0.55, 95% CI: 0.38-0.79).

**Figure 2.**
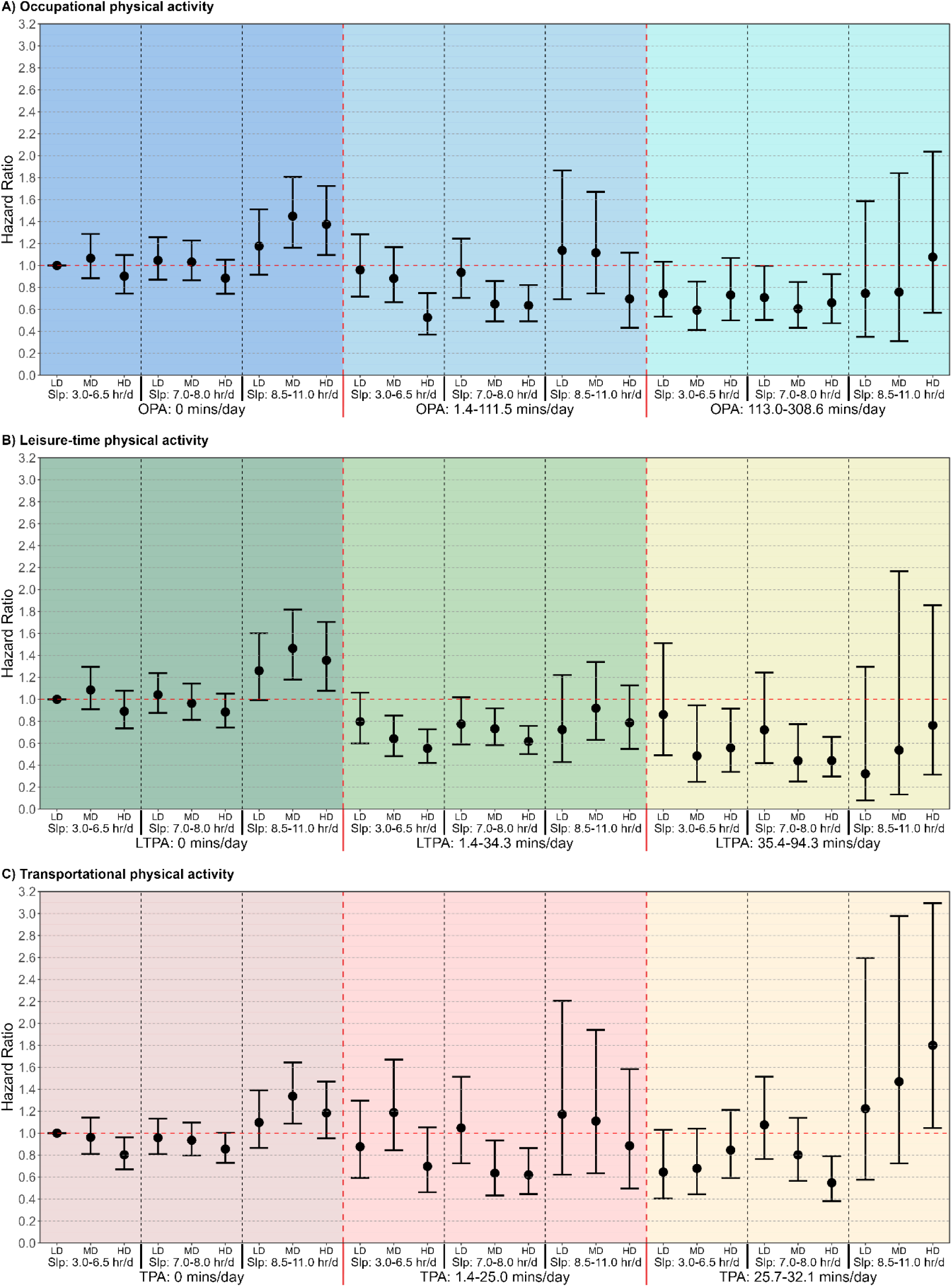
Multivariable-adjusted associations of combined sleep, physical activity domains and nutrition with all-cause mortality risk. A: Occupational physical activity (OPA); B: Leisure-time physical activity (LTPA); C: Transportational physical activity (TPA). **Legend:** Models are adjusted for adjusted for age, sex, ethnicity, education, ratio of family income to poverty, alcohol consumption, smoking status, sedentary activity time, total energy intake, previous diagnosis of cardiovascular disease, cancer or type 2 diabetes, and family history of heart attack and diabetes (n = 31,875; events = 2,623). When examining one physical activity domain as the primary exposure, the other two physical activity domains were adjusted in the multivariable models. Sleep duration (hours/day), OPA/LTPA/TPA (minutes/day), and nutrition (Healthy Eating Index-2020 (HEI2020)) were treated as a joint term. The specific groups for each exposure were divided as sleep duration: 3.0-6.5 hours/day (short), 7.0-8.0 hours/day (optimal) and 8.5-11.0 hours/day (long); OPA: 0 minutes/day (low), 1.4-111.5 minutes/day (moderate) and 113.0-308.6 minutes/day (high); LTPA: 0 minutes/day (low), 1.4-34.3 minutes/day (moderate) and 35.4-94.3 minutes/day (high); TPA: 0 minutes/day (low), 1.4-25.0 minutes/day (moderate) and 25.7-32.1 minutes/day (high); HEI-2020: 10.0-45.0 (low), 45.0-56.0 (moderate), and 56.0-93.8 (high). The lowest group for all three exposures in each physical activity domain were treated as reference group. LD: low diet quality; MD: moderate diet quality; HD: high diet quality; Slp: sleep duration.

### Minimum variations of SPAN behaviours and all-cause mortality

We present the minimum variations in SPAN behaviours associated with 10% to 60% lower predicted all-cause mortality risk in **Table 2**. Compared with the reference point, a combination of an additional 15 minutes of sleep, 10.8 minutes of MVPA, and 5-points in HEI-2020 was associated with a 10% lower predicted risk of all-cause mortality (hazard ratio [HR] = 0.90, 95% CI: 0.86-0.95). Across physical activity domains, similar minimum combined variations were observed for OPA, with an additional 15 minutes of sleep, 24.0 minutes of OPA, and 5 HEI-2020 points (HR = 0.90, 95% CI 0.86–0.96), and for LTPA, with an additional 15 minutes of sleep, 5.2 minutes of LTPA, and 5 HEI-2020 points (HR = 0.90, 95% CI 0.86–0.95). A combined variation of additional 30 minutes of sleep, 5.5 minutes of TPA and 5 points of diet quality was associated with 10% lower risk of all-cause mortality (HR = 0.90, 95% CI: 0.82-1.00).

**Table 2.** Minimum Concurrent or Individual Variations in Sleep, Physical Activity, and Nutrition Across Physical Activity Domains Associated With Clinically Meaningful Lower Risks of All-Cause Mortality Compared With the 5th Percentile of SPAN Exposures. **Legend:** minimum variations in combined or individual sleep, physical activity and nutrition behaviours in association with clinical meaningful lower risks of all-cause mortality. The variations in combined SPAN behaviours and corresponding mortality risk compared to the 5^th^ percentile of sleep (5.0 h/day), physical activity (0 min/day) for MVPA and all physical activity domains, and nutrition (HEI-2020: 32.29) in increments of 10% are shown in the left. The variation needed for individual SPAN behaviours is shown on the right for comparison. NA denotes that the vairation of combined or individual SPAN behaviours could not reach the level of lower risks. All models wre adjusted for age, sex, ethnicity, education, ratio of family income to poverty, alcohol consumption, smoking status, sedentary activity time, total energy intake, previous diagnosis of cardiovascular disease, cancer or type 2 diabetes, and family history of heart attack and diabetes. When examining one PA domain as the primary exposure, the other two physical activity domains were adjusted in the multivariable models.

| Combined SPAN (Minimum additional variations) |  |  |  | Individual SPAN (Minimum additional variations) |  |  |
| --- | --- | --- | --- | --- | --- | --- |
| MVPA | Sleep | PA | Nutrition | Sleep | PA | Nutrition |
| 0.9 (0.86-0.95) | 15 | 10.8 | 5 | 78 | 14.3 | 25<sup>#</sup> |
| 0.8 (0.73-0.88) | 30 | 24.1 | 10 | NA | 31.3 | 38 |
| 0.7 (0.62-0.80) | 45 | 39.8 | 15 | NA | 51.9 | 51 |
| 0.6 (0.52-0.71) | 60 | 60.0 | 20 | NA | 82.3 | 60<sup>b</sup> |
| 0.5 (0.42-0.60) | 75 | 92.4 | 25 | NA | 158.8<sup>a</sup> | NA |
| 0.4 (0.34-0.49) | 90 | 135.4 | 40 | NA | NA | NA |
| OPA | Sleep | PA | Nutrition | Sleep | PA | Nutrition |
| 0.9 (0.86-0.96) | 15 | 24.0 | 5 | 72<sup>c</sup> | 31.2 | 29<sup>#</sup> |
| 0.8 (0.72-0.90) | 30 | 54.5 | 10 | NA | 70.0 | 45 |
| 0.7 (0.60-0.83) | 45 | 94.6 | 15 | NA | 130.5 | 60<sup>d</sup> |
| 0.6 (0.50-0.73) | 60 | 185.7 | 20 | NA | NA | NA |
| 0.5 (0.41-0.63) | 75 | 226.3 | 45 | NA | NA | NA |
| 0.4 (--) | NA | NA | NA | NA | NA | NA |
| LTPA | Sleep | PA | Nutrition | Sleep | PA | Nutrition |
| 0.9 (0.86-0.95) | 15 | 5.2 | 5 | 72<sup>c</sup> | 6.6 | 29<sup>#</sup> |
| 0.8 (0.73-0.88) | 30 | 11.4 | 10 | NA | 14.4 | 45 |
| 0.7 (0.62-0.81) | 45 | 18.8 | 15 | NA | 23.8 | 60<sup>d</sup> |
| 0.6 (0.51-0.72) | 60 | 28.3 | 20 | NA | 37.5 | NA |
| 0.5 (0.42-0.61) | 75 | 45.1 | 25 | NA | 66.5<sup>e</sup> | NA |
| 0.4 (0.32-0.51) | 90 | 57.5 | 50 | NA | NA | NA |
| TPA | Sleep | PA | Nutrition | Sleep | PA | Nutrition |
| 0.9 (0.82-1.00) | 30 | 5.5 | 5 | 72<sup>c</sup> | 10.8<sup>#</sup> | 29<sup>#</sup> |
| 0.8 (0.66-0.98) | 45 | 10.2 | 25 | NA | 21.8<sup>f</sup> | 45 |
| 0.7 (0.57-0.88) | 60 | 12.6 | 40 | NA | NA | 60<sup>d</sup> |
| 0.6 (0.45-0.81) | 135 | 18.9 | 60 | NA | NA | NA |
| 0.5 (--) | NA | NA | NA | NA | NA | NA |
| 0.4 (--) | NA | NA | NA | NA | NA | NA |

a: HR = 0.51 (0.45-0.59); b: HR = 0.64 (0.51-0.81); c: HR = 0.91 (0.87-0.97); d: HR = 0.71 (0.57-0.90); e: HR = 0.53 (0.45-0.64); f: HR = 0.86 (0.75-1.00); #: Statistically not significant. NA: not applicable; PA: physical activity; MVPA: moderate-to-vigorous
physical activity; OPA: occupational physical activity; LTPA: leisure-time physical activity; TPA: transportational physical activity.

### Sensitivity analysis

We repeated the main analysis by additionally adjusting for BMI and results still showed a similar pattern (n = 31,875; events = 2,623, **Supplementary Figure 5-6**). Excluding participants with low BMI (BMI <18.5 kg/m², n = 31,308; events = 2,524), previous CVD or cancer (n = 26,374; events = 1,328), implausible total energy intake (n = 30,600; events = 2,511), current smokers (n = 25,553; events = 2,088), poor health status (n = 29,107; events = 2,267), and without two day diet data available (n = 27,787; events = 2,323) did not materially influence the results (**Supplementary Figure 7-19**). We further repeated the main analyses using NHANES day 1 dietary recall weights (n = 31,875; events = 2,623) and the findings were generally consistent with the main unweighted analyses (**Supplementary Figures 18–23**). Replacing HEI-2020 with the AHEI did not materially change the findings (**Supplementary Figure 24-25**). In our subgroup analyses, we still found generally lower all-cause mortality for combinations of optimal sleep, high MVPA and moderate to high diet quality across different sex, ethnicity, education and PIR groups (**Supplementary Figure 27-30**). Besides, we did not observe materially change in the complete sample, primary sample with 5 times of multiple imputation, and 10 times of multiple imputations (**Supplementary Figure 34-37**).

## Discussion

In this study of US adults, we examined combined sleep duration, physical activity, and diet quality in association with all-cause mortality, with consideration of physical activity domains. We found that an additional 15 min/day of sleep, 10.8 min/day of MVPA, and a 5-point HEI-2020 score was associated with a 10% lower risk of all-cause mortality. Minimum variation patterns in leisure-time physical activity showed the strongest associations with lower all-cause mortality, followed by occupational and transportational physical activity. We also found that combinations of optimal sleep (7-8 h/day), moderate and above diet quality (HEI-2020: > 45.1) and high physical activity were generally associated with the lowest all-cause mortality, although there were slight differences across physical activity domains.

Our findings are broadly consistent with the previous UK Biobank analysis of 59,078 participants, in which accelerometer-derived measures showed that sleep of 7.2– 8.0h/day, higher MVPA, and higher diet quality were associated with a 64% lower all-cause mortality risk(16). In both cohorts, MVPA showed the strongest gradient in association with all-cause mortality, likely reflecting its direct physiological pathways to cardiorespiratory fitness, metabolic health, and systemic inflammation(62–64). However, we noticed larger minimum MVPA variations (e.g., 11 min/day vs 1.5 min/day) in our self-reported sample. It is potentially due to overreporting in questionnaires and the GPAQ requirement to report only activity lasting at least 10 continuous minutes(65, 66). We also observed a slightly weaker SPAN-mortality association, which may reflect differences in exposure measurement (HEI-2020 vs dietary quality score) or cohort composition. Compared to the UK Biobank, our participants were younger (48.0 vs 64.0), with a higher median BMI (28.0 vs 26.1) and more current smokers (19.8% vs 6.3%), consistent with a higher burden of adverse lifestyle risk factors in the US population(38, 39, 67). Beyond these differences, we further observed a homogeneity in the SPAN-mortality associations across demographic and socioeconomic strata. Combinations of 7–8 h/day of sleep, high total MVPA (> 72.9 min/day), and at least moderate diet quality (HEI-2020: >45.0) were generally associated with lower all-cause mortality. The findings were robust in supplementary analyses that applied sample weights and used WHO guideline-based MVPA categorisation. Lower mortality associated with optimal SPAN combination may generalise broadly in US adults rather than being confined to a single demographic or socioeconomic subgroup, although smaller sample sizes within specific strata need cautious interpretation.

To our knowledge, this is the first study to examine PA accumulation pattern by different domains within combined SPAN behaviours in relation to all-cause mortality risk. Leisure-time physical activity showed the most favourable profile, followed by occupational and transportational physical activity. LTPA was associated with up to 60% lower all-cause mortality, compared with 50% for OPA and 40% for transportational physical activity, when combined with concurrent variation in sleep duration and diet quality. With fixed concurrent variations of sleep (15 min/day) and diet (5 points of HEI2020, e.g., equivalent to approximately 1.5 cups of fruit in a typical 2,000-kcal/day diet among individuals with no fruit intake), the minimum additional 5.0 min/day of LTPA was associated with 10% lower all-cause mortality, compared with 24.0 min/day for OPA. These patterns are consistent with previous evidence showing that higher LTPA was associated with lower all-cause mortality, whereas higher OPA may show weaker or even adverse associations(31–33, 68). LTPA tends to be short in duration and is typically undertaken under self-regulated conditions that allow pacing and adequate recovery(68, 69). OPA is often performed under prolonged workload, limited recovery, and repetitive physical strain, which may not confer the same health benefits as LTPA(69). In exploratory analyses, we further observed that among adults who are physically active at work, over half an hour of LTPA (>35.4 minutes) per day was still associated with a lower all-cause mortality when combined with recommended sleep duration (7-8 h/day) and high diet quality. These may therefore suggest that OPA and LTPA each have distinct health benefits and should not be viewed as competing or interchangeable components within a daily SPAN combination.

In contrast, transportational physical activity required larger concurrent variation in sleep duration and diet quality to correspond to comparable theoretical lower all-cause mortality (e.g., 10% lower). Heterogeneity in both transportation modes and intensity composition may explain the situation. In NHANES, walking and cycling were combined in the transportational physical activity questionnaire, meaning that transportational physical activity in our study largely reflected active commuting as a heterogeneous exposure. Consistent with this, a previous UK Biobank study including 263,450 participants observed that commuting by bicycle, but not by foot, was associated with lower all-cause mortality(35). Additionally, due to the questionnaire, vigorous intensity of physical activity from transport in NHANES cannot be ascertained, limiting our analysis of TPA to MPA. As VPA may show stronger mortality associations than MPA(70), its absence may partly explain the less favourable profile in transportational domain. Despite this heterogeneity across PA domains, our study still highlighted that engaging in even a small amount of physical activity in any domain may be more beneficial than remaining inactive(71), particularly when sleep duration and diet quality are theoretically improved simultaneously. Beyond total MVPA, the domain in which activity is accumulated, and behavioural choice (e.g., walking or cycling) may also matter when interpreting physical activity within the SPAN combination and its association with mortality.

In our study, we also found that associations of higher PA and better diet quality with lower all-cause mortality were attenuated among participants with long sleep duration (>8.5 h/day). Recent consortia evidence from population level cohorts suggests that long sleep duration (>8 h/day) was associated with higher all-cause mortality and with biological ageing across multiple organ systems(72). Prolonged sleep may therefore partly reflect underlying subclinical disease burdens and accelerated biological ageing, rather than being a simple adverse behaviour in itself. In this context, inflammation may be a plausible explanation, as evidence indicates that prolonged sleep duration is associated with higher levels of inflammatory biomarkers such as interleukin-6 (73, 74). However, this mechanistic interpretation remains speculative. Prolonged sleep may also relate to fatigue or undiagnosed mental illness, and therefore, the observed association should be interpreted cautiously, particularly given the possibility of residual confounding and reverse causation(11, 72).

### Strengths and Limitations

Our study had several strengths. First, we used self-reported SPAN behaviours to assess all three behaviours concurrently in a sample of 31,875 US adults. Although self-report may introduce recall bias and misclassification(75–77), this approach provides measurement-mode consistency across SPAN behaviours and allows us to examine physical activity domains that cannot be captured by accelerometers(43, 78). Second, we were, to our knowledge, the first to incorporate physical activity domains within the SPAN combination and examine their differences in associations with all-cause mortality. Third, we conducted multifaceted sensitivity analyses, including replacing HEI-2020 with AHEI, to ensure the robustness of our results. Unlike previous SPAN studies (15–17) with a measurement lag of 3-9 years between wearable and dietary data collection, the NHANES study collected sleep duration, physical activity, and dietary intake simultaneously. This approach reduces potential exposure misclassification arising from temporal mismatch between SPAN behaviour assessments.

We acknowledge some limitations of the study. First, because physical activity was highly right-skewed, once we categorised physical activity in each domain into three groups, it resulted in smaller numbers of participants in some categories, particularly the moderate and high groups, which may have reduced statistical power. Second, in NHANES, TPA only captures MPA and may therefore underestimate its potential association with lower all-cause mortality risk. In addition, the relatively small number of participants in the prolonged sleep-duration group may have reduced the stability of these estimates. Moreover, our SPAN sleep component did not capture sleep disorders, such as obstructive sleep apnoea and insomnia, both of which are associated with mortality risk (79). Lastly, sleep duration was self-reported and differed in measurement across NHANES cycles. Cycles from 2007 to 2014 directly assessed usual weekday/workday sleep duration, whereas 2015-2016 and 2017-2018 derived sleep hours from reported sleep and wake times during the main sleep period. The latter may reflect sleep period rather than actual sleep duration, especially among individuals with long sleep latency or wake after sleep onset. This may have introduced measurement heterogeneity and misclassification of sleep categories. Therefore, future studies should also consider using harmonised sleep measures in SPAN studies.

## Conclusions

Modest combined increments in sleep, physical activity, and nutrition were associated with lower all-cause mortality among US adults across physical activity domains, with the strongest association observed for LTPA, followed by OPA and TPA. 7–8 h/day of sleep, moderate to high physical activity depending on physical activity domains, and at least moderate diet quality were generally associated with lower all-cause mortality. Small increments in physical activity in any domain have potential health value in relation to mortality risk, particularly when accompanied by concurrent improvements in sleep duration and diet quality.

## Data Availability

All data produced are available online at the National Health and Nutrition Examination Survey (NHANES).

## Funding

MPP is funded by an NHMRC Investigator Grant (GTN2009264).

## Data Availability

NHANES data are publicly available through the National Center for Health Statistics, Centers for Disease Control and Prevention. Public-use linked mortality data are also available through the National Center for Health Statistics. The derived analytic dataset and statistical code may be made available from the corresponding author upon reasonable request, subject to applicable data-use requirements.

## Notes

### Competing Interest Statement

The authors have declared no competing interest.

### Author Declarations

The study used ONLY openly available human data that were originally located at: the National Health and Nutrition Examination Survey (NHANES).

