## Supplementary files for "Combined sleep, domain-specific physical activity, and nutrition in relation to all-cause mortality among US adults"

**Table of Supplements**


| Pages | Items |
|---|---|
| 3 | **Supplementary Figure 1** Participants flow chart |
| 4 | **Supplementary Figure 2** Dose-response associations of each individual exposure of A) sleep duration, B) moderate-to-vigorous physical activity (MVPA) and C) nutrition (HEI-2020) with all-cause mortality risk |
| 5-6 | **Supplementary Figure 3** Dose-response associations of each individual physical activity domain of A) occupational physical activity (OPA) B) leisure-time physical activity (LTPA) and C) transportational physical activity (TPA) with all-cause mortality risk |
| 7 | **Supplementary Figure 4** Multivariable-adjusted associations of combined sleep, moderate-to-vigorous physical activity (MVPA) and nutrition with all-cause mortality risk, adjusted for BMI |
| 8-9 | **Supplementary Figure 5** Multivariable-adjusted associations of combined sleep, physical activity domains and nutrition with all-cause mortality risk, adjusted for BMI |
| 10 | **Supplementary Figure 6** Multivariable-adjusted associations of combined sleep, moderate-to-vigorous physical activity (MVPA) and nutrition with all-cause mortality risk, excluding underweight participants |
| 11-12 | **Supplementary Figure 7** Multivariable-adjusted associations of combined sleep, physical activity domains and nutrition with all-cause mortality risk, excluding underweight participants |
| 13 | **Supplementary Figure 8** Multivariable-adjusted associations of of combined sleep, moderate-to-vigorous physical activity (MVPA) and nutrition with all-cause mortality risk, excluding participants with only one day dietary recall available |
| 14-15 | **Supplementary Figure 9** Multivariable-adjusted associations of combined sleep, physical activity domains and nutrition with all-cause mortality risk, excluding participants with only one day dietary recall available |
| 16 | **Supplementary Figure 10** Multivariable-adjusted associations of combined sleep, moderate-to-vigorous physical activity (MVPA) and nutrition with all-cause mortality risk, excluding participants with previous diagnosed cardiovascular diseases or cancer |
| 17-18 | **Supplementary Figure 11** Multivariable-adjusted associations of combined sleep, physical activity domains and nutrition with all-cause mortality risk, excluding participants with previous diagnosed cardiovascular diseases or cancer |
| 19 | **Supplementary Figure 12** Multivariable-adjusted associations of combined sleep, moderate-to-vigorous physical activity (MVPA) and nutrition with all-cause mortality risk, excluding participants with implausible daily total energy intake |
| 20-21 | **Supplementary Figure 13** Multivariable-adjusted associations of combined sleep, physical activity domains and nutrition with all-cause mortality risk, excluding participants with implausible daily total energy intake |
| 22 | **Supplementary Figure 14** Multivariable-adjusted associations of combined sleep, moderate-to-vigorous physical activity (MVPA) and nutrition with all-cause mortality risk, excluding current smokers |
| 23-24 | **Supplementary Figure 15** Multivariable-adjusted associations of combined sleep, physical activity domains and nutrition with all-cause mortality risk, excluding current smokers |
| 25 | **Supplementary Figure 16** Multivariable-adjusted associations of combined sleep, moderate-to-vigorous physical activity (MVPA) and nutrition with all-cause mortality risk, excluding participants with self-reported poor health status |
| 26-27 | **Supplementary Figure 17** Multivariable-adjusted associations of combined sleep, physical activity domains and nutrition with all-cause mortality risk, excluding participants with current poor health status |
| 28 | **Supplementary Figure 18** Multivariable-adjusted associations of combined sleep, moderate-to-vigorous physical activity (MVPA) and nutrition with all-cause mortality risk, using day-1 sample weights |
| 29-30 | **Supplementary Figure 19** Multivariable-adjusted associations of combined sleep, physical activity domains and nutrition with all-cause mortality risk, using day-1 sample weights |
| 31 | **Supplementary Figure 20** Multivariable-adjusted associations of combined sleep, moderate-to-vigorous physical activity (MVPA) and nutrition with all-cause mortality risk, using Alternative Healthy Eating Index-2010 |
| 32-33 | **Supplementary Figure 21** Multivariable-adjusted associations of combined sleep, physical activity domains and nutrition with all-cause mortality risk, using Alternative Healthy Eating Index-2010 |

| | |
|---|---|
| 34 | **Supplementary Figure 22** Multivariable-adjusted associations of combined sleep, leisure-time physical activity (LTPA) and Nutrition with all-cause mortality risk, excluding participants with no occupational physical activity |
| 35 | **Supplementary Figure 23** Multivariable-adjusted associations of combined sleep, moderate-to-vigorous physical activity (MVPA) and nutrition with all-cause mortality risk across sex groups |
| 36 | **Supplementary Figure 24** Multivariable-adjusted associations of combined sleep, moderate-to-vigorous physical activity (MVPA) and nutrition with all-cause mortality risk across different ethnic groups |
| 37 | **Supplementary Figure 25** Multivariable-adjusted associations of combined sleep, moderate-to-vigorous physical activity (MVPA) and nutrition with all-cause mortality risk across different educational levels |
| 38 | **Supplementary Figure 26** Multivariable-adjusted associations of combined sleep, moderate-to-vigorous physical activity (MVPA) and nutrition with all-cause mortality risk across different Poverty income ratio (PIR) groups |
| 39 | **Supplementary Figure 27** Multivariable-adjusted associations of combined sleep, WHO-guideline based moderate-to-vigorous physical activity (MVPA) groups and nutrition with all-cause mortality risk |
| 40 | **Supplementary Figure 28** Multivariable-adjusted associations of combined sleep, moderate-to-vigorous physical activity (MVPA) and nutrition with all-cause mortality risk, not adjusting for total energy intake |
| 41-42 | **Supplementary Figure 29** Multivariable-adjusted associations of combined sleep, physical activity domains and nutrition with all-cause mortality risk, not adjusting for total energy intake |
| 43 | **Supplementary Figure 30** Multivariable-adjusted associations of combined sleep, moderate-to-vigorous physical activity (MVPA) and nutrition with all-cause mortality risk, using complete data |
| 44-45 | **Supplementary Figure 31** Multivariable-adjusted associations of combined sleep, physical activity domains and nutrition with all-cause mortality risk, using complete data |
| 46-47 | **Supplementary Figure 32** Multivariable-adjusted associations of combined sleep, moderate-to-vigorous physical activity (MVPA) and nutrition with all-cause mortality risk, using multiple imputation for 5 times (primary) and 10 times (MI10) |
| 48-49 | **Supplementary Figure 33** Multivariable-adjusted associations of combined sleep, physical activity domains and nutrition with all-cause mortality risk, not adjusting for total energy intake |
| 50-51 | **Supplementary Table 1** STROBE statement |
| 52 | **Supplementary Table 2** Variables extracted from NHANES |
| 53 | **Supplementary Table 3** Components of Health Eating Index-2020 and standards of scoring |
| 54-55 | **Supplementary Table 4** Sample characteristics of individuals in the primary analysis sample using multiple imputations versus individuals with incomplete or missing data. |

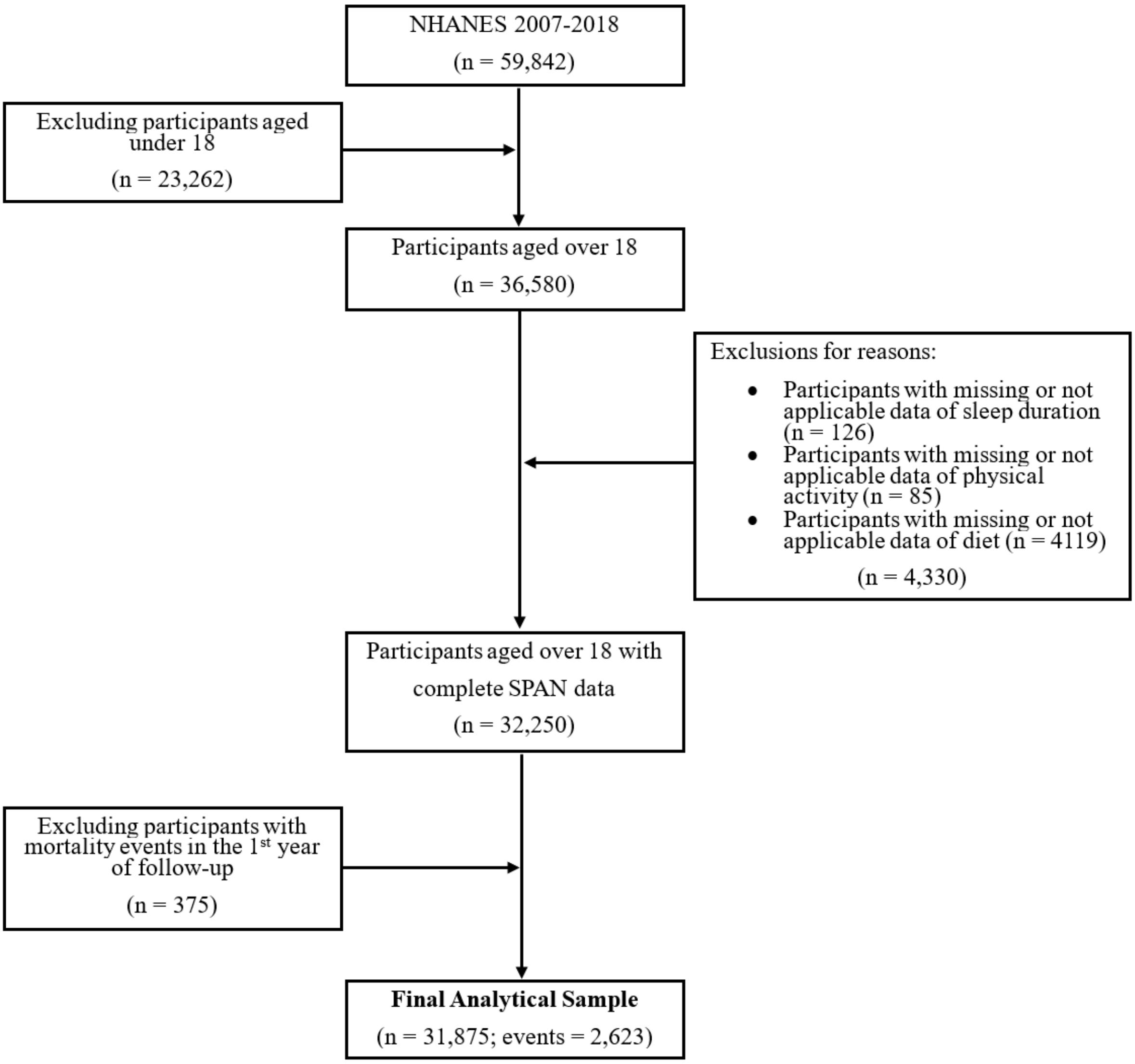


**Supplementary Figure 1 Participants flow chart**

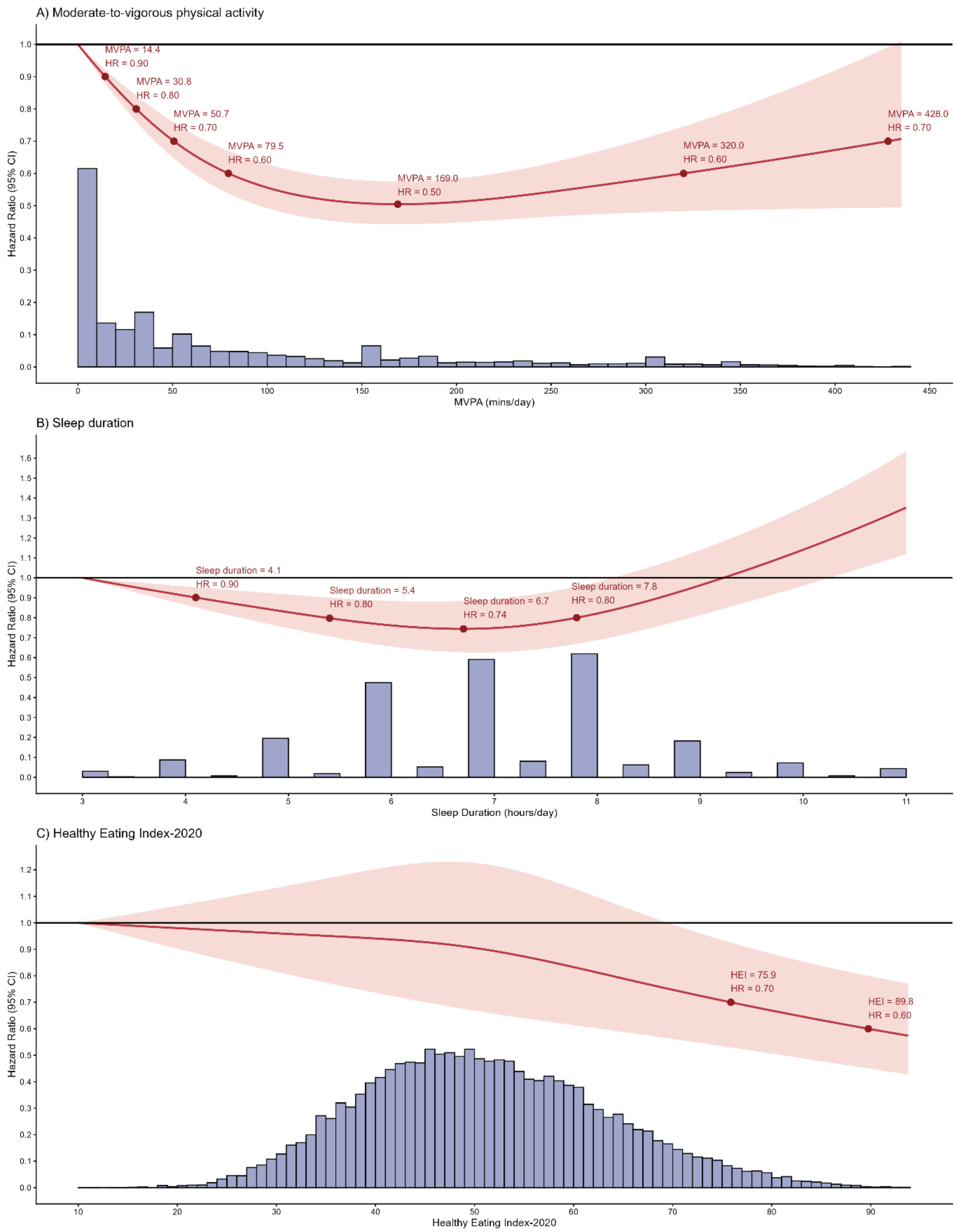


**Supplementary Figure 2 Dose-response associations of each individual exposure of A) moderate-to-vigorous physical activity (MVPA), B) sleep duration, and C) nutrition (HEI-2020) with all-cause mortality risk (n = 31,875; events = 2,623)**. Legend: referent point was set up as the lowest value of each exposure. Red points denote the value of each exposure associating with increments of 10% lower risks or the nearest value. Models were adjusted for age, sex, ethnicity, education, ratio of family income to poverty, alcohol consumption, smoking status, sedentary activity time, total energy intake, previous diagnosis of cardiovascular disease, cancer or type 2 diabetes, and family history of heart attack and diabetes.

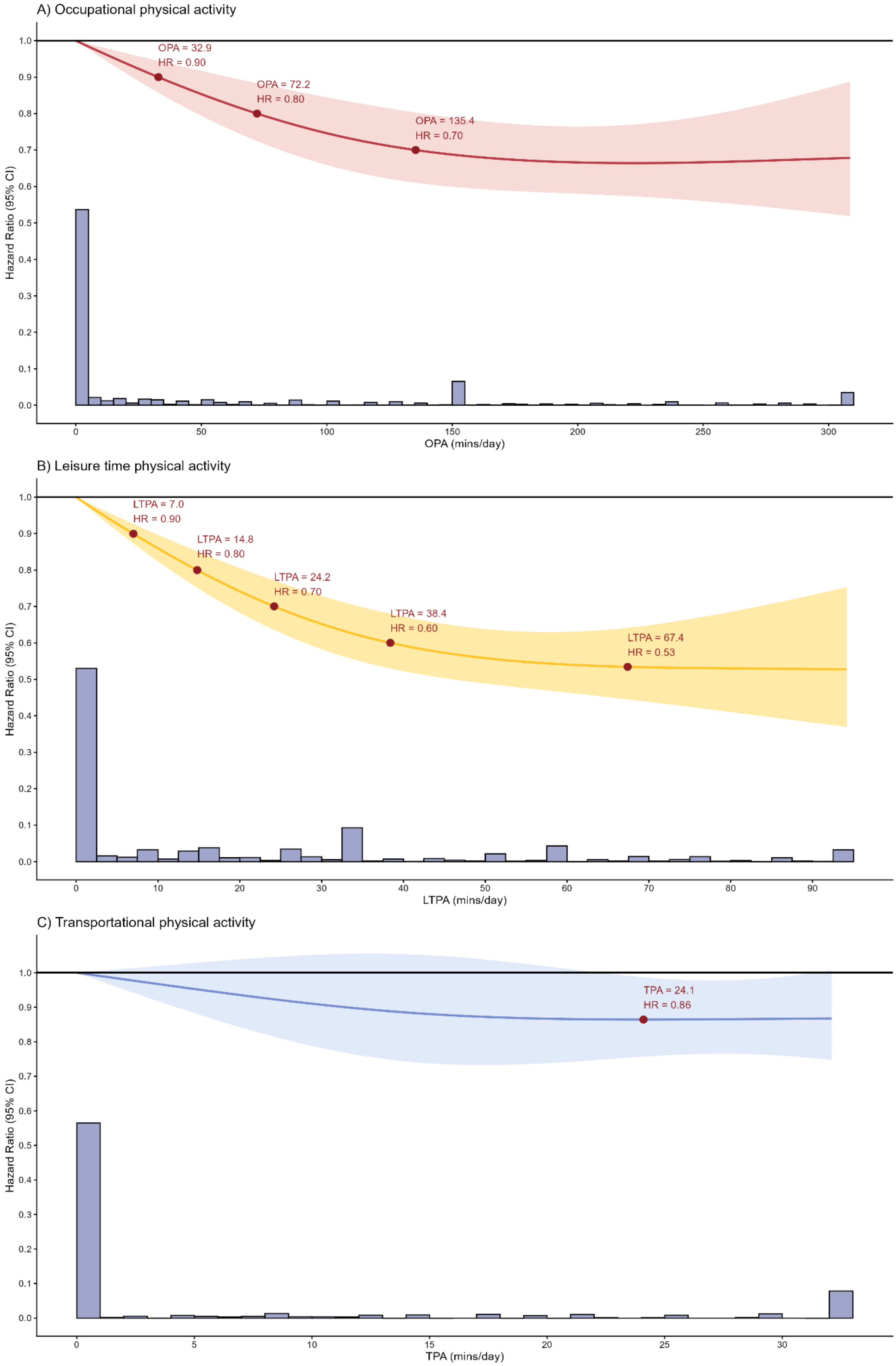


**Supplementary Figure 3 Dose-response associations of each individual physical activity domain of A)**

**occupational physical activity (OPA) B) leisure-time physical activity (LTPA) and C) transportational physical activity (TPA) with all-cause mortality risk (n = 31,875; events = 2,623)**. **Legend**: referent point was set up as the lowest value of each exposure. Red points denote the value of each exposure associating with increments of 10% lower risks or the nearest value. Models were adjusted for age, sex, ethnicity, education, ratio of family income to poverty, alcohol consumption, smoking status, sedentary activity time, total energy intake, previous diagnosis of cardiovascular disease, cancer or type 2 diabetes, and family history of heart attack and diabetes. When examining one domain as the primary exposure, the other two physical activity domains were adjusted in the multivariable models.

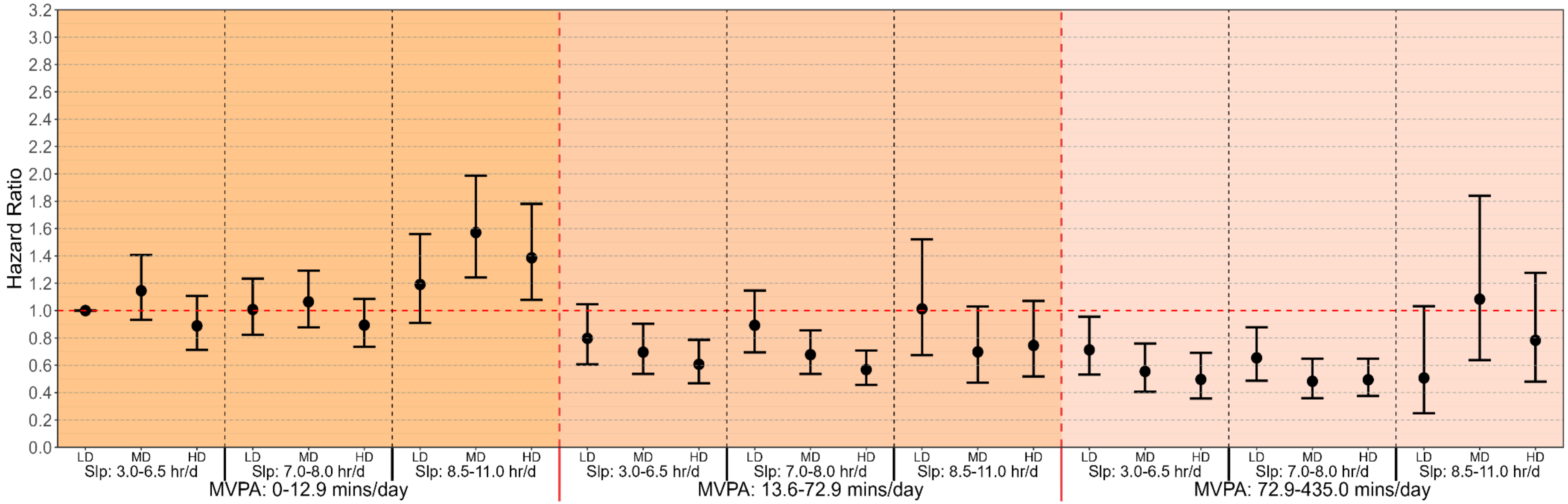


**Supplementary Figure 4 Multivariable-adjusted associations of combined sleep, moderate-to-vigorous physical activity (MVPA) and nutrition with all-cause mortality risk, adjusted for BMI (n = 31,875; events = 2,623). Legend:** Model is adjusted for age, sex, ethnicity, education, body mass index (BMI), ratio of family income to poverty, alcohol consumption, smoking status, sedentary activity time, total energy intake, previous diagnosis of cardiovascular disease, cancer or type 2 diabetes, and family history of heart attack and diabetes. Sleep duration (hours/day), MVPA (minutes/day), and nutrition (Healthy Eating Index-2020 (HEI2020)) were treated as a joint term. The specific groups for each exposure were divided as sleep duration: 3.0-6.5 hours/day (short), 7.0-8.0 hours/day (optimal) and 8.5-11.0 hours/day (long); MVPA: 0-12.9 minutes/day (low), 13.6-72.9 minutes/day (moderate) and 72.9-435.0 minutes/day (high); HEI-2020: 10.0-45.0 (low), 45.0-56.0 (moderate), and 56.0-93.8 (high). The lowest group for all three exposures were treated as reference group. LD: low diet quality; MD: moderate diet quality; HD: high diet quality; Slp: sleep duration.

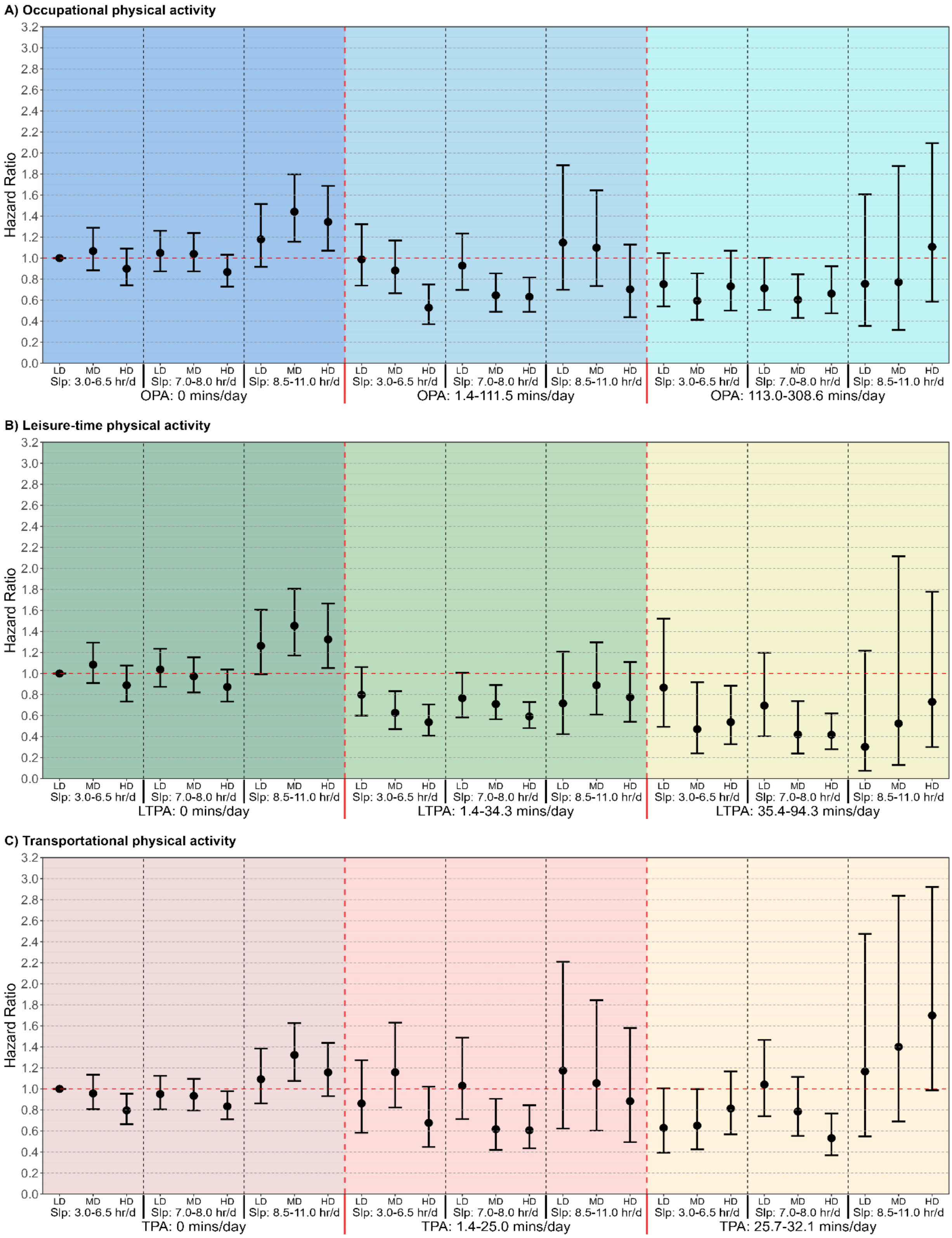


**Supplementary Figure 5 Multivariable-adjusted associations of combined sleep, physical activity domains and nutrition with all-cause mortality risk, adjusted for BMI (n = 31,875; events = 2,623). Legend:** A: Occupational physical activity (OPA); B: Leisure-time physical activity (LTPA); C: Transportational physical activity (TPA). Models are adjusted for adjusted for age, sex, ethnicity, education, body mass index (BMI), ratio of family income to poverty, alcohol consumption, smoking status, sedentary activity time, total energy intake, previous diagnosis of cardiovascular disease, cancer or type 2 diabetes, and family history of heart attack and diabetes. When examining one PA domain as the primary exposure, the other two physical activity domains were adjusted in the multivariable models. Sleep duration (hours/day), OPA/LTPA/TPA (minutes/day), and nutrition (Healthy Eating Index-2020 (HEI2020)) were treated as a joint term. The specific groups for each exposure were divided as sleep duration: 3.0-6.5 hours/day (short), 7.0-8.0 hours/day (optimal) and 8.5-11.0 hours/day (long); OPA: 0 minutes/day (low), 1.4-111.5 minutes/day (moderate) and 113.0-308.6 minutes/day (high); LTPA: 0 minutes/day (low), 1.4-34.3 minutes/day (moderate) and 35.4-94.3 minutes/day (high); TPA: 0 minutes/day (low), 1.4-25.0 minutes/day (moderate) and 25.7-32.1 minutes/day (high); HEI-2020: 10.0-45.0 (low), 45.0-56.0 (moderate), and 56.0-93.8 (high). The lowest group for all three exposures in each physical activity domain were treated as reference group. LD: low diet quality; MD: moderate diet quality; HD: high diet quality; Slp: sleep duration.

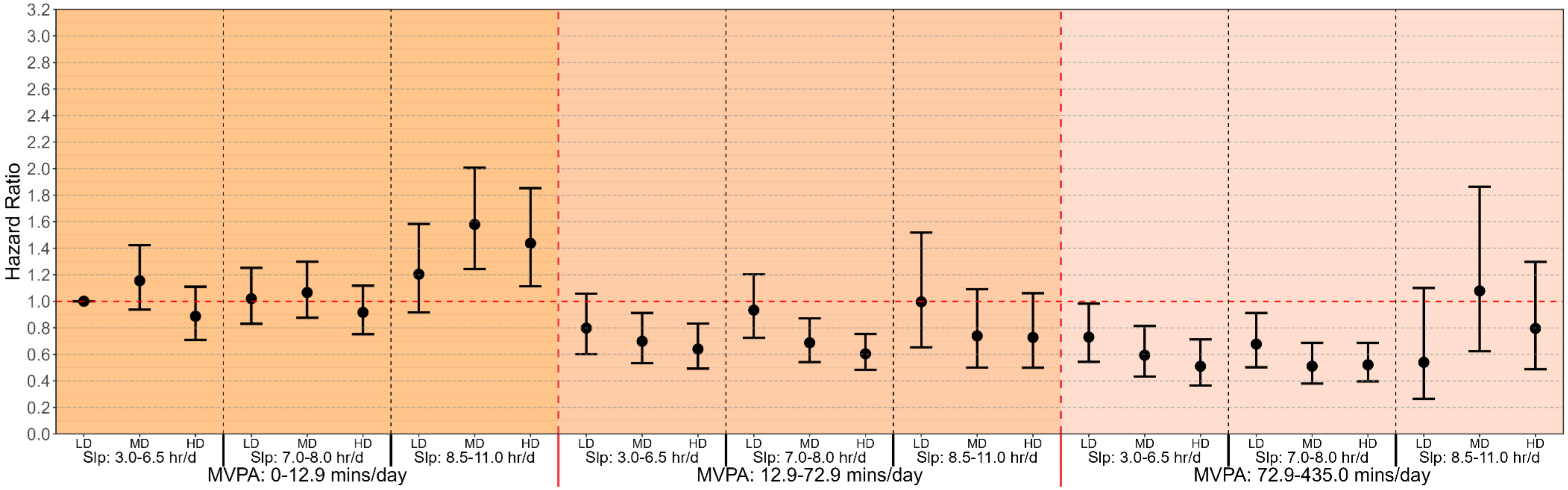


**Supplementary Figure 6 Multivariable-adjusted associations of combined sleep, moderate-to-vigorous physical activity (MVPA) and nutrition with all-cause mortality risk, excluding underweight participants (BMI <18.5 kg/m²) (n = 31,308; events = 2,524). Legend:** Model is adjusted for age, sex, ethnicity, education, ratio of family income to poverty, alcohol consumption, smoking status, sedentary activity time, total energy intake, previous diagnosis of cardiovascular disease, cancer or type 2 diabetes, and family history of heart attack and diabetes. Sleep duration (hours/day), MVPA (minutes/day), and nutrition (Healthy Eating Index-2020 (HEI2020)) were treated as a joint term. The specific groups for each exposure were divided as sleep duration: 3.0-6.5 hours/day (short), 7.0-8.0 hours/day (optimal) and 8.5-11.0 hours/day (long); MVPA: 0-12.9 minutes/day (low), 12.9-72.9 minutes/day (moderate) and 72.9-435.0 minutes/day (high); HEI-2020: 10.0-45.0 (low), 45.0-56.0 (moderate), and 56.0-93.8 (high). The lowest group for all three exposures were treated as reference group. LD: low diet quality; MD: moderate diet quality; HD: high diet quality; Slp: sleep duration.

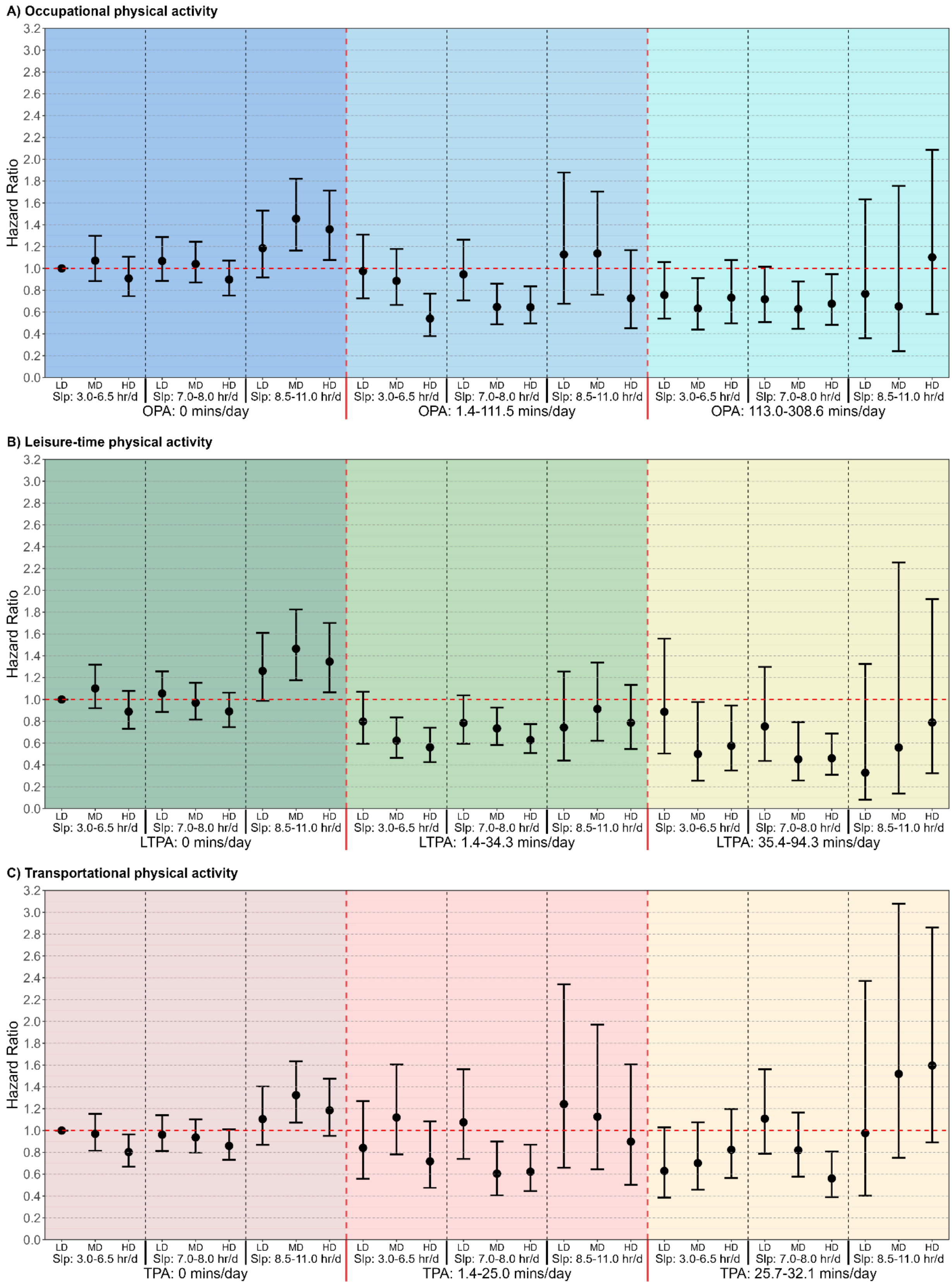


**Supplementary Figure 7 Multivariable-adjusted associations of combined sleep, physical activity domains and nutrition with all-cause mortality risk, excluding underweight participants (BMI <18.5 kg/m²) (n = 31,308; events = 2,524). Legend:** A: Occupational physical activity (OPA); B: Leisure-time physical activity (LTPA); C: Transportational physical activity (TPA). Models are adjusted for adjusted for age, sex, ethnicity, education, ratio of family income to poverty, alcohol consumption, smoking status, sedentary activity time, total energy intake, previous diagnosis of cardiovascular disease, cancer or type 2 diabetes, and family history of heart attack and diabetes. When examining one PA domain as the primary exposure, the other two physical activity domains were adjusted in the multivariable models. Sleep duration (hours/day), OPA/LTPA/TPA (minutes/day), and nutrition (Healthy Eating Index-2020 (HEI2020)) were treated as a joint term. The specific groups for each exposure were divided as sleep duration: 3.0-6.5 hours/day (short), 7.0-8.0 hours/day (optimal) and 8.5-11.0 hours/day (long); OPA: 0 minutes/day (low), 1.4-111.5 minutes/day (moderate) and 113.0-308.6 minutes/day (high); LTPA: 0 minutes/day (low), 1.4-34.3 minutes/day (moderate) and 35.4-94.3 minutes/day (high); TPA: 0 minutes/day (low), 1.4-25.0 minutes/day (moderate) and 25.7-32.1 minutes/day (high); HEI-2020: 10.0-45.0 (low), 45.0-56.0 (moderate), and 56.0-93.8 (high). The lowest group for all three exposures in each physical activity domain were treated as reference group. LD: low diet quality; MD: moderate diet quality; HD: high diet quality; Slp: sleep duration.

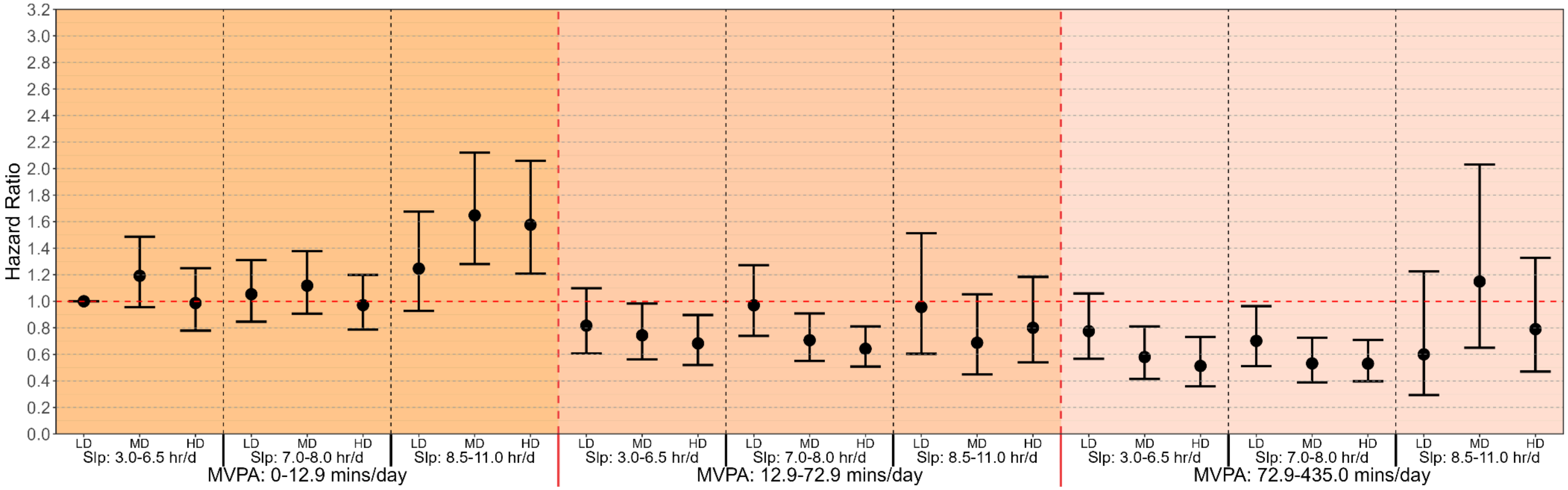


**Supplementary Figure 8 Multivariable-adjusted associations of combined sleep, moderate-to-vigorous physical activity (MVPA) and nutrition with all-cause mortality risk, excluding participants with only one day dietary recall available (n = 27,787; events = 2,323). Legend:** Model is adjusted for age, sex, ethnicity, education, ratio of family income to poverty, alcohol consumption, smoking status, sedentary activity time, total energy intake, previous diagnosis of cardiovascular disease, cancer or type 2 diabetes, and family history of heart attack and diabetes. Sleep duration (hours/day), MVPA (minutes/day), and nutrition (Healthy Eating Index-2020 (HEI2020)) were treated as a joint term. The specific groups for each exposure were divided as sleep duration: 3.0-6.5 hours/day (short), 7.0-8.0 hours/day (optimal) and 8.5-11.0 hours/day (long); MVPA: 0-12.9 minutes/day (low), 12.9-72.9 minutes/day (moderate) and 72.9-435.0 minutes/day (high); HEI-2020: 10.0-45.3 (low), 45.3-56.2 (moderate), and 56.2-93.8 (high). The lowest group for all three exposures were treated as reference group. LD: low diet quality; MD: moderate diet quality; HD: high diet quality; Slp: sleep duration.

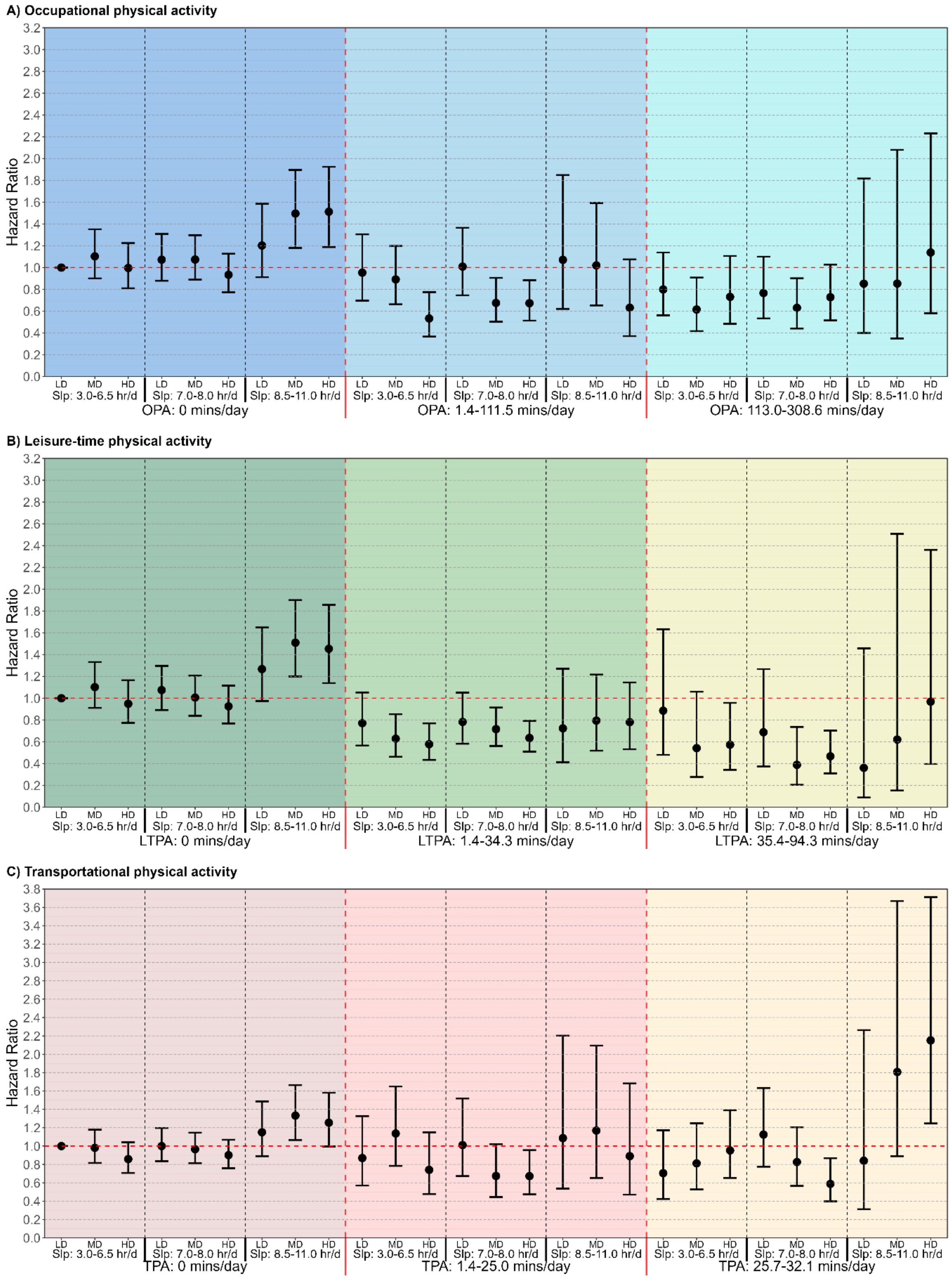


**Supplementary Figure 9 Multivariable-adjusted associations of combined sleep, physical activity domains and nutrition with all-cause mortality risk, excluding participants with only one day dietary recall available (n = 27,787; events = 2,323). Legend:** A: Occupational physical activity (OPA); B: Leisure-time physical activity (LTPA); C: Transportational physical activity (TPA). Models are adjusted for adjusted for age, sex, ethnicity, education, ratio of family income to poverty, alcohol consumption, smoking status, sedentary activity time, total energy intake, previous diagnosis of cardiovascular disease, cancer or type 2 diabetes, and family history of heart attack and diabetes. When examining one PA domain as the primary exposure, the other two physical activity domains were adjusted in the multivariable models. Sleep duration (hours/day), OPA/LTPA/TPA (minutes/day), and nutrition (Healthy Eating Index-2020 (HEI2020)) were treated as a joint term. The specific groups for each exposure were divided as sleep duration: 3.0-6.5 hours/day (short), 7.0-8.0 hours/day (optimal) and 8.5-11.0 hours/day (long); OPA: 0 minutes/day (low), 1.4-111.5 minutes/day (moderate) and 113.0-308.6 minutes/day (high); LTPA: 0 minutes/day (low), 1.4-34.3 minutes/day (moderate) and 35.4-94.3 minutes/day (high); TPA: 0 minutes/day (low), 1.4-25.0 minutes/day (moderate) and 25.7-32.1 minutes/day (high); HEI-2020: 10.0-45.3 (low), 45.3-56.2 (moderate), and 56.2-93.8 (high). The lowest group for all three exposures in each physical activity domain were treated as reference group. LD: low diet quality; MD: moderate diet quality; HD: high diet quality; Slp: sleep duration.

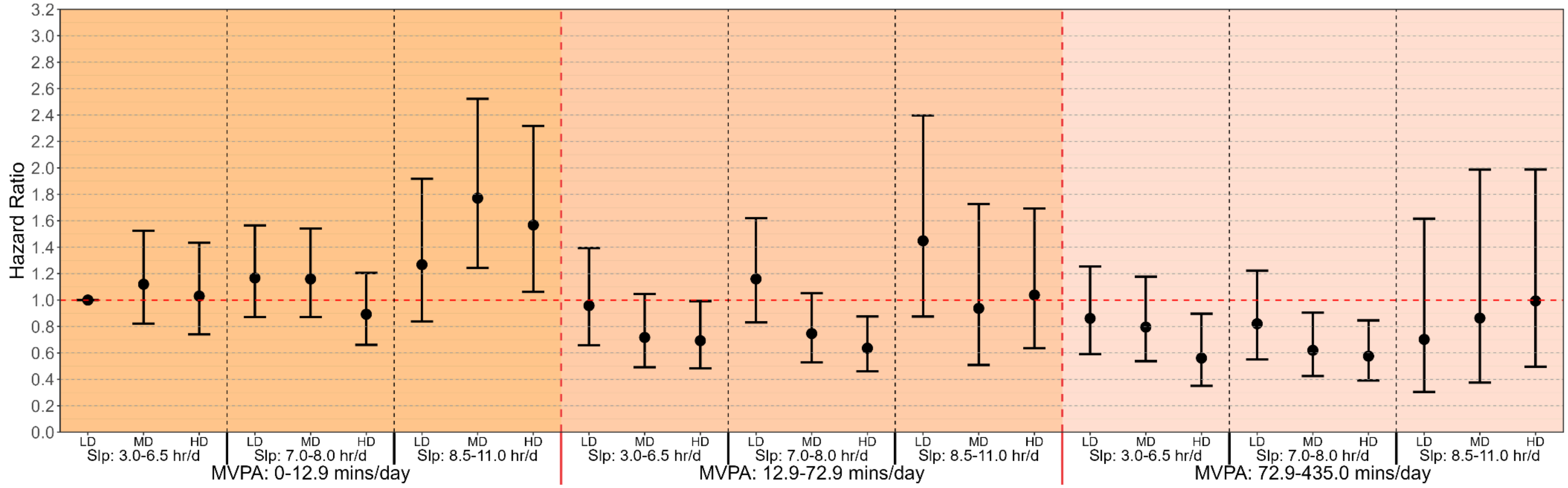


**Supplementary Figure 10 Multivariable-adjusted associations of combined sleep, moderate-to-vigorous physical activity (MVPA) and nutrition with all-cause mortality risk, excluding participants with previous diagnosed cardiovascular diseases or cancer (n = 26,374; events = 1,328). Legend:** Model is adjusted for age, sex, ethnicity, education, ratio of family income to poverty, alcohol consumption, smoking status, sedentary activity time, total energy intake, previous diagnosis of type 2 diabetes, and family history of heart attack and diabetes. Sleep duration (hours/day), MVPA (minutes/day), and nutrition (Healthy Eating Index-2020 (HEI2020)) were treated as a joint term. The specific groups for each exposure were divided as sleep duration: 3.0-6.5 hours/day (short), 7.0-8.0 hours/day (optimal) and 8.5-11.0 hours/day (long); MVPA: 0-12.9 minutes/day (low), 12.9-72.9 minutes/day (moderate) and 72.9-435.0 minutes/day (high); HEI-2020: 10.0-45.0 (low), 45.0-56.0 (moderate), and 56.0-93.8 (high). The lowest group for all three exposures were treated as reference group. LD: low diet quality; MD: moderate diet quality; HD: high diet quality; Slp: sleep duration.

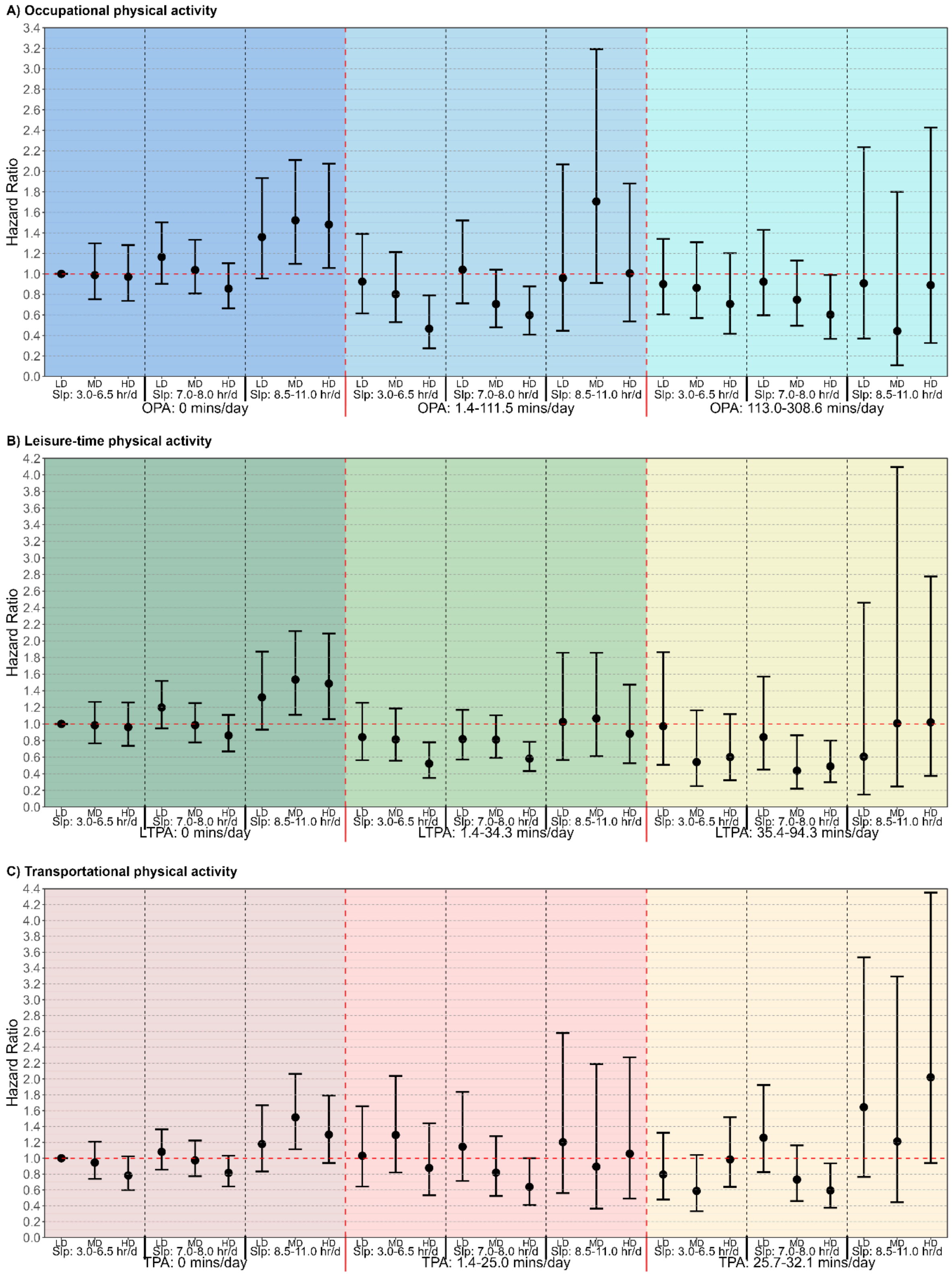


**Supplementary Figure 11 Multivariable-adjusted associations of combined sleep, physical activity domains and nutrition with all-cause mortality risk, excluding participants with previous diagnosed cardiovascular diseases or cancer (n = 26,374; events = 1,328). Legend:** A: Occupational physical activity (OPA); B: Leisure-time physical activity (LTPA); C: Transportational physical activity (TPA). Models are adjusted for adjusted for age, sex, ethnicity, education, ratio of family income to poverty, alcohol consumption, smoking status, sedentary activity time, total energy intake, previous diagnosis of type 2 diabetes, and family history of heart attack and diabetes. When examining one PA domain as the primary exposure, the other two physical activity domains were adjusted in the multivariable models. Sleep duration (hours/day), OPA/LTPA/TPA (minutes/day), and nutrition (Healthy Eating Index-2020 (HEI2020)) were treated as a joint term. The specific groups for each exposure were divided as sleep duration: 3.0-6.5 hours/day (short), 7.0-8.0 hours/day (optimal) and 8.5-11.0 hours/day (long); OPA: 0 minutes/day (low), 1.4-111.5 minutes/day (moderate) and 113.0-308.6 minutes/day (high); LTPA: 0 minutes/day (low), 1.4-34.3 minutes/day (moderate) and 35.4-94.3 minutes/day (high); TPA: 0 minutes/day (low), 1.4-25.0 minutes/day (moderate) and 25.7-32.1 minutes/day (high); HEI-2020: 10.0-45.0 (low), 45.0-56.0 (moderate), and 56.0-93.8 (high). The lowest group for all three exposures in each physical activity domain were treated as reference group. LD: low diet quality; MD: moderate diet quality; HD: high diet quality; Slp: sleep duration.

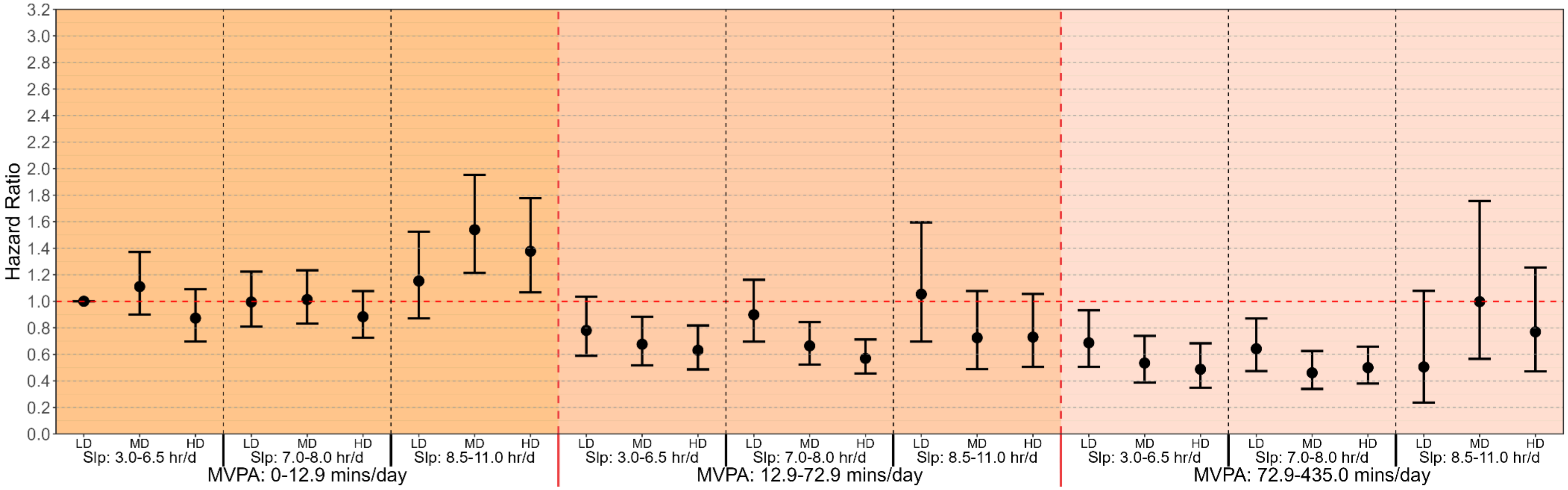


**Supplementary Figure 12 Multivariable-adjusted associations of combined sleep, moderate-to-vigorous physical activity (MVPA) and nutrition with all-cause mortality risk, excluding participants with implausible daily total energy intake [(1)] (n = 30,600; events = 2,511). Legend:** Sex-specific implausible daily energy intakes are applied as <600 or >3500 kcal/day for women; <800 or >4200 kcal/day for men [(1)]. Model is adjusted for age, sex, ethnicity, education, ratio of family income to poverty, alcohol consumption, smoking status, sedentary activity time, total energy intake, previous diagnosis of cardiovascular disease, cancer or type 2 diabetes, and family history of heart attack and diabetes. Sleep duration (hours/day), MVPA (minutes/day), and nutrition (Healthy Eating Index-2020 (HEI2020)) were treated as a joint term. The specific groups for each exposure were divided as sleep duration: 3.0-6.5 hours/day (short), 7.0-8.0 hours/day (optimal) and 8.5-11.0 hours/day (long); MVPA: 0-12.9 minutes/day (low), 12.9-72.9 minutes/day (moderate) and 72.9-435.0 minutes/day (high); HEI-2020: 12.0-45.0 (low), 45.0-56.0 (moderate), and 56.0-93.8 (high). The lowest group for all three exposures were treated as reference group. LD: low diet quality; MD: moderate diet quality; HD: high diet quality; Slp: sleep duration.

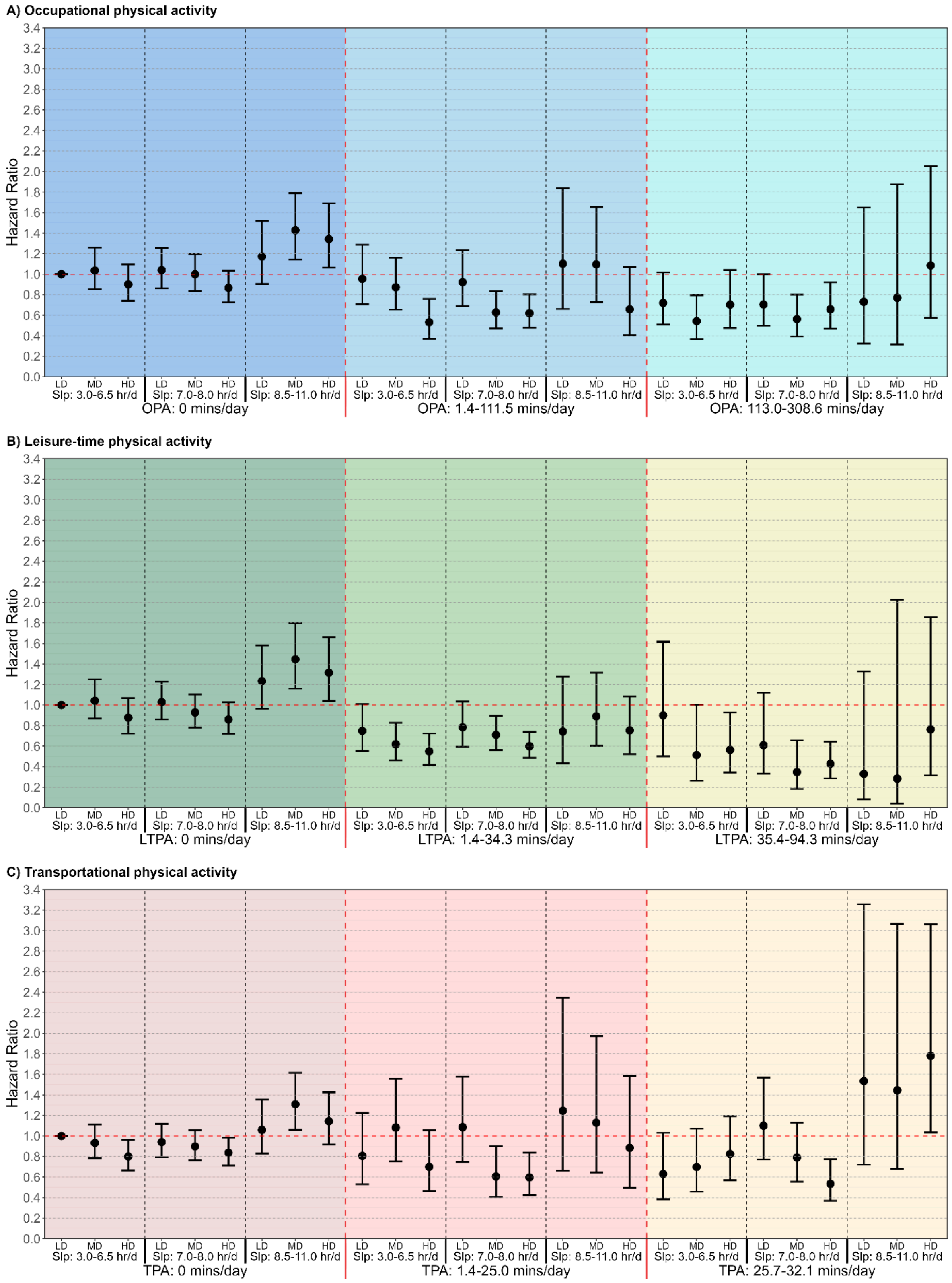


**Supplementary Figure 13 Multivariable-adjusted associations of combined sleep, physical activity domains and nutrition with all-cause mortality risk, excluding participants with implausible daily total energy intake [(1)] (n = 30,600; events = 2,511). Legend:** A: Occupational physical activity (OPA); B: Leisure-time physical activity (LTPA); C: Transportational physical activity (TPA). Sex-specific implausible daily energy intakes are applied as <600 or >3500 kcal/day for women; <800 or >4200 kcal/day for men [(1)]. Models are adjusted for adjusted for age, sex, ethnicity, education, ratio of family income to poverty, alcohol consumption, smoking status, sedentary activity time, total energy intake, previous diagnosis of cardiovascular disease, cancer or type 2 diabetes, and family history of heart attack and diabetes. When examining one PA domain as the primary exposure, the other two physical activity domains were adjusted in the multivariable models. Sleep duration (hours/day), OPA/LTPA/TPA (minutes/day), and nutrition (Healthy Eating Index-2020 (HEI2020)) were treated as a joint term. The specific groups for each exposure were divided as sleep duration: 3.0-6.5 hours/day (short), 7.0-8.0 hours/day (optimal) and 8.5-11.0 hours/day (long); OPA: 0 minutes/day (low), 1.4-111.5 minutes/day (moderate) and 113.0-308.6 minutes/day (high); LTPA: 0 minutes/day (low), 1.4-34.3 minutes/day (moderate) and 35.4-94.3 minutes/day (high); TPA: 0 minutes/day (low), 1.4-25.0 minutes/day (moderate) and 25.7-32.1 minutes/day (high); HEI-2020: 12.0-45.0 (low), 45.0-56.0 (moderate), and 56.0-93.8 (high). The lowest group for all three exposures in each physical activity domain were treated as reference group. LD: low diet quality; MD: moderate diet quality; HD: high diet quality; Slp: sleep duration.

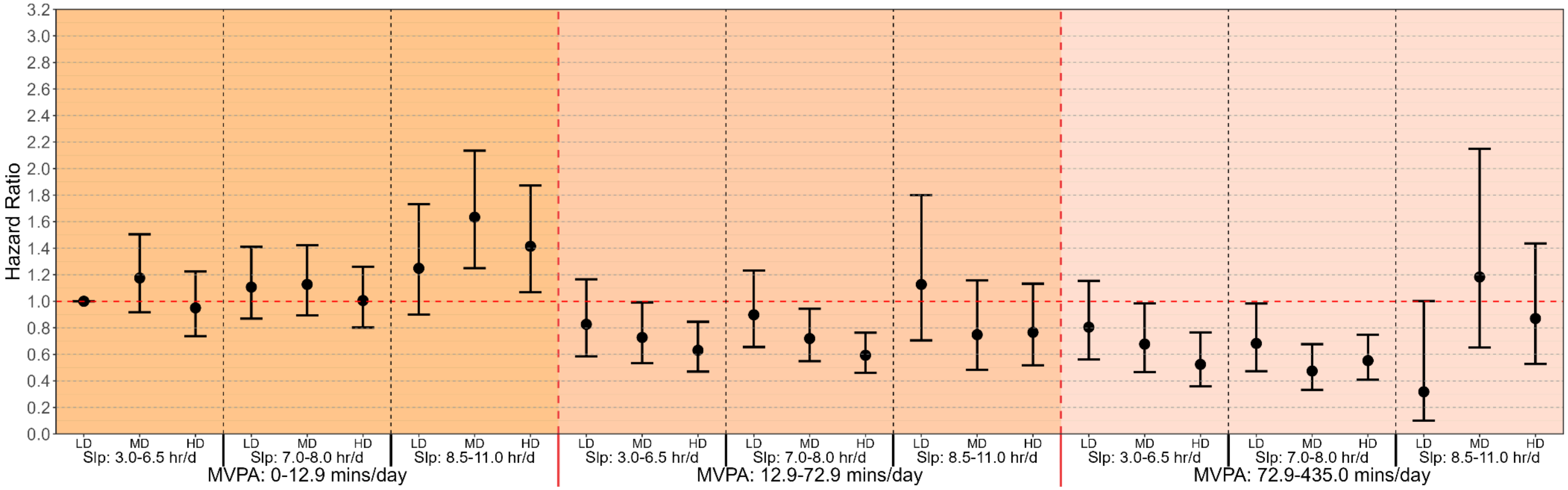


**Supplementary Figure 14 Multivariable-adjusted associations of combined sleep, moderate-to-vigorous physical activity (MVPA) and nutrition with all-cause mortality risk, excluding current smokers (n = 25,553; events = 2,088). Legend:** Forest plot presents the associations of SPAN and all-cause mortality after removing participants who were current smokers. Model is adjusted for age, sex, ethnicity, education, ratio of family income to poverty, alcohol consumption, smoking status, sedentary activity time, total energy intake, previous diagnosis of cardiovascular disease, cancer or type 2 diabetes, and family history of heart attack and diabetes. Sleep duration (hours/day), MVPA (minutes/day), and nutrition (Healthy Eating Index-2020 (HEI2020)) were treated as a joint term. The specific groups for each exposure were divided as sleep duration: 3.0-6.5 hours/day (short), 7.0-8.0 hours/day (optimal) and 8.5-11.0 hours/day (long); MVPA: 0-12.9 minutes/day (low), 12.9-72.9 minutes/day (moderate) and 72.9-435.0 minutes/day (high); HEI-2020: 12.4-45.0 (low), 45.0-56.0 (moderate), and 56.0-93.8 (high). The lowest group for all three exposures were treated as reference group. LD: low diet quality; MD: moderate diet quality; HD: high diet quality; Slp: sleep duration.

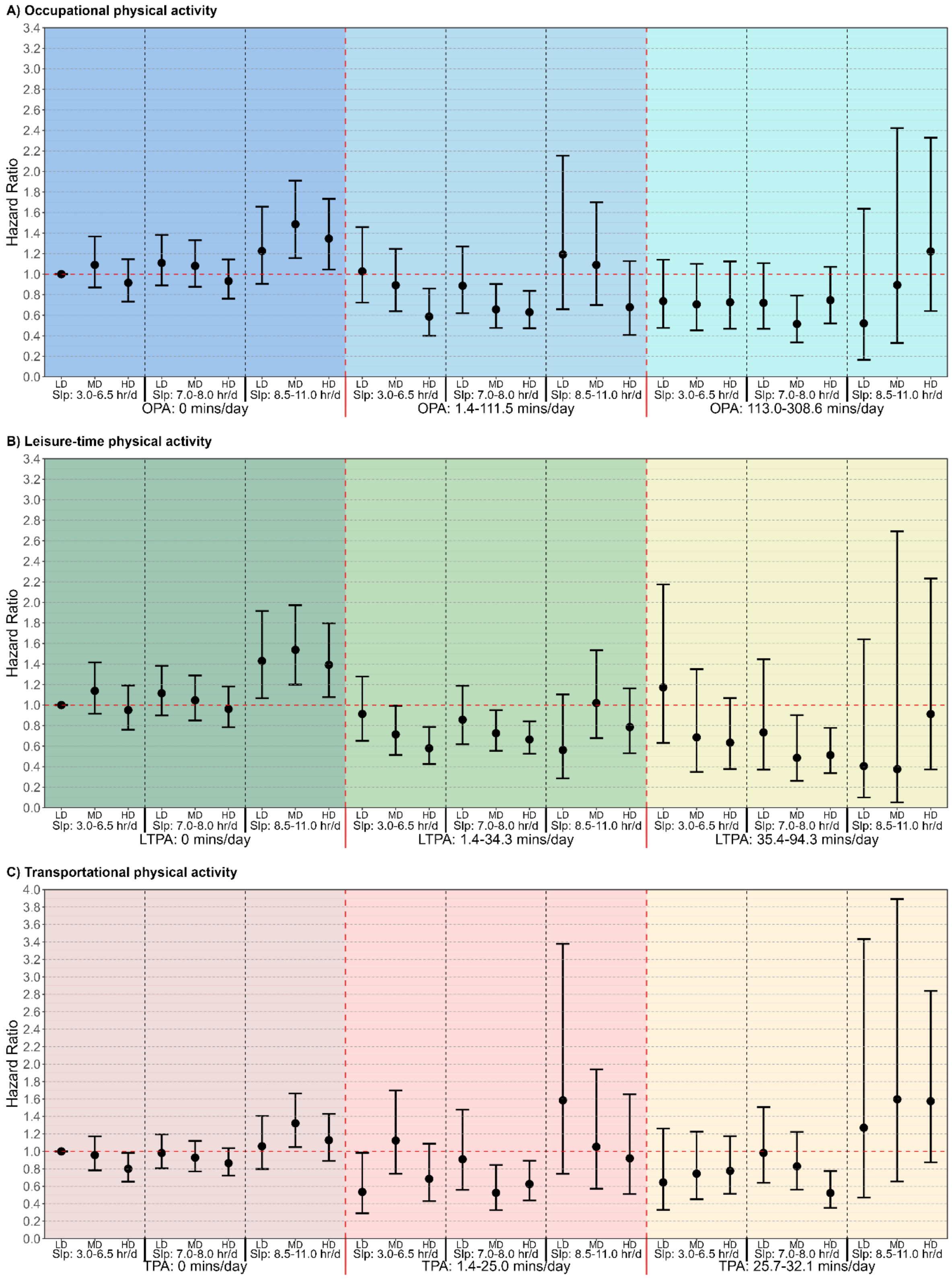


**Supplementary Figure 15 Multivariable-adjusted associations of combined sleep, physical activity domains and nutrition with all-cause mortality risk, excluding current smokers (n = 25,553; events = 2,088). Legend:** Forest plot presents the associations of SPAN and all-cause mortality after removing participants who were current smokers, across physical activity domains. The figure shows: A: Occupational physical activity (OPA); B: Leisure-time physical activity (LTPA); C: Transportational physical activity (TPA). Models are adjusted for adjusted for age, sex, ethnicity, education, ratio of family income to poverty, alcohol consumption, smoking status, sedentary activity time, total energy intake, previous diagnosis of cardiovascular disease, cancer or type 2 diabetes, and family history of heart attack and diabetes. When examining one PA domain as the primary exposure, the other two physical activity domains were adjusted in the multivariable models. Sleep duration (hours/day), OPA/LTPA/TPA (minutes/day), and nutrition (Healthy Eating Index-2020 (HEI2020)) were treated as a joint term. The specific groups for each exposure were divided as sleep duration: 3.0-6.5 hours/day (short), 7.0-8.0 hours/day (optimal) and 8.5-11.0 hours/day (long); OPA: 0 minutes/day (low), 1.4-111.5 minutes/day (moderate) and 113.0-308.6 minutes/day (high); LTPA: 0 minutes/day (low), 1.4-34.3 minutes/day (moderate) and 35.4-94.3 minutes/day (high); TPA: 0 minutes/day (low), 1.4-25.0 minutes/day (moderate) and 25.7-32.1 minutes/day (high); HEI-2020: 12.4-45.0 (low), 45.0-56.0 (moderate), and 56.0-93.8 (high). The lowest group for all three exposures in each physical activity domain were treated as reference group. LD: low diet quality; MD: moderate diet quality; HD: high diet quality; Slp: sleep duration.

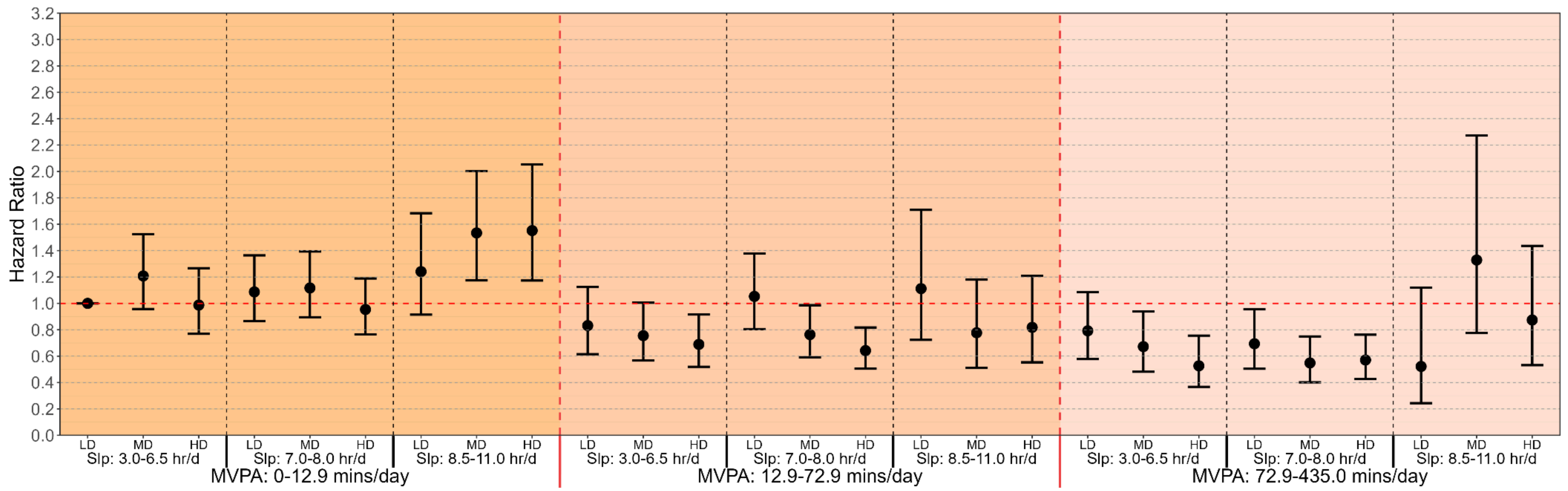


**Supplementary Figure 16 Multivariable-adjusted associations of combined sleep, moderate-to-vigorous physical activity (MVPA) and nutrition with all-cause mortality risk, excluding participants with self-reported poor health status (n = 29,107; events = 2,267). Legend:** Forest plot presents the associations of SPAN and all-cause mortality after removing participants with current poor health status. Model is adjusted for age, sex, ethnicity, education, ratio of family income to poverty, alcohol consumption, smoking status, sedentary activity time, total energy intake, previous diagnosis of cardiovascular disease, cancer or type 2 diabetes, and family history of heart attack and diabetes. Sleep duration (hours/day), MVPA (minutes/day), and nutrition (Healthy Eating Index-2020 (HEI2020)) were treated as a joint term. The specific groups for each exposure were divided as sleep duration: 3.0-6.5 hours/day (short), 7.0-8.0 hours/day (optimal) and 8.5-11.0 hours/day (long); MVPA: 0-12.9 minutes/day (low), 12.9-72.9 minutes/day (moderate) and 72.9-435.0 minutes/day (high); HEI-2020: 10.0-45.0 (low), 45.0-56.0 (moderate), and 56.0-93.8 (high). The lowest group for all three exposures were treated as reference group. LD: low diet quality; MD: moderate diet quality; HD: high diet quality; Slp: sleep duration.

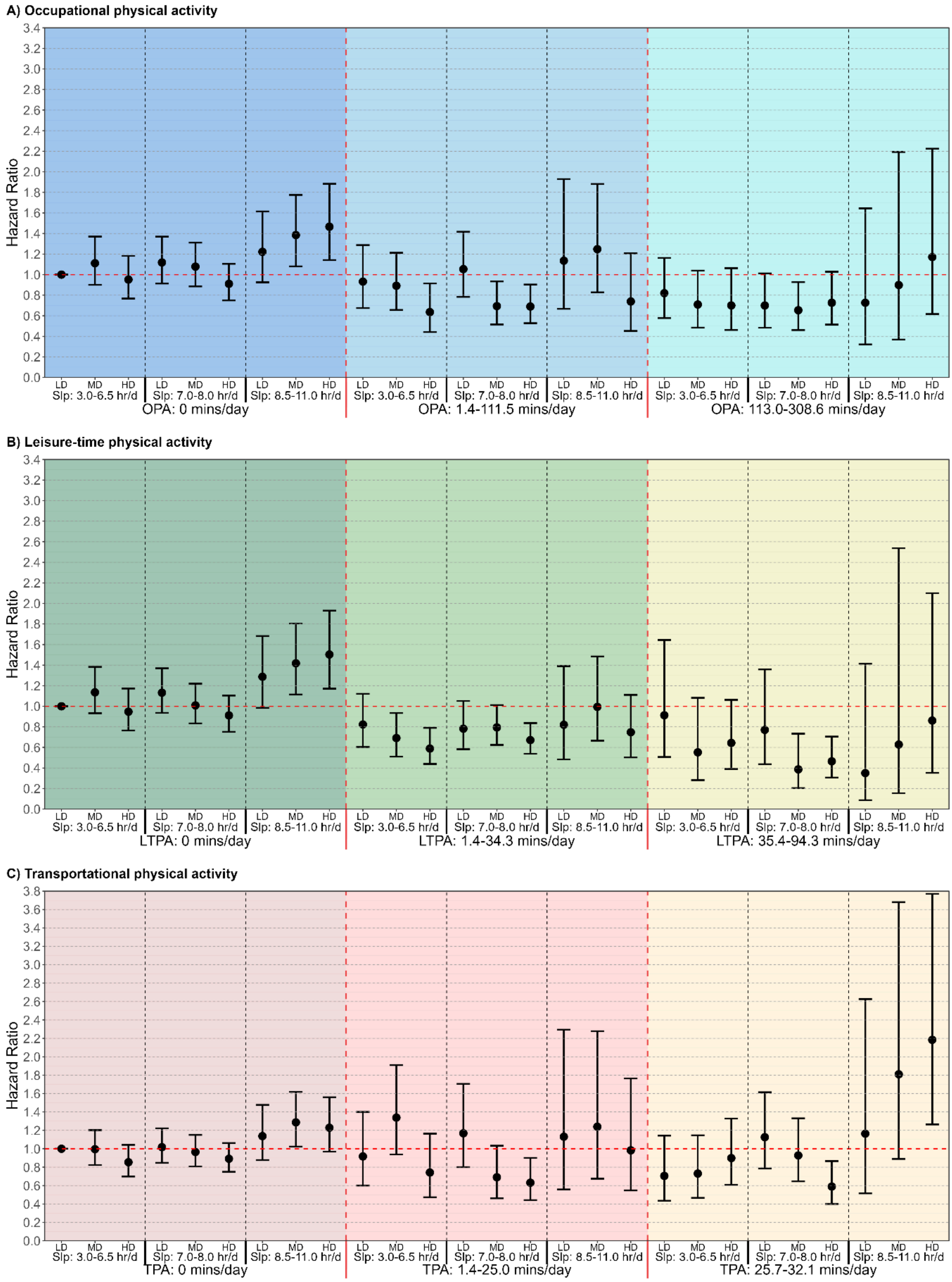


**Supplementary Figure 17 Multivariable-adjusted associations of combined sleep, physical activity domains and nutrition with all-cause mortality risk, excluding participants with current poor health status (n = 29,107; events = 2,267). Legend:** Forest plot presents the associations of SPAN and all-cause mortality after removing participants with current poor health status, across physical activity domains. The figure shows: A: Occupational physical activity (OPA); B: Leisure-time physical activity (LTPA); C: Transportational physical activity (TPA). Models are adjusted for adjusted for age, sex, ethnicity, education, ratio of family income to poverty, alcohol consumption, smoking status, sedentary activity time, total energy intake, previous diagnosis of cardiovascular disease, cancer or type 2 diabetes, and family history of heart attack and diabetes. When examining one PA domain as the primary exposure, the other two physical activity domains were adjusted in the multivariable models. Sleep duration (hours/day), OPA/LTPA/TPA (minutes/day), and nutrition (Healthy Eating Index-2020 (HEI2020)) were treated as a joint term. The specific groups for each exposure were divided as sleep duration: 3.0-6.5 hours/day (short), 7.0-8.0 hours/day (optimal) and 8.5-11.0 hours/day (long); OPA: 0 minutes/day (low), 1.4-111.5 minutes/day (moderate) and 113.0-308.6 minutes/day (high); LTPA: 0 minutes/day (low), 1.4-34.3 minutes/day (moderate) and 35.4-94.3 minutes/day (high); TPA: 0 minutes/day (low), 1.4-25.0 minutes/day (moderate) and 25.7-32.1 minutes/day (high); HEI-2020: 10.0-45.0 (low), 45.0-56.0 (moderate), and 56.0-93.8 (high). The lowest group for all three exposures in each physical activity domain were treated as reference group. LD: low diet quality; MD: moderate diet quality; HD: high diet quality; Slp: sleep duration.

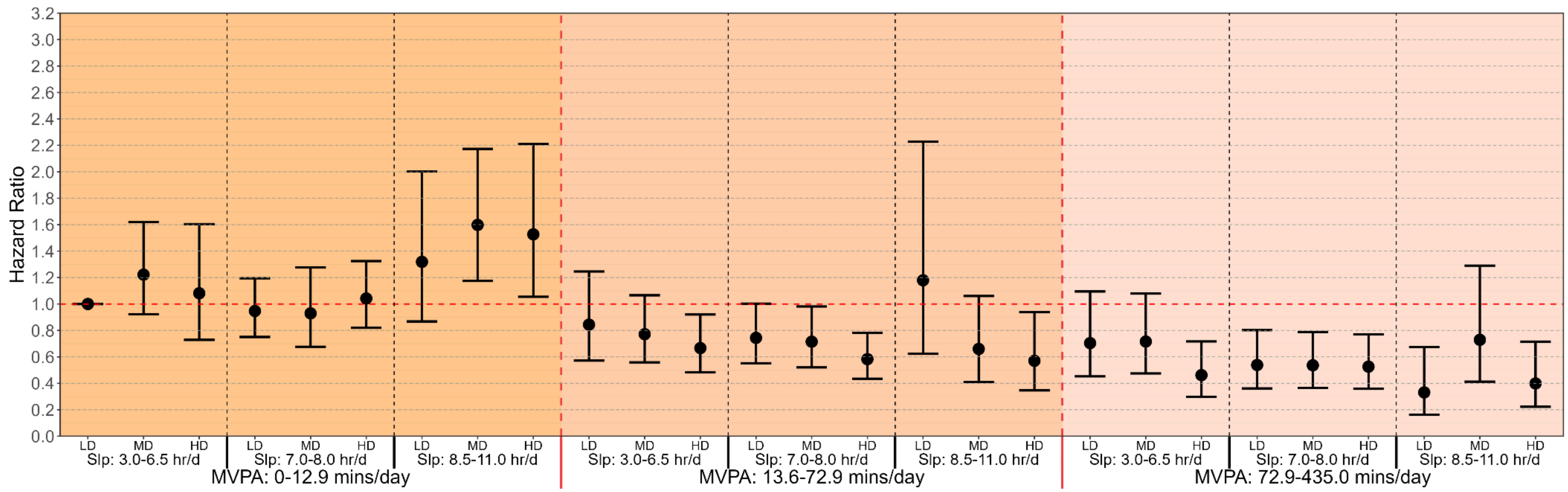


**Supplementary Figure 18 Multivariable-adjusted associations of combined sleep, moderate-to-vigorous physical activity (MVPA) and nutrition with all-cause mortality risk, using day-1 sample weights (n = 31,875; events = 2,623). Legend:** We used day 1 dietary recall sample weight to represent the general US population. Model is adjusted for age, sex, ethnicity, education, ratio of family income to poverty, alcohol consumption, smoking status, sedentary activity time, total energy intake, previous diagnosis of cardiovascular disease, cancer or type 2 diabetes, and family history of heart attack and diabetes. Sleep duration (hours/day), MVPA (minutes/day), and nutrition were treated as a joint term. The specific groups for each exposure were divided as sleep duration: 3.0-6.5 hours/day (short), 7.0-8.0 hours/day (optimal) and 8.5-11.0 hours/day (long); MVPA: 0-12.9 minutes/day (low), 12.9-72.9 minutes/day (moderate) and 72.9-435.0 minutes/day (high); HEI-2020: 0.0-43.8 (low), 43.8-56.3 (moderate), and 56.3-97.9 (high). The lowest group for all three exposures were treated as reference group. LD: low diet quality; MD: moderate diet quality; HD: high diet quality; Slp: sleep duration.

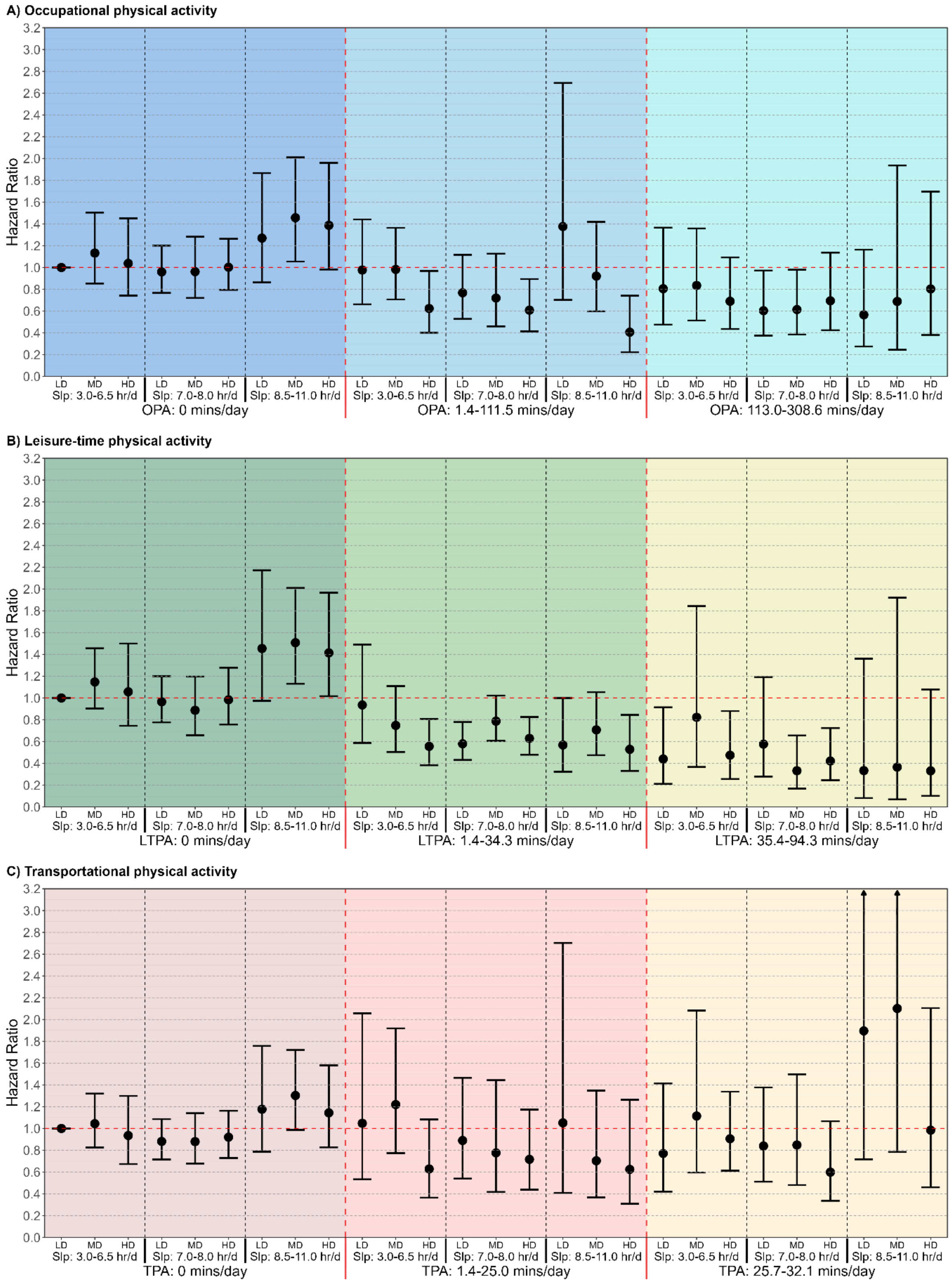


**Supplementary Figure 19 Multivariable-adjusted associations of combined sleep, physical activity domains and nutrition with all-cause mortality risk, using day-1 sample weights (n = 31,875; events = 2,623). Legend:** We used day 1 dietary recall sample weight to represent the general US population, across physical activity domains. The figure shows: A: Occupational physical activity (OPA); B: Leisure-time physical activity (LTPA); C: Transportational physical activity (TPA). Models are adjusted for adjusted for age, sex, ethnicity, education, ratio of family income to poverty, alcohol consumption, smoking status, sedentary activity time, total energy intake, previous diagnosis of cardiovascular disease, cancer or type 2 diabetes, and family history of heart attack and diabetes. When examining one PA domain as the primary exposure, the other two physical activity domains were adjusted in the multivariable models. Sleep duration (hours/day), OPA/LTPA/TPA (minutes/day), and nutrition were treated as a joint term. The specific groups for each exposure were divided as sleep duration: 3.0-6.5 hours/day (short), 7.0-8.0 hours/day (optimal) and 8.5-11.0 hours/day (long); OPA: 0 minutes/day (low), 1.4-111.5 minutes/day (moderate) and 113.0-308.6 minutes/day (high); LTPA: 0 minutes/day (low), 1.4-34.3 minutes/day (moderate) and 35.4-94.3 minutes/day (high); TPA: 0 minutes/day (low), 1.4-25.0 minutes/day (moderate) and 25.7-32.1 minutes/day (high); HEI-2020: 0.0-43.8 (low), 43.8-56.3 (moderate), and 56.3-97.9 (high). The lowest group for all three exposures in each physical activity domain were treated as reference group. LD: low diet quality; MD: moderate diet quality; HD: high diet quality; Slp: sleep duration.

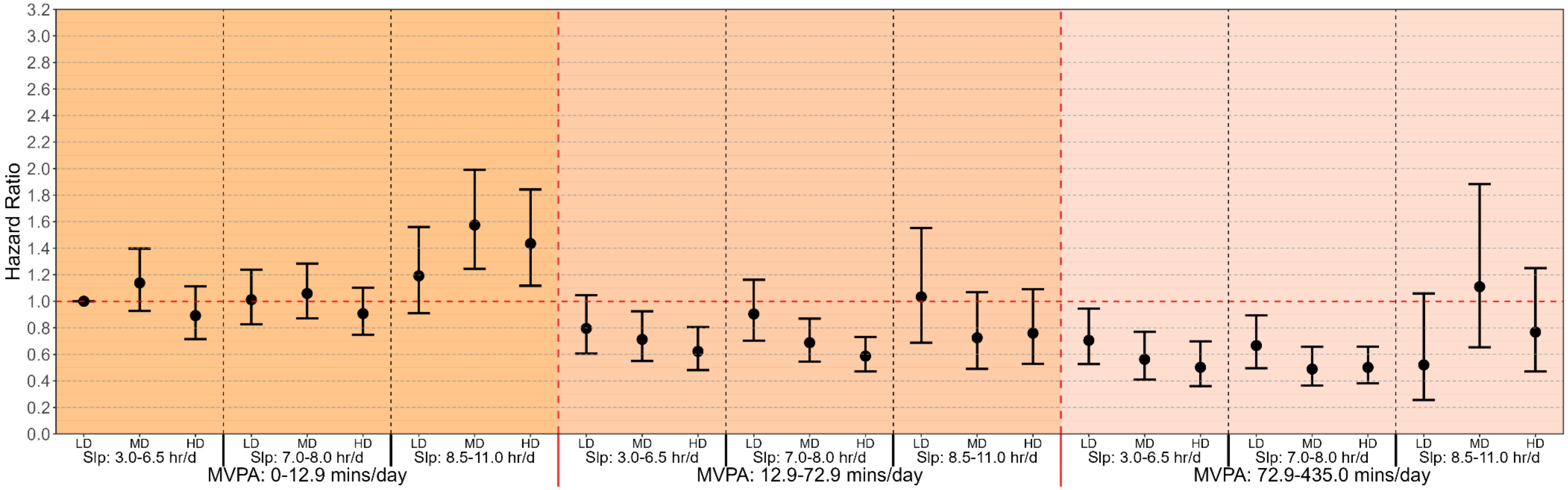


**Supplementary Figure 20 Multivariable-adjusted associations of combined sleep, moderate-to-vigorous physical activity (MVPA) and nutrition with all-cause mortality risk, using Alternative Healthy Eating Index-2010 (AHEI-2010) [2] (n = 31,875; events = 2,623). Legend:** Forest plot uses AHEI-2010 as an alternative index for diet quality, which includes 11 components with a total score from 0 to 110 [2]. Higher scores indicate a better healthy food adherence. As AHEI-2010 contains alcohol as one component, model here is adjusted for age, sex, ethnicity, education, ratio of family income to poverty, smoking status, sedentary activity time, total energy intake, previous diagnosis of cardiovascular disease, cancer or type 2 diabetes, and family history of heart attack and diabetes. Sleep duration (hours/day), MVPA (minutes/day), and nutrition were treated as a joint term. The specific groups for each exposure were divided as sleep duration: 3.0-6.5 hours/day (short), 7.0-8.0 hours/day (optimal) and 8.5-11.0 hours/day (long); MVPA: 0-12.9 minutes/day (low), 12.9-72.9 minutes/day (moderate) and 72.9-435.0 minutes/day (high); AHEI-2010: 4.3-32.4 (low), 32.4-42.7 (moderate), and 42.7-87.6 (high). The lowest group for all three exposures were treated as reference group. LD: low diet quality; MD: moderate diet quality; HD: high diet quality; Slp: sleep duration.

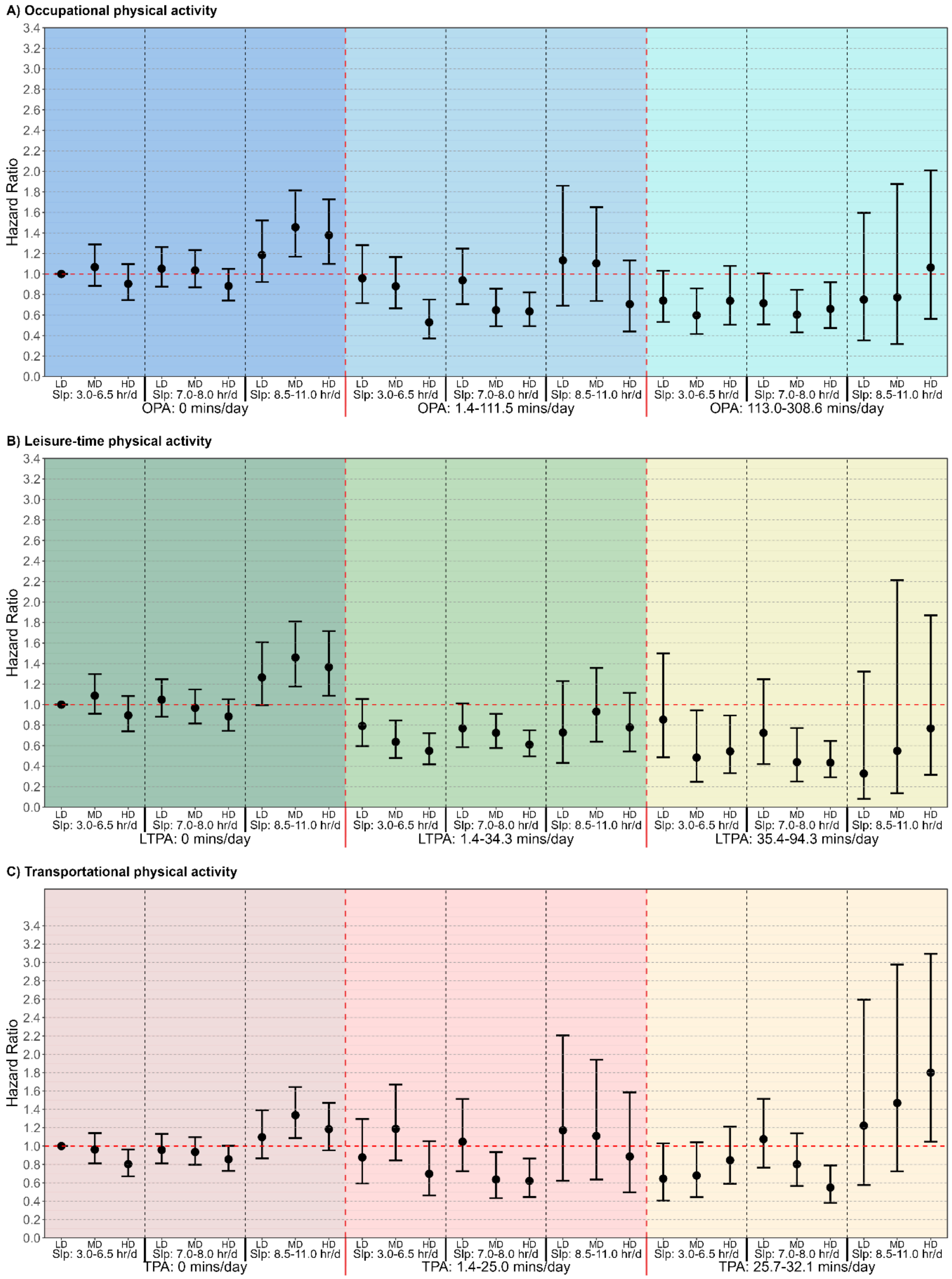


**Supplementary Figure 21 Multivariable-adjusted associations of combined sleep, physical activity domains and nutrition with all-cause mortality risk, using Alternative Healthy Eating Index-2010 (AHEI-2010) [(2)] (n = 31,875; events = 2,623). Legend:** Forest plot uses AHEI-2010 as an alternative index across physical activity domains for diet quality, which includes 11 components with a total score from 0 to 110 [(2)]. Higher scores indicate a better healthy food adherence. As AHEI-2010 contains alcohol as one component, models here are adjusted for age, sex, ethnicity, education, ratio of family income to poverty, smoking status, sedentary activity time, total energy intake, previous diagnosis of cardiovascular disease, cancer or type 2 diabetes, and family history of heart attack and diabetes. When examining one PA domain as the primary exposure, the other two physical activity domains were adjusted in the multivariable models. The figure shows: A: Occupational physical activity (OPA); B: Leisure-time physical activity (LTPA); C: Transportational physical activity (TPA). Sleep duration (hours/day), OPA/LTPA/TPA (minutes/day), and nutrition were treated as a joint term. The specific groups for each exposure were divided as sleep duration: 3.0-6.5 hours/day (short), 7.0-8.0 hours/day (optimal) and 8.5-11.0 hours/day (long); OPA: 0 minutes/day (low), 1.4-111.5 minutes/day (moderate) and 113.0-308.6 minutes/day (high); LTPA: 0 minutes/day (low), 1.4-34.3 minutes/day (moderate) and 35.4-94.3 minutes/day (high); TPA: 0 minutes/day (low), 1.4-25.0 minutes/day (moderate) and 25.7-32.1 minutes/day (high); AHEI-2010: 4.3-32.4 (low), 32.4-42.7 (moderate), and 42.7-87.6 (high). The lowest group for all three exposures in each physical activity domain were treated as reference group. LD: low diet quality; MD: moderate diet quality; HD: high diet quality; Slp: sleep duration.

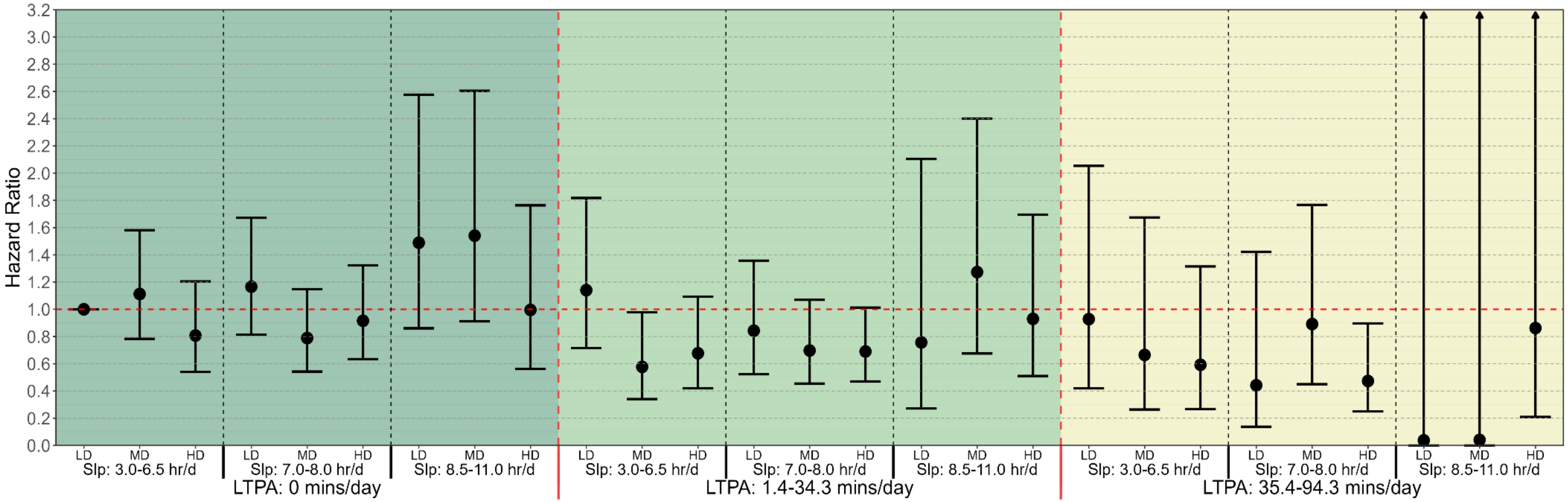


**Supplementary Figure 22 Multivariable-adjusted associations of combined sleep, leisure-time physical activity (LTPA) and nutrition with all-cause mortality risk, excluding participants with no physical activity from occupation and work (n = 13,133; events = 690). Legend:** Forest plot excludes participants with 0 minutes of occupational physical activity. Model here is adjusted for age, sex, ethnicity, education, occupational physical activity, transportational physical activity, ratio of family income to poverty, smoking status, alcohol status, sedentary activity time, total energy intake, previous diagnosis of cardiovascular disease, cancer or type 2 diabetes, and family history of heart attack and diabetes. Sleep duration (hours/day), LTPA (minutes/day), and nutrition were treated as a joint term. The specific groups for each exposure were divided as sleep duration: 3.0-6.5 hours/day (short), 7.0-8.0 hours/day (optimal) and 8.5-11.0 hours/day (long); LTPA: 0minutes/day (low), 1.4-34.3 minutes/day (moderate) and 35.4-94.3 minutes/day (high); HEI-2020: 12.7-43.8 (low), 43.8-54.5 (moderate), and 54.5-92.0 (high). The lowest group for all three exposures were treated as reference group. LD: low diet quality; MD: moderate diet quality; HD: high diet quality; Slp: sleep duration.

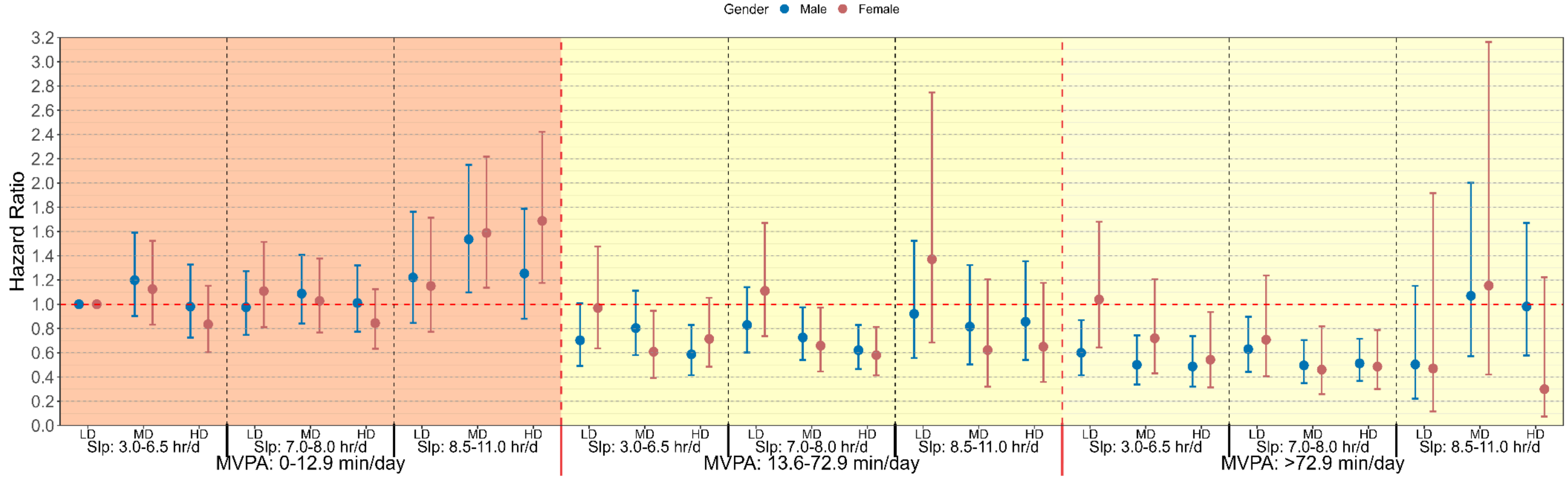


**Supplementary Figure 23 Multivariable-adjusted associations of combined sleep, moderate-to-vigorous physical activity (MVPA) and nutrition with all-cause mortality risk across sex groups (n = 31,875, events = 2,623; Male: n = 15,504, events = 1,506; Female: n = 16,371, events = 1,117). Legend:** Models are adjusted for age, ethnicity, education, ratio of family income to poverty, alcohol consumption, smoking status, sedentary activity time, total energy intake, previous diagnosis of cardiovascular disease, cancer or type 2 diabetes, and family history of heart attack and diabetes. Sleep duration (hours/day), MVPA (minutes/day), and nutrition (Healthy Eating Index-2020 (HEI2020)) were treated as a joint term. The specific groups for each exposure were divided based on overall sample size for better comparisons as sleep duration: 3.0-6.5 hours/day (short), 7.0-8.0 hours/day (optimal) and 8.5-11.0 hours/day (long); MVPA: 0-12.9 minutes/day (low), 13.6-72.9 minutes/day (moderate) and 72.9-435.0 minutes/day (high); HEI-2020: 10.0-45.0 (low), 45.0-56.0 (moderate), and 56.0-93.8 (high). The lowest group for all three exposures were treated as reference group. LD: low diet quality; MD: moderate diet quality; HD: high diet quality; Slp: sleep duration.

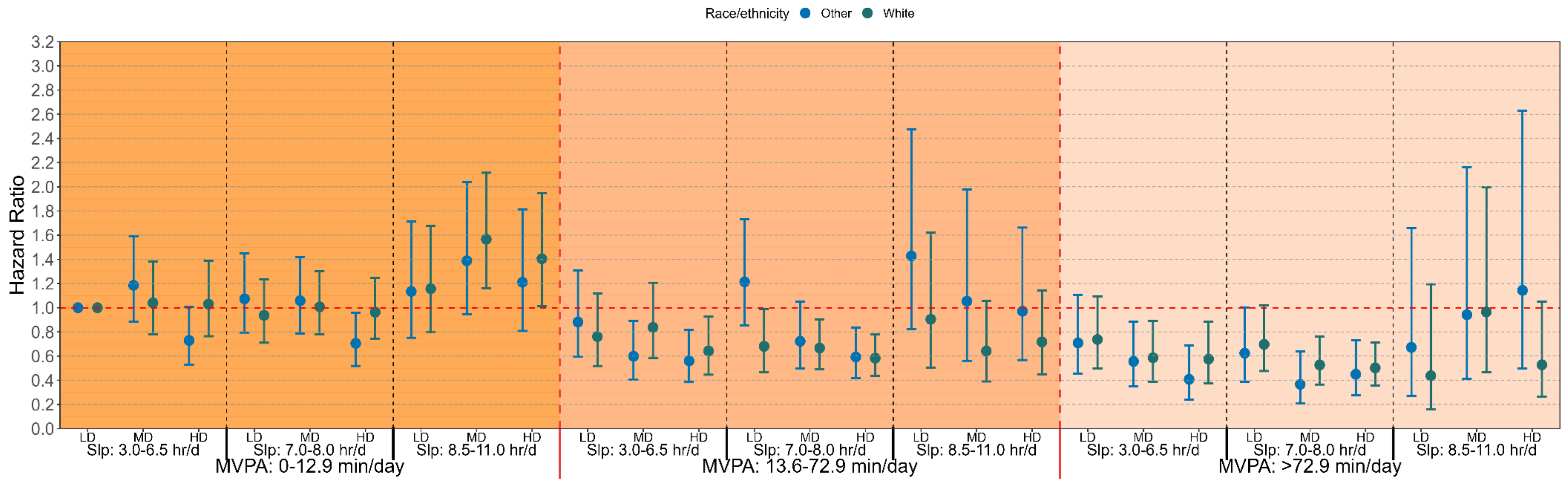


**Supplementary Figure 24 Multivariable-adjusted associations of combined sleep, moderate-to-vigorous physical activity (MVPA) and nutrition with all-cause mortality risk across different ethnic groups (n = 31,875, events = 2,623; White: n = 13,021, events = 1,578; Others: n = 18,854, events = 1,045). Legend:** To save sample size within each group, we divided race group as 1) White: non-Hispanic white, and 2) Others: Non-Hispanic Black, Hispanic and Mexican American, and Other. Models are adjusted for age, sex, education, ratio of family income to poverty, alcohol consumption, smoking status, sedentary activity time, total energy intake, previous diagnosis of cardiovascular disease, cancer or type 2 diabetes, and family history of heart attack and diabetes. Sleep duration (hours/day), MVPA (minutes/day), and nutrition (Healthy Eating Index-2020 (HEI2020)) were treated as a joint term. The specific groups for each exposure were divided based on overall sample size for better comparisons across groups as sleep duration: 3.0-6.5 hours/day (short), 7.0-8.0 hours/day (optimal) and 8.5-11.0 hours/day (long); MVPA: 0-12.9 minutes/day (low), 13.6-72.9 minutes/day (moderate) and 72.9-435.0 minutes/day (high); HEI-2020: 10.0-45.0 (low), 45.0-56.0 (moderate), and 56.0-93.8 (high). The lowest group for all three exposures were treated as reference group. LD: low diet quality; MD: moderate diet quality; HD: high diet quality; Slp: sleep duration.

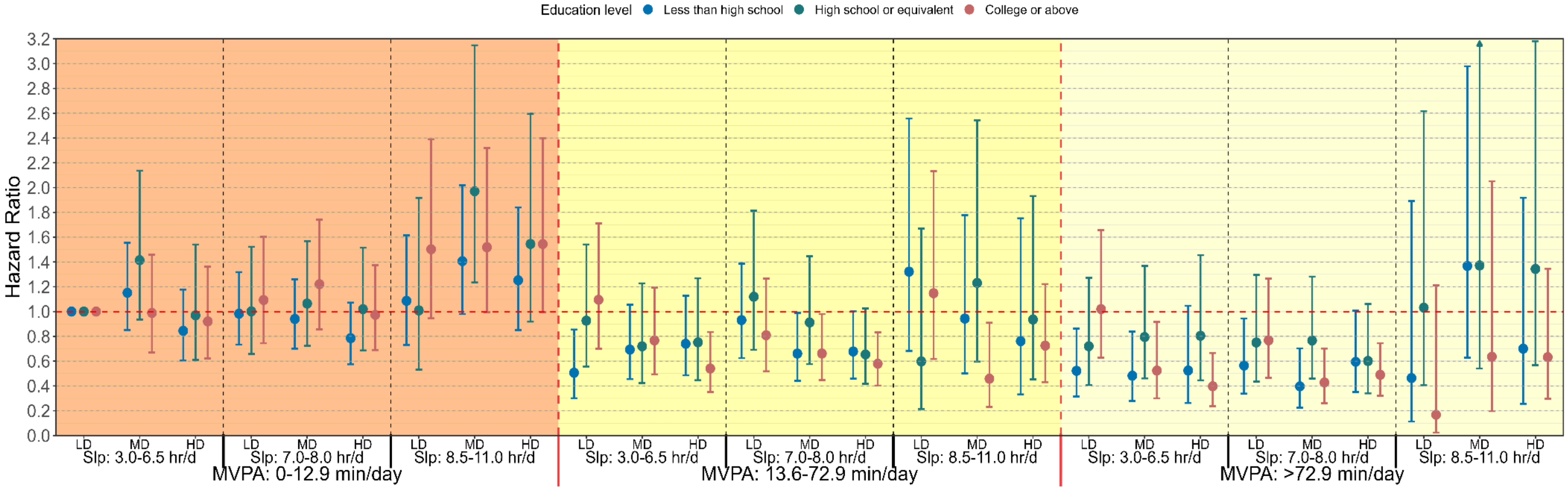


**Supplementary Figure 25 Multivariable-adjusted associations of combined sleep, moderate-to-vigorous physical activity (MVPA) and nutrition with all-cause mortality risk across different educational levels (n = 31,875, events = 2,623; Less than high school: n = 7,631, events = 944; High school or equivalent: n = 7,267, events = 660; College or above: n = 16,977, events = 1,019). Legend:** Models are adjusted for age, sex, ratio of family income to poverty, alcohol consumption, smoking status, sedentary activity time, total energy intake, previous diagnosis of cardiovascular disease, cancer or type 2 diabetes, and family history of heart attack and diabetes. Sleep duration (hours/day), MVPA (minutes/day), and nutrition (Healthy Eating Index-2020 (HEI2020)) were treated as a joint term. The specific groups for each exposure were divided based on overall sample size for better comparisons across groups as sleep duration: 3.0-6.5 hours/day (short), 7.0-8.0 hours/day (optimal) and 8.5-11.0 hours/day (long); MVPA: 0-12.9 minutes/day (low), 13.6-72.9 minutes/day (moderate) and 72.9-435.0 minutes/day (high); HEI-2020: 10.0-45.0 (low), 45.0-56.0 (moderate), and 56.0-93.8 (high). The lowest group for all three exposures were treated as reference group. LD: low diet quality; MD: moderate diet quality; HD: high diet quality; Slp: sleep duration.

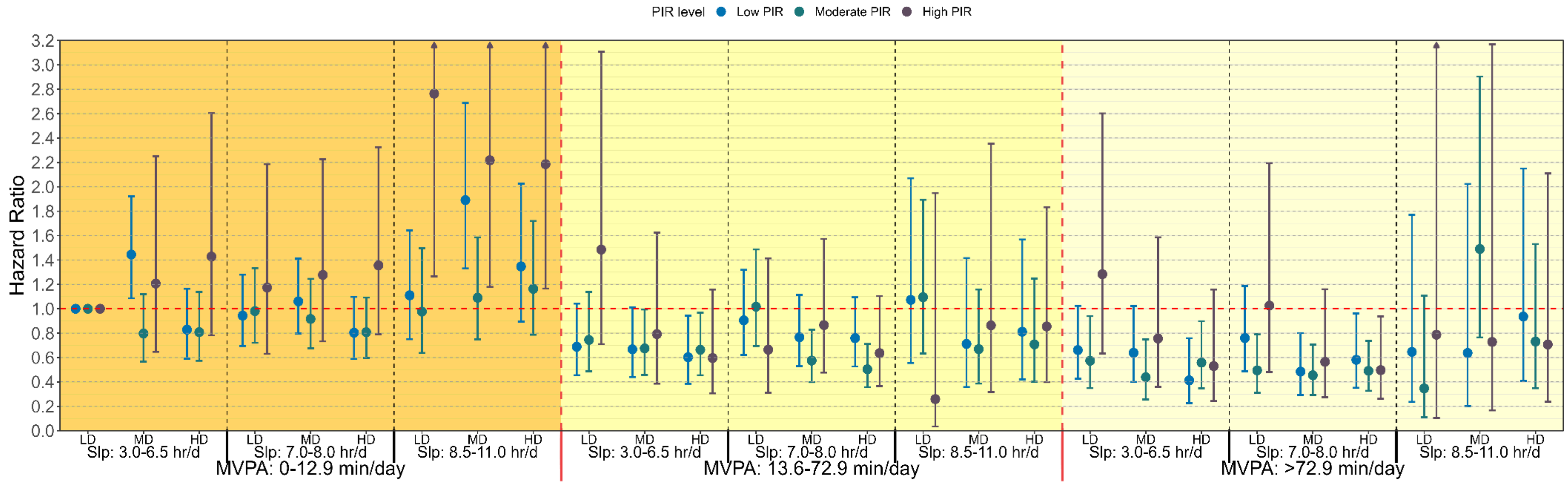


**Supplementary Figure 26 Multivariable-adjusted associations of combined sleep, moderate-to-vigorous physical activity (MVPA) and nutrition with all-cause mortality risk across different Poverty income ratio (PIR) groups (n = 31,875, events = 2,623; Low PIR: n = 10,590, events = 994; Moderate PIR: n = 11,969, events = 1,141; High PIR: n = 9,316, events = 488). Legend:** Models are adjusted for age, sex, education, alcohol consumption, smoking status, sedentary activity time, total energy intake, previous diagnosis of cardiovascular disease, cancer or type 2 diabetes, and family history of heart attack and diabetes. Sleep duration (hours/day), MVPA (minutes/day), and nutrition (Healthy Eating Index-2020 (HEI2020)) were treated as a joint term. The specific groups for each exposure were divided based on overall sample size for better comparisons across groups as sleep duration: 3.0-6.5 hours/day (short), 7.0-8.0 hours/day (optimal) and 8.5-11.0 hours/day (long); MVPA: 0-12.9 minutes/day (low), 13.6-72.9 minutes/day (moderate) and 72.9-435.0 minutes/day (high); HEI-2020: 10.0-45.0 (low), 45.0-56.0 (moderate), and 56.0-93.8 (high). The lowest group for all three exposures were treated as reference group. Low PIR: < 1.3; Moderate PIR: 1.3-3.5; High PIR: >3.5. LD: low diet quality; MD: moderate diet quality; HD: high diet quality; Slp: sleep duration.

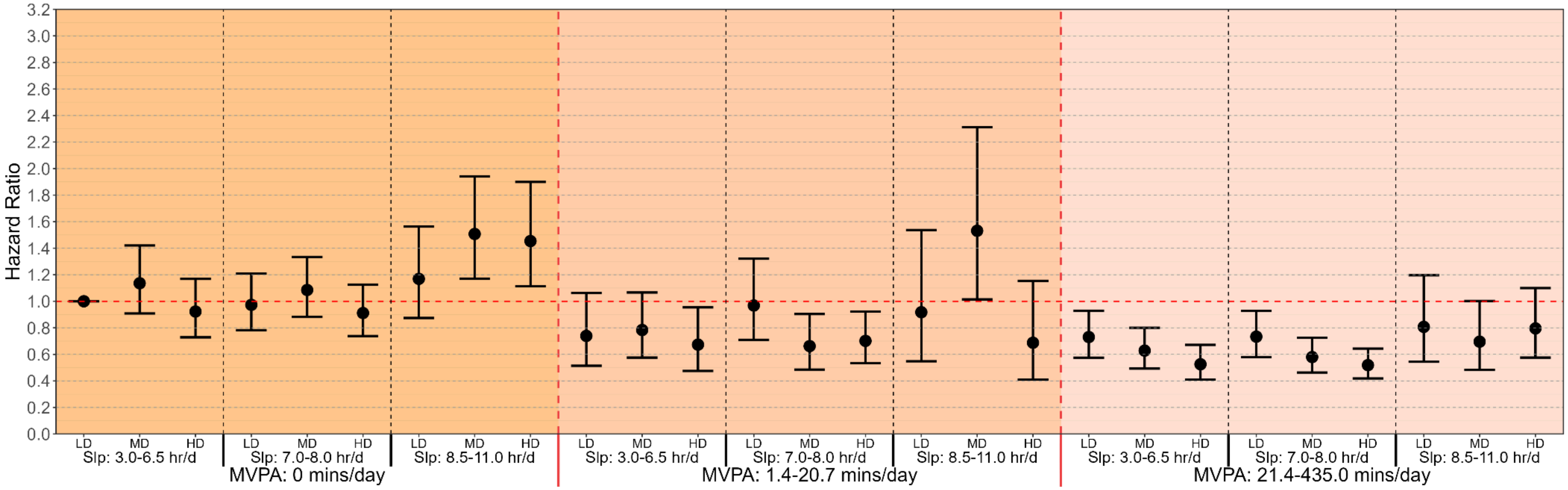


**Supplementary Figure 27 Multivariable-adjusted associations of combined sleep, WHO-guideline based moderate-to-vigorous physical activity (MVPA) and nutrition with all-cause mortality risk (n = 31,875; events = 2,623). Legend:** Based on the World Health Organization guideline recommending at least 150 min/week of MVPA[3], equivalent to approximately 21.4 min/day, we classified MVPA into three groups: 0 min/day as low, 1.4–20.7 min/day as moderate, and ≥21.4 min/day as high. Model is adjusted for age, sex, ethnicity, education, ratio of family income to poverty, alcohol consumption, smoking status, sedentary activity time, total energy intake, previous diagnosis of cardiovascular disease, cancer or type 2 diabetes, and family history of heart attack and diabetes. Sleep duration (hours/day), MVPA (minutes/day), and nutrition (Healthy Eating Index-2020 (HEI2020)) were treated as a joint term. The specific groups for each exposure were divided as sleep duration: 3.0-6.5 hours/day (short), 7.0-8.0 hours/day (optimal) and 8.5-11.0 hours/day (long); HEI-2020: 10.0-45.0 (low), 45.0-56.0 (moderate), and 56.0-93.8 (high). The lowest group for all three exposures were treated as reference group. LD: low diet quality; MD: moderate diet quality; HD: high diet quality; Slp: sleep duration.

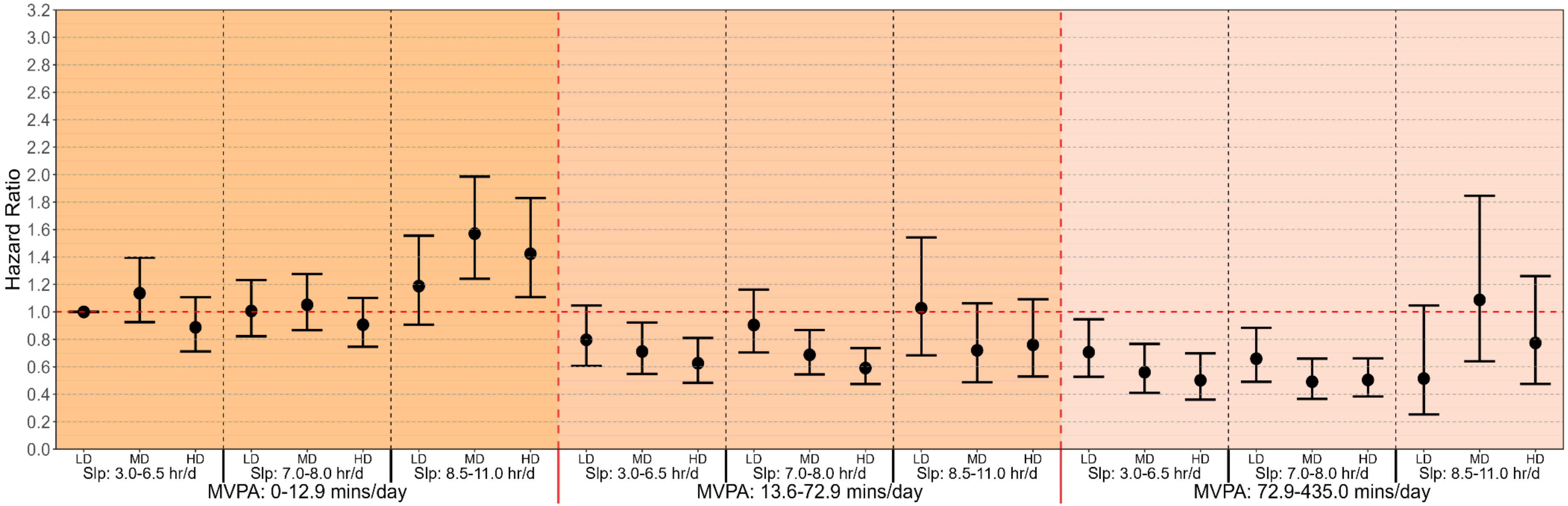


**Supplementary Figure 28 Multivariable-adjusted associations of combined sleep, moderate-to-vigorous physical activity (MVPA) and nutrition with all-cause mortality risk, not adjusting for total energy intake (n = 31,875; events = 2,623). Legend:** Model is adjusted for age, sex, ethnicity, education, ratio of family income to poverty, alcohol consumption, smoking status, sedentary activity time, previous diagnosis of cardiovascular disease, cancer or type 2 diabetes, and family history of heart attack and diabetes. Sleep duration (hours/day), MVPA (minutes/day), and nutrition (Healthy Eating Index-2020 (HEI2020)) were treated as a joint term. The specific groups for each exposure were divided as sleep duration: 3.0-6.5 hours/day (short), 7.0-8.0 hours/day (optimal) and 8.5-11.0 hours/day (long); MVPA: 0-12.9 minutes/day (low), 12.9-72.9 minutes/day (moderate) and 72.9-435.0 minutes/day (high); HEI-2020: 10.0-45.0 (low), 45.0-56.0 (moderate), and 56.0-93.8 (high). The lowest group for all three exposures were treated as reference group. LD: low diet quality; MD: moderate diet quality; HD: high diet quality; Slp: sleep duration.

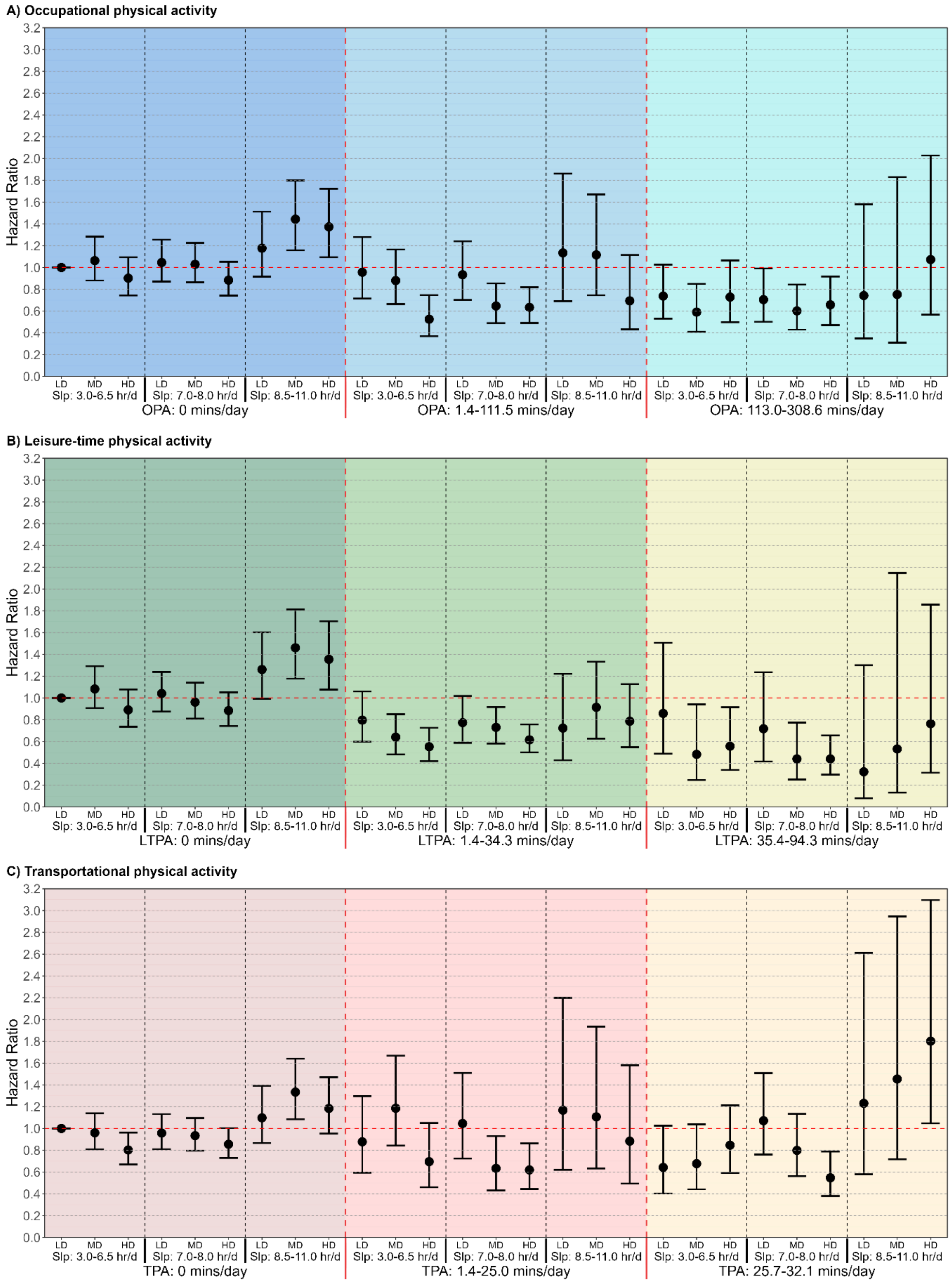


**Supplementary Figure 29 Multivariable-adjusted associations of combined sleep, physical activity domains and nutrition with all-cause mortality risk, not adjusting for total energy intake (n = 31,875; events = 2,623). Legend:** Daily total energy intake was not adjusted in the model. The figure shows: A: Occupational physical activity (OPA); B: Leisure-time physical activity (LTPA); C: Transportational physical activity (TPA). Models are adjusted for adjusted for age, sex, ethnicity, education, ratio of family income to poverty, alcohol consumption, smoking status, sedentary activity time, previous diagnosis of cardiovascular disease, cancer or type 2 diabetes, and family history of heart attack and diabetes. When examining one PA domain as the primary exposure, the other two physical activity domains were adjusted in the multivariable models. Sleep duration (hours/day), OPA/LTPA/TPA (minutes/day), and nutrition were treated as a joint term. The specific groups for each exposure were divided as sleep duration: 3.0-6.5 hours/day (short), 7.0-8.0 hours/day (optimal) and 8.5-11.0 hours/day (long); OPA: 0 minutes/day (low), 1.4-111.5 minutes/day (moderate) and 113.0-308.6 minutes/day (high); LTPA: 0 minutes/day (low), 1.4-34.3 minutes/day (moderate) and 35.4-94.3 minutes/day (high); TPA: 0 minutes/day (low), 1.4-25.0 minutes/day (moderate) and 25.7-32.1 minutes/day (high); HEI-2020: 10.0-45.3 (low), 45.3-56.2 (moderate), and 56.2-93.8 (high). The lowest group for all three exposures in each physical activity domain were treated as reference group. LD: low diet quality; MD: moderate diet quality; HD: high diet quality; Slp: sleep duration.

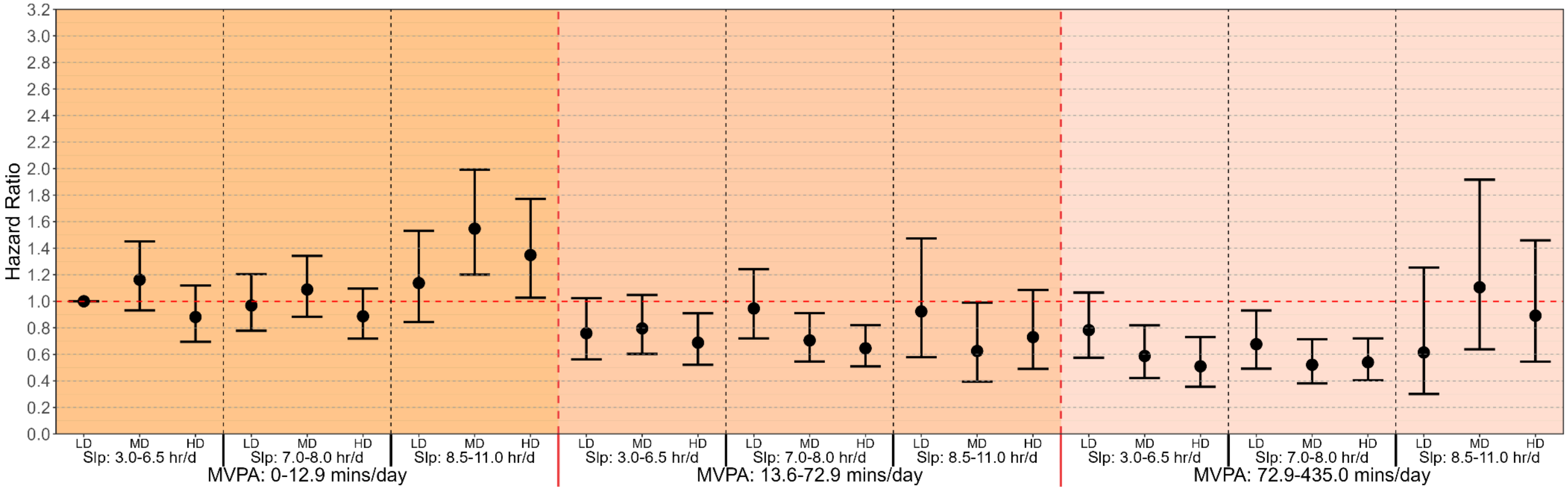


**Supplementary Figure 30 Multivariable-adjusted associations of combined sleep, moderate-to-vigorous physical activity (MVPA) and nutrition with all-cause mortality risk, using complete data (n = 26,454; events = 2,245). Legend:** Sample with complete data was used. Model is adjusted for age, sex, ethnicity, education, ratio of family income to poverty, alcohol consumption, smoking status, sedentary activity time, previous diagnosis of cardiovascular disease, cancer or type 2 diabetes, and family history of heart attack and diabetes. Sleep duration (hours/day), MVPA (minutes/day), and nutrition (Healthy Eating Index-2020 (HEI2020)) were treated as a joint term. The specific groups for each exposure were divided as sleep duration: 3.0-6.5 hours/day (short), 7.0-8.0 hours/day (optimal) and 8.5-11.0 hours/day (long); MVPA: 0-12.9 minutes/day (low), 12.9-72.9 minutes/day (moderate) and 72.9-435.0 minutes/day (high); HEI-2020: 10.0-45.0 (low), 45.0-56.0 (moderate), and 56.0-93.8 (high). The lowest group for all three exposures were treated as reference group. LD: low diet quality; MD: moderate diet quality; HD: high diet quality; Slp: sleep duration.

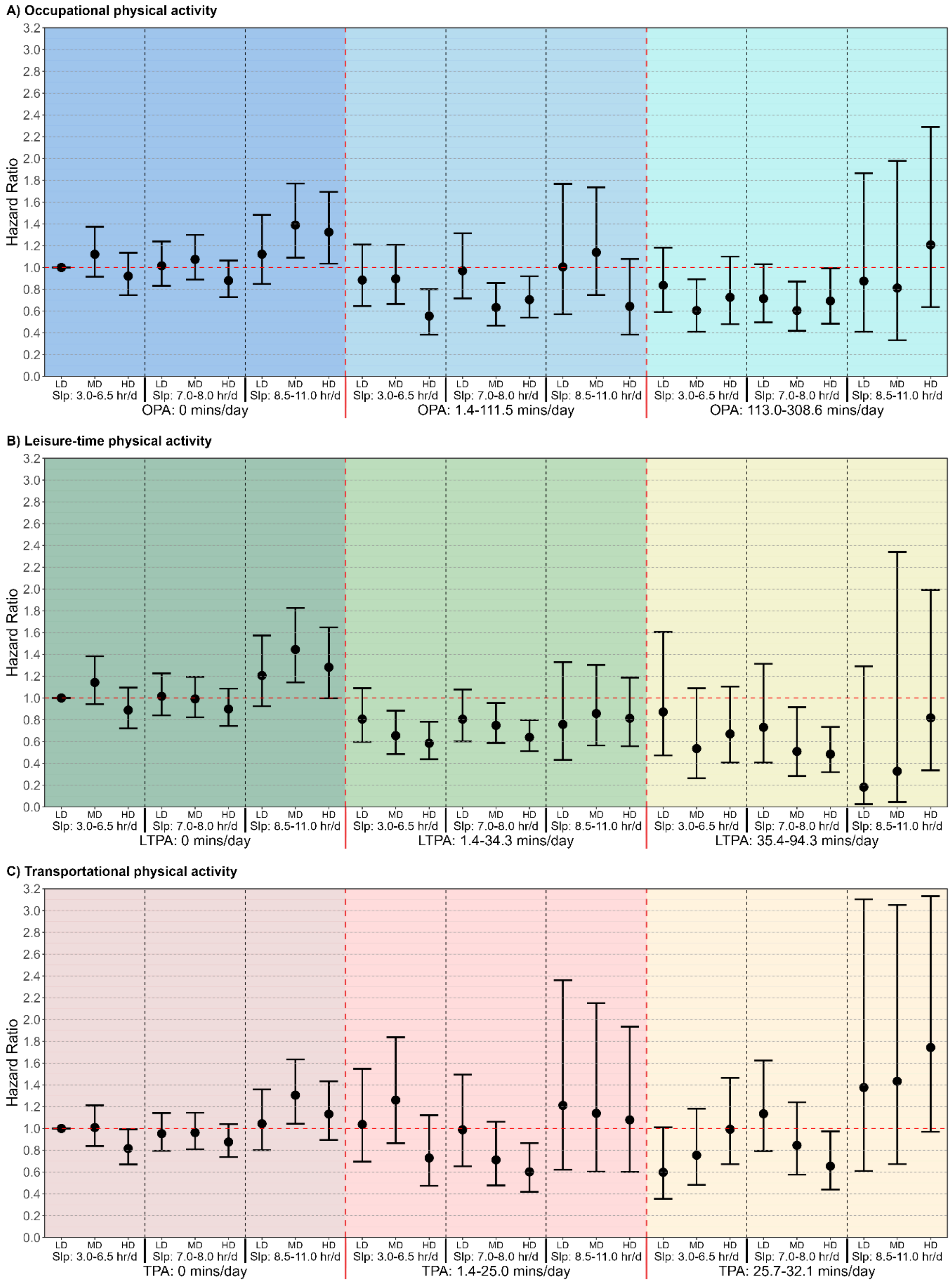


**Supplementary Figure 31 Multivariable-adjusted associations of combined sleep, physical activity domains and nutrition with all-cause mortality risk, using complete data (n = 26,454; events = 2,245).**
**Legend:** Sample without missing values was used**.** The figure shows: A: Occupational physical activity (OPA); B: Leisure-time physical activity (LTPA); C: Transportational physical activity (TPA). Models are adjusted for

adjusted for age, sex, ethnicity, education, ratio of family income to poverty, alcohol consumption, smoking status, sedentary activity time, previous diagnosis of cardiovascular disease, cancer or type 2 diabetes, and family history of heart attack and diabetes. When examining one PA domain as the primary exposure, the other two physical activity domains were adjusted in the multivariable models. Sleep duration (hours/day), OPA/LTPA/TPA (minutes/day), and nutrition were treated as a joint term. The specific groups for each exposure were divided as sleep duration: 3.0-6.5 hours/day (short), 7.0-8.0 hours/day (optimal) and 8.5-11.0 hours/day (long); OPA: 0 minutes/day (low), 1.4-111.5 minutes/day (moderate) and 113.0-308.6 minutes/day (high); LTPA: 0 minutes/day (low), 1.4-34.3 minutes/day (moderate) and 35.4-94.3 minutes/day (high); TPA: 0 minutes/day (low), 1.4-25.0 minutes/day (moderate) and 25.7-32.1 minutes/day (high); HEI-2020: 10.0-45.0 (low), 45.0-56.2 (moderate), and 56.2-93.8 (high). The lowest group for all three exposures in each physical activity domain were treated as reference group. LD: low diet quality; MD: moderate diet quality; HD: high diet quality; Slp: sleep duration.

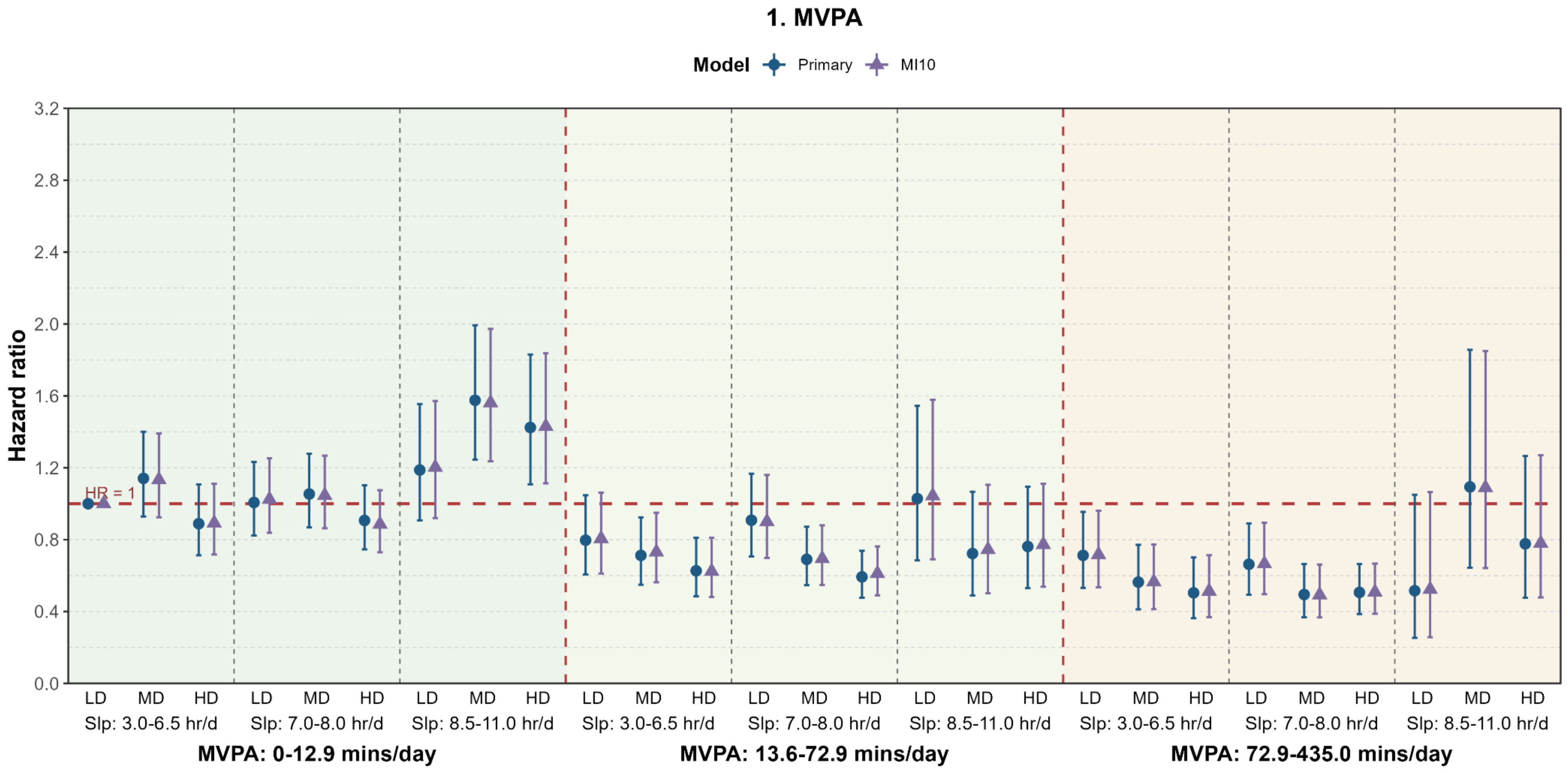


**Supplementary Figure 32 Multivariable-adjusted associations of combined sleep, moderate-to-vigorous physical activity (MVPA) and nutrition with all-cause mortality risk, using multiple imputation for 5 times (primary) and 10 times (MI10) (n = 31,875; events = 2,623). Legend:** We compared primary sample (using 5 times of multiple imputation) with 10 times of multiple imputation. Models are adjusted for age, sex, ethnicity, education, ratio of family income to poverty, alcohol consumption, smoking status, sedentary activity time, previous diagnosis of cardiovascular disease, cancer or type 2 diabetes, and family history of heart attack and diabetes. Sleep duration (hours/day), MVPA (minutes/day), and nutrition (Healthy Eating Index-2020 (HEI2020)) were treated as a joint term. The specific groups for each exposure were divided as sleep duration: 3.0-6.5 hours/day (short), 7.0-8.0 hours/day (optimal) and 8.5-11.0 hours/day (long); MVPA: 0-12.9 minutes/day (low), 12.9-72.9 minutes/day (moderate) and 72.9-435.0 minutes/day (high); HEI-2020: 10.0-45.0 (low), 45.0-56.0 (moderate), and 56.0-93.8 (high). The lowest group for all three exposures were treated as reference group. LD: low diet quality; MD: moderate diet quality; HD: high diet quality; Slp: sleep duration.

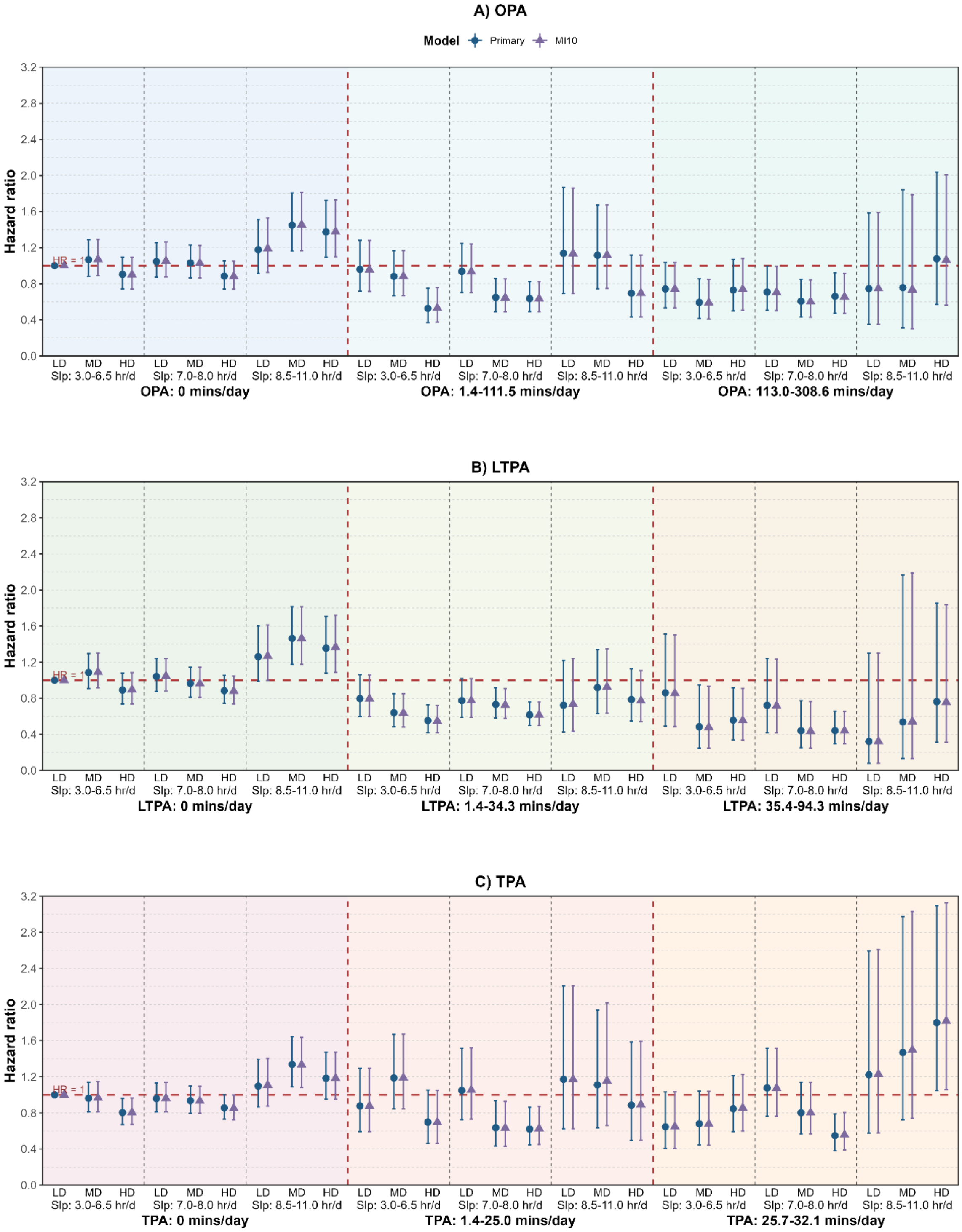


**Supplementary Figure 33 Multivariable-adjusted associations of combined sleep, physical activity domains and nutrition with all-cause mortality risk, using multiple imputation for 5 times (primary) and 10 times (MI10) (n = 31,875; events = 2,623). Legend:** We compared primary sample (using 5 times of multiple imputation) with 10 times of multiple imputation. The figure shows: A: Occupational physical activity (OPA); B: Leisure-time physical activity (LTPA); C: Transportational physical activity (TPA). Models are adjusted for adjusted for age, sex, ethnicity, education, ratio of family income to poverty, alcohol consumption, smoking status, sedentary activity time, previous diagnosis of cardiovascular disease, cancer or type 2 diabetes, and family history of heart attack and diabetes. When examining one PA domain as the primary exposure, the other two physical activity domains were adjusted in the multivariable models. Sleep duration (hours/day), OPA/LTPA/TPA (minutes/day), and nutrition were treated as a joint term. The specific groups for each exposure were divided as sleep duration: 3.0-6.5 hours/day (short), 7.0-8.0 hours/day (optimal) and 8.5-11.0 hours/day (long); OPA: 0 minutes/day (low), 1.4-111.5 minutes/day (moderate) and 113.0-308.6 minutes/day (high); LTPA: 0 minutes/day (low), 1.4-34.3 minutes/day (moderate) and 35.4-94.3 minutes/day (high); TPA: 0 minutes/day (low), 1.4-25.0 minutes/day (moderate) and 25.7-32.1 minutes/day (high); HEI-2020: 10.0-45.3 (low), 45.3-56.2 (moderate), and 56.2-93.8 (high). The lowest group for all three exposures in each physical activity domain were treated as reference group. LD: low diet quality; MD: moderate diet quality; HD: high diet quality; Slp: sleep duration.

Supplementary Table 1 STROBE statement

| | Item No | Recommendation | Page No |
|---|---|---|---|
| **Title and abstract** | 1 | (*a*) Indicate the study's design with a commonly used term in the title or the abstract | 1 |
| | | (*b*) Provide in the abstract an informative and balanced summary of what was done and what was found | 3-4 |
| **Introduction** | | | |
| Background/rationale | 2 | Explain the scientific background and rationale for the investigation being reported | 5-6 |
| Objectives | 3 | State specific objectives, including any prespecified hypotheses | 6 |
| **Methods** | | | |
| Study design | 4 | Present key elements of study design early in the paper | 7-8 |
| Setting | 5 | Describe the setting, locations, and relevant dates, including periods of recruitment, exposure, follow-up, and data collection | 7 |
| Participants | 6 | (*a*) Give the eligibility criteria, and the sources and methods of selection of participants. Describe methods of follow-up | 7 |
| | | (*b*) For matched studies, give matching criteria and number of exposed and unexposed | - |
| Variables | 7 | Clearly define all outcomes, exposures, predictors, potential confounders, and effect modifiers. Give diagnostic criteria, if applicable | 7-8 |
| Data sources/ measurement | 8* | For each variable of interest, give sources of data and details of methods of assessment (measurement). Describe comparability of assessment methods if there is more than one group | 7-8 |
| Bias | 9 | Describe any efforts to address potential sources of bias | 8-10 |
| Study size | 10 | Explain how the study size was arrived at | 7, 10 |
| Quantitative variables | 11 | Explain how quantitative variables were handled in the analyses. If applicable, describe which groupings were chosen and why | 7-10 |
| Statistical methods | 12 | (*a*) Describe all statistical methods, including those used to control for confounding | 8-10 |
| | | (*b*) Describe any methods used to examine subgroups and interactions | 8-10 |
| | | (*c*) Explain how missing data were addressed | 10 |
| | | (*d*) If applicable, explain how loss to follow-up was addressed | 7 |
| | | (*e*) Describe any sensitivity analyses | 10 |
| **Results** | | | |
| Participants | 13* | (a) Report numbers of individuals at each stage of study—eg numbers potentially eligible, examined for eligibility, confirmed eligible, included in the study, completing follow-up, and analysed | 11, Supplemental figure 1 |
| | | (b) Give reasons for non-participation at each stage | Supplemental figure 1 |
| | | (c) Consider use of a flow diagram | Supplemental figure 1 |
| Descriptive data | 14* | (a) Give characteristics of study participants (eg demographic, clinical, social) and information on exposures and potential confounders | Table 1 |
| | | (b) Indicate number of participants with missing data for each variable of interest | Supplemental figure 1 |
| | | (c) Summarise follow-up time (eg, average and total amount) | Table 1 |
| Outcome data | 15* | Report numbers of outcome events or summary measures over time | 11 |

| | | | |
|---|---|---|---|
| Main results | 16 | (*a*) Give unadjusted estimates and, if applicable, confounder-adjusted estimates and their precision (eg, 95% confidence interval). Make clear which confounders were adjusted for and why they were included | 11-13 |
| | | (*b*) Report category boundaries when continuous variables were categorized | 12-13 |
| | | (*c*) If relevant, consider translating estimates of relative risk into absolute risk for a meaningful time period | - |
| Other analyses | 17 | Report other analyses done—eg analyses of subgroups and interactions, and sensitivity analyses | 12-13 |
| **Discussion** | | | |
| Key results | 18 | Summarise key results with reference to study objectives | 14 |
| Limitations | 19 | Discuss limitations of the study, taking into account sources of potential bias or imprecision. Discuss both direction and magnitude of any potential bias | 17-19 |
| Interpretation | 20 | Give a cautious overall interpretation of results considering objectives, limitations, multiplicity of analyses, results from similar studies, and other relevant evidence | 14-19 |
| Generalisability | 21 | Discuss the generalisability (external validity) of the study results | 15, 17 |
| **Other information** | | | |
| Funding | 22 | Give the source of funding and the role of the funders for the present study and, if applicable, for the original study on which the present article is based | 21 |

* Information given separately for exposed and unexposed groups.

Supplementary Table 2. Variables extracted from NHANES

| Construct | Variables | Variable codes |
|---|---|---|
| **SPAN** | Sleep duration | SLD010H, SLD012 (2015-2018) |
| | Physical activity | PAQ605, PAQ610, PAD615, PAQ620, PAQ625, PAD630, PAQ635, PAQ640, PAD645, PAQ650, PAQ655, PAD660, PAQ665, PAQ670, PAD675 |
| | Diet quality | Sources: DR1TOT, DR2TOT, fped_dr1tot, fped_dr2tot |
| **Age** | Age | RIDAGEYR |
| **Sex** | Gender | RIAGENDR |
| **Ethnicity** | Ethnicity | RIDRETH1(we divided them as:<br>Non-Hispanic White<br>Non-Hispanic Black<br>Hispanic and Mexican American<br>Other) |
| **Socioeconomic background** | Education levels | DMDEDUC2 (we divided them as:<br>less than high school<br>high school or equivalent<br>college or above) |
| | Family poverty income ratio (PIR) | INDFMPIR |
| **Body fat** | BMI | BMXBMI |
| **Lifestyle behaviours** | Total Energy Intake | DR1TKCAL;<br>DR2TKCAL |
| | Alcohol Consumption | DR1TALCO<br>DR2TALCO (we divided them as:<br>Non-drinker<br>Moderate drinker<br>Heavy drinker) |
| | Smoking Status | SMQ020;<br>SMQ040 (we divided them as:<br>Never<br>Former<br>Current) |
| | Sedentary activity time<br>*(Include sitting at a desk, sitting with friends, traveling in a car, bus, or train, reading, playing cards, watching television, or using a computer)* | PAD680 |
| **Previous Diagnosis of Chronic Diseases** | Cardiovascular diseases | MCQ160B, MCQ160C, MCQ160D, MCQ160E, MCQ160F |
| | Cancers | MCQ220 |
| | Type 2 Diabetes | DIQ010 |
| **Family History of Chronic Diseases** | Heart Attack | MCQ300a |
| | Diabetes | MCQ300c |

Supplementary Table 3. Components of Health Eating Index-2020 and standards of scoring[4]

| Component | Maximum points | Standard for maximum score | Standard for minimum score of 0 |
|---|---|---|---|
| **Adequacy components** | | | |
| Total Fruits[b] | 5 | ≥0.8 cup eq/1,000 kcal | No Fruit |
| Whole Fruits[c] | 5 | ≥0.4 cup eq/1,000 kcal | No Whole Fruit |
| Total Vegetables[d] | 5 | ≥1.1 cup eq/1,000 kcal | No Vegetables |
| Greens and Beans[d] | 5 | ≥0.2 cup eq/1,000 kcal | No Dark Green Vegetables or Legumes |
| Whole Grains | 10 | ≥1.5 oz eq/1,000 kcal | No Whole Grains |
| Dairy[e] | 10 | ≥1.3 cup eq/1,000 kcal | No Dairy |
| Total Protein Foods[d] | 5 | ≥2.5 oz eq/1,000 kcal | No Protein Foods |
| Seafood and Plant Proteins[f] | 5 | ≥0.8 oz eq/1,000 kcal | No Seafood or Plant Proteins |
| Fatty Acids[g] | 10 | (PUFAs[h] + MUFAs[i]) / SFAs[j] ≥2.5 | (PUFAs + MUFAs) / SFAs ≤1.2 |
| **Moderation components** | | | |
| Refined Grains | 10 | ≤1.8 oz eq/1,000 kcal | ≥4.3 oz eq/1,000 kcal |
| Sodium | 10 | ≤1.1 g/1,000 kcal | ≥2.0 g/1,000 kcal |
| Added Sugars | 10 | ≤6.5% of energy | ≥26% of energy |
| Saturated Fats | 10 | ≤8% of energy | ≥16% of energy |

[a] Intake levels between the minimum and maximum standards are scored proportionately, and the component scores are then summed to derive the total score[4].

[b] Includes 100% fruit juice.

[c] Includes all forms excluded juice.

[d] Includes beans, peas, and lentils.

[e] Includes all milk products, such as fluid milk, yogurt, and cheese, and fortified soy beverages.

[f] Includes seafood, nuts, seeds, soy products (other than beverages), and beans, peas, and lentils.

[g] Ratio of poly- and monounsaturated fatty acids (PUFAs and MUFAs) to saturated fatty acids (SFAs).

[h] PUFA = polyunsaturated fatty acid.

[i] MUFA = monounsaturated fatty acid.

[j] SFA = saturated fatty acid.

**Supplementary Table 4** Sample characteristics of individuals in the primary analysis sample using multiple imputations versus individuals with incomplete or missing data.

| | Primary Sample using MI | Sample with missing data |
|---|---|---|
| **Sample** | 31875 | 31875 |
| **Events** | 2623 | 2623 |
| **Follow up, months** (median [IQR]) | 81.00 [46.00, 118.00] | 81.00 [46.00, 118.00] |
| **Age, years** (median [IQR]) | 48.00 [32.00, 63.00] | 48.00 [32.00, 63.00] |
| **Sex, n (%)** | | |
| Male | 15504 (48.6) | 15504 (48.6) |
| Female | 16371 (51.4) | 16371 (51.4) |
| **Race, n (%)** | | |
| Non-Hispanic White | 13021 (40.9) | 13021 (40.9) |
| Non-Hispanic Black | 6899 (21.6) | 6899 (21.6) |
| Hispanic and Mexican American | 8283 (26.0) | 8283 (26.0) |
| Other | 3672 (11.5) | 3672 (11.5) |
| **Education, n (%)** | | |
| Less than high school | 7631 (23.9) | 7298 (22.9) |
| High school or equivalent | 7267 (22.8) | 6925 (21.7) |
| College or above | 16977 (53.3) | 16005 (50.2) |
| Missing | NA | 1647 (5.2) |
| **PIR, n (%)** | | |
| <1.3 | 10590 (33.2) | 9524 (29.9) |
| 1.3-3.5 | 11969 (37.5) | 10900 (34.2) |
| ≥3.5 | 9316 (29.2) | 8575 (26.9) |
| Missing | NA | 2876 (9.0) |
| **BMI** (median [IQR]) | 28.00 [24.20, 32.65] | 28.00 [24.20, 32.65] |
| **Sedentary time, mins/day** (median [IQR]) | 300.00 [180.00, 480.00] | 300.00 [180.00, 480.00] |
| **Total energy intake, kcal/day** (median [IQR]) | 1903.00 [1448.50, 2471.00] | 1903.00 [1448.50, 2471.00] |
| **Sleep, hours/day** (median [IQR]) | 7.00 [6.00, 8.00] | 7.00 [6.00, 8.00] |
| **HEI-2020** (median [IQR]) | 50.25 [42.21, 59.27] | 50.25 [42.21, 59.27] |
| **MVPA, mins/day** (median [IQR]) | 34.29 [0.00, 107.14] | 34.29 [0.00, 107.14] |
| **OPA, mins/day** (median [IQR]) | 0.00 [0.00, 68.57] | 0.00 [0.00, 68.57] |
| **LTPA, mins/day** (median [IQR]) | 0.00 [0.00, 34.29] | 0.00 [0.00, 34.29] |
| **TPA, mins/day** (median [IQR]) | 0.00 [0.00, 4.29] | 0.00 [0.00, 4.29] |
| **Smoke status, n (%)** | | |
| Never | 18225 (57.2) | 17583 (55.2) |
| Former | 7328 (23.0) | 7265 (22.8) |
| Current | 6322 (19.8) | 6184 (19.4) |
| Missing | NA | 843 (2.6) |
| **Alcohol consumption, n (%)** | | |
| Non-drinker | 23127 (72.6) | 23127 (72.6) |
| Moderate drinker | 4548 (14.3) | 4548 (14.3) |
| Heavy drinker | 4200 (13.2) | 4200 (13.2) |
| **Previous CVD, n (%)** | | |
| No | 28615 (89.8) | 26896 (84.4) |
| Yes | 3260 (10.2) | 3226 (10.1) |

| | | |
|---|---|---|
| Missing | NA | 1753 (5.5) |
| **Previous cancer, n (%)** | | |
| No | 28946 (90.8) | 27322 (85.7) |
| Yes | 2929 (9.2) | 2911 (9.1) |
| Missing | NA | 1642 (5.2) |
| **Previous T2D, n (%)** | | |
| No | 27883 (87.5) | 27867 (87.4) |
| Yes | 3992 (12.5) | 3992 (12.5) |
| Missing | NA | 16 (0.1) |
| **Family history of diabetes, n (%)** | | |
| No | 18540 (58.2) | 17043 (53.5) |
| Yes | 13335 (41.8) | 12631 (39.6) |
| Missing | NA | 2201 (6.9) |
| **Family history of heart attack, n (%)** | | |
| No | 28088 (88.1) | 25850 (81.1) |
| Yes | 3787 (11.9) | 3657 (11.5) |
| Missing | NA | 2368 (7.4) |

Note: MI: Multiple Imputation; NA: not applicable; MVPA: moderate-to-vigorous physical activity; OPA: occupational physical activity; LTPA: leisure-time physical activity; TPA: transportational physical activity; CVD: cardiovascular disease; T2D: type 2 diabetes; HEI-2020: Healthy Eating Index-2020.
